# Prevalence of cancer-related cognitive impairment in pediatric patients and survivors of non-CNS cancers: A systematic review and meta-analysis

**DOI:** 10.1101/2025.01.22.25320991

**Authors:** Ines Semendric, Timothy H. Barker, Olivia J. Haller, Danielle Pollock, Lyndsey E. Collins-Praino, Alexandra L. Whittaker

## Abstract

**Background:** Neurocognitive toxicity is a common sequela of cancer and its treatment, with 40-60% of childhood cancer survivors experiencing cancer-related cognitive impairment (CRCI) following cancer and its treatment. CNS cancers and CNS-directed therapy are considered to pose the greatest risk; however, such impairment has also been reported in non-CNS cancers, the most commonly diagnosed in children. Despite initial findings, no formal systematic assimilation of evidence has previously reported estimates in pediatric populations, or identified the influence of cancer, treatment, and patient variables. Thus, this review aims to investigate the prevalence of CRCI in pediatric non-CNS cancers.

**Methods:** A systematic search was conducted using PubMed via MEDLINE, Scopus, PsycINFO via OvidSP, and CINAHL via EBSCOhost. Simple proportional meta-analyses were performed using PERSyst, and the certainty of evidence was assessed employing GRADE.

**Results:** Twenty-six studies contributed to two cognitive domains - (1) global cognitive function and intelligence, and (2) achievement, aptitude and school readiness. Pooled prevalence estimates suggest that survivors of non-CNS cancers experience impairments in measures of intelligence (12.46%-18.05%) and attention (9.83%-12.05%). Non-pooled prevalence estimates ranged from 0-67% in global cognitive function and intelligence, and 0-73% in achievement, aptitude and school readiness.

**Conclusion:** Survivors of non-CNS cancers experience significant impairments in these cognitive domains; however, certainty of evidence is generally low to very low, warranting cautious interpretation of prevalence estimates.

**Implication for Cancer Survivors:** Understanding the prevalence of CRCI following childhood cancer can inform survivorship strategies and provide impetus for provision of dedicated supports, such as rehabilitation and school reintegration supports.

## Introduction

The most common cancer diagnosed in childhood are leukemias, particularly acute lymphoblastic leukemia (ALL), representing 78% of childhood leukemias [1]. Age is a strong influencing factor for cancer occurrence, with disease predominantly occurring under the age of 5 years, at a time when there is rapid brain development [2, 3]. A second peak occurs during adolescence [1], another period of rapid brain development (for review, see Konrad et al. [4]). As a result of advances in detection and treatment, the 5-year survival rate of childhood cancers is 85-90% [5]. As a result, issues of survivorship have risen to the forefront.

Neurocognitive toxicity is a common sequela of cancer and its treatment, with suggestions that between 40%-60% of children who are childhood cancer survivors may experience symptoms of this toxicity, with a consequent impact on their quality of life [6]. In many cases, neurocognitive toxicity is the result of combined polychemotherapy and radiation treatments [7]. However, recent longitudinal studies in childhood ALL patients aged 2 to 12 years old treated with polychemotherapy alone also demonstrated similar outcomes. Specifically, patients demonstrated lower performance IQ (intelligence quotient) scores at neuropsychological follow-up 3 and 6 years post-diagnosis (n = 8) or at 6 and 9 years (n = 2)) post-diagnosis compared to their baseline assessment conducted at age 6 or at diagnosis, whichever occurred first [8]. Furthermore, it is not just cancer treatments that can cause cognitive impairment [7]. It is now recognized that cancer itself can cause impairment, with the predominately adult clinical literature suggesting that between 20-50% of patients may experience such impairment prior to undergoing treatment [9–12]. Of note, adult patients with non-CNS cancers have also been shown to suffer issues with cognition, even in the absence of treatment [9, 13]. This may be caused by direct effects of the tumor itself, as a result of associated co-morbidities, or due to psychological factors, such as worry and fatigue [14]. Specifically, higher worry has been associated with reduced performance in response to higher task demand in pretreatment chemotherapy and pretreatment radiation breast cancer patients, due to the inability to deactivate the precuneus/posterior cingulate region of the default mode network [14]. In recognition of the contribution of cancer itself to cognitive change, the presence of cognitive decline post diagnosis and treatment is known as cancer-related cognitive impairment (CRCI), as opposed to the more colloquially used term ‘chemobrain’.

CRCI symptoms vary in severity and duration [10, 15]. Moreover, diverse cognitive domains may be affected, including facets of memory, processing speed, attention and executive function [16]. The resulting cognitive disability has a significant negative impact on patients’ quality of life, leading to reduced ability to multi-task, perform everyday activities, spatially navigate and remember familiar concepts, such as names of individuals [17, 18]. Together, these functional deficits lead to challenges in participating in work and study [17, 18]. In line with this, many pediatric cancer survivors require special education programs, experience extended absences, repeat (or fail) courses/classes at school, and demonstrate difficulty with multiple subjects, including mathematics, English, and science [19]. Similarly, they achieve lower marks and levels of educational attainment, exhibit behavioural problems at school, and have lower IQ scores compared to healthy children their age [20, 21]. In comparison to their healthy siblings, these children are 3-4 times more likely to suffer from learning disabilities and display deficits in social skills and development [22]. Consequently, survivors of childhood cancer also experience higher risk of job discrimination and lower levels of career success over their lifetime [19]. The St Jude Lifetime Cohort Study has demonstrated that CRCI can also pose long-term challenges, with adult survivors of childhood cancer reporting cognitive impairment up to 26-years post diagnosis and treatment [23]. Impairment rates ranged from 28.6% to 58.9% across measures of intelligence, academic skills, attention, memory, processing speed and executive function in survivors who received chemotherapy and/or cranial radiation therapy [23]. Of significance, with increased time since diagnosis, there was an increased risk for executive function impairment.

This may be a consequence of the fact that executive functions continue to develop well into early adulthood, with early injury potentially disrupting this expected development trajectory [23]. Further, age at time of treatment may also influence outcomes, with a younger age at time of treatment posing a higher risk of the subsequent development of cognitive deficits [23].

Importantly, survivors highlighted that impairments extending into adulthood impacted their ability to attain and/or maintain education or employment, become financially independent, engage in social and family life and maintain psychosocial wellbeing [23].

Despite these previous findings, key questions remain, particularly around the prevalence of CRCI in pediatric cancer survivors. Prevalence refers to the proportion of the population that has a condition at a particular time, typically used to demonstrate the overall burden of a condition in a population [24]. Prevalence rates for CRCI can be influenced by risk factors, which include genetics, female sex, younger age at diagnosis, type of cancer, associated psychological distress, chemotherapy dose and both dose and field size for radiation [25]. Based on data from female breast cancer survivors, prevalence rates may also be influenced by study design differences, such as the population size sampled, method used to diagnose cognitive impairment, time since cessation of chemotherapy and use of longitudinal versus cross-sectional designs [26]. For example, higher prevalence rates were associated with lower sample sizes, with self-reporting of cognitive complaints, and in those either currently undergoing treatment or just recently finishing therapy. These factors likely also contribute to the variable prevalence rates reported for pediatric patients in the literature to date. For example, it has been estimated that 40-100% of childhood brain tumor survivors experience neurocognitive problems [27, 28]. Conversely, childhood ALL patients treated with CNS prophylactic therapy, with or without cranial radiation, experienced impairment at the rate of 33% and 25%, respectively [23, 29]. In another study, pediatric ALL survivors that were treated with radio-chemotherapy were shown to have attention deficits in 67% of cases and memory deficits in 3–28% of cases [30, 31]. Despite this, how individual risk factors may influence prevalence rates of cognitive impairment following childhood cancer remains largely unexplored.

It is widely recognized that there is an inherently high risk of long-term neurocognitive impairments arising from CNS cancers, which are the second most common type of childhood cancer following leukaemias [11, 12, 32]. In comparison, CNS involvement is not implicated in the majority of acute lymphoblastic leukaemia cases, particularly at diagnosis, suggesting that the inclusion of CNS cancers may skew prevalence rates [33]. As such, it is important to investigate the contribution of CNS and non-CNS cancers towards the development and presentation of CRCI independently. Understanding the prevalence of CRCI following non-CNS childhood cancer, and the factors that influence risk of CRCI development, will help inform survivorship strategies for this population and provide impetus for provision of dedicated supports, such as rehabilitation and additional assistance for patients upon school return.

With these questions in mind, a preliminary search of PROSPERO, PubMed via MEDLINE, the Cochrane Database of Systematic Reviews and *JBI Evidence Synthesis* was conducted and no current or prospective systematic reviews on the topic were identified. Thus, this review aims to address this gap and investigate the prevalence of CRCI in pediatric patients and survivors of non-CNS cancers. An additional focus will be on subgroup analyses where there are sufficient data to investigate how individual factors may influence and contribute to prevalence rates.

### Review question

What is the prevalence of CRCI following cancer and its treatment in paediatric patients and survivors of non-CNS cancers?

## Methods

The review was conducted in accordance with the JBI methodology for systematic reviews of prevalence and incidence [34]. This review has been conducted in accordance with an *a priori* protocol registered with PROSPERO (CRD42022321930). This review has been prepared following the Preferred Reporting Items for Systematic reviews and Meta-Analyses (PRISMA) 2020 reporting guidelines **(Online Resource 1)** [35].

### Inclusion criteria Participants

Studies were eligible if they included data on children from the age of 0 to 18 years old who either have had or currently have any non-CNS cancer and have either undergone or are currently undergoing any form of common cancer treatment. Common cancer treatments were defined as including surgery, radiation and/or chemotherapy, alone or in combination. Studies assessing alternative treatments, such as traditional or complementary medicines, were not included. Participants over the age of 18 years old who had cancer treatment as either a child (0-11) or adolescent (12-18) were eligible for inclusion. Reports of cognitive function in pediatric patients who have cancer, but who have not yet received any cancer-related treatments, were also eligible for inclusion. There were no restrictions on time since cessation of treatment. Individuals with cancers with central nervous system involvement were not included. Whilst these are the second most common type of childhood cancer, there is an inherently high risk of long-term neurocognitive impairments as a result of the tumor itself, which may skew prevalence rates for non-CNS cancers [11, 12, 32]. CNS-directed treatment in the absence of CNS involvement, such as CNS prophylaxis, was included, as the use of intrathecal chemotherapy and/or irradiation is standard procedure to minimize the risk of patients developing CNS relapse in the future.

### Condition

Cognitive decline following cancer has been variously labelled as: Chemotherapy-induced cognitive impairment (CICI); cancer-treatment cognitive impairment (CTCI); CRCI; or, colloquially, as ‘chemofog’ or ‘chemobrain’. For the purposes of this review, we will use the terminology CRCI. This review considered studies that report on the prevalence of CRCI in pediatric patients and survivors of non-CNS cancers. This includes cognitive impairment that may result from the cancer alone, as well as those arising as a result of neurotoxicity due to common treatments. Methods of diagnosis include either self-report (or report by a parent/teacher/other caregiver) or formal neurocognitive testing. Currently, there are no universally validated and adopted tools to measure or diagnose CRCI; however, tools designed to measure aspects of cognitive function affected in cancer patients/survivors were included [36].

### Context

This review considered studies where the target population were at home, or in hospital or out-patient settings. Patients in hospices were excluded, due to differences in the treatment pathways and treatment regimens in patients receiving palliative care [37]. Studies were not restricted by geographic location.

### Types of studies

Analytical observational studies, including longitudinal cohort studies and cross-sectional studies, were considered for inclusion. Randomized controlled trials were included if studies reported baseline cognitive measures of eligible populations.

### Search strategy

An initial search of PubMed Central, Scopus, PsycINFO via OvidSP, and CINAHL via EBSCOhost was conducted in July 2022 to identify eligible peer-review publications. The text words contained in the titles and abstracts of relevant articles, and the index terms used to describe the articles, were used to identify appropriate key terms and controlled vocabulary to develop a draft search strategy for all included databases. In conjunction with the Faculty of Health and Medical Science’s research library liaison, Vikki Langton, the search strategy was refined and finalized **(Online Resource 2)**. The initial search was conducted in April 2023 and included four key search terms comprised of variations of “children”, “non-CNS cancer”, “cancer treatment”, and “cognitive impairment”. Examples of synonyms/free-text words for each concept identified include child, young adult, neoplasm, leukemia, chemotherapy, CICI and CRCI. The search strategy, including all identified free-text words and index terms, was individually adapted for PubMed, Scopus, PsycINFO via OvidSP, and CINAHL via EBSCOhost. The reference lists of all included studies were hand searched and screened for additional studies. Forward citation searching was also conducted where studies contributed to broader cohort studies by employing the “cited by” function. Theses and dissertations were included for academic rigor. Other gray literature, such as conference papers/proceedings, were excluded, unless an associated full-text publication was available upon searching. Only English-language publications were included, due to lack of ready accessibility to a translator. An updated search following the same procedure was conducted a year following the original search, in June 2024.

### Study selection

Following searches of included databases, all identified records were collated and uploaded into EndNote v.X9 (Clarivate Analytics, PA, USA), then imported into Covidence systematic review software (Veritas Health Innovation, Melbourne, Australia), where duplicates were automatically removed. Any remaining duplicates were manually identified and removed by reviewers through the screening process. All stages of the screening process were conducted in Covidence. Each stage of the screening process was piloted by assigning a random subset of studies (10 studies per reviewer) to three independent reviewers (IS, OJH, ALW) (Fleiss’ Kappa 0.82). Where conflicts arose on consensus at any stage of the screening process, it was resolved through discussion, or consultation with an additional independent reviewer (ALW). Reasons for exclusion at the full-text stage were recorded and presented. The results of all stages of the identification and removal process are reported in full in the PRISMA flow diagram [35] **(Figure 1).**

**Figure 1.**
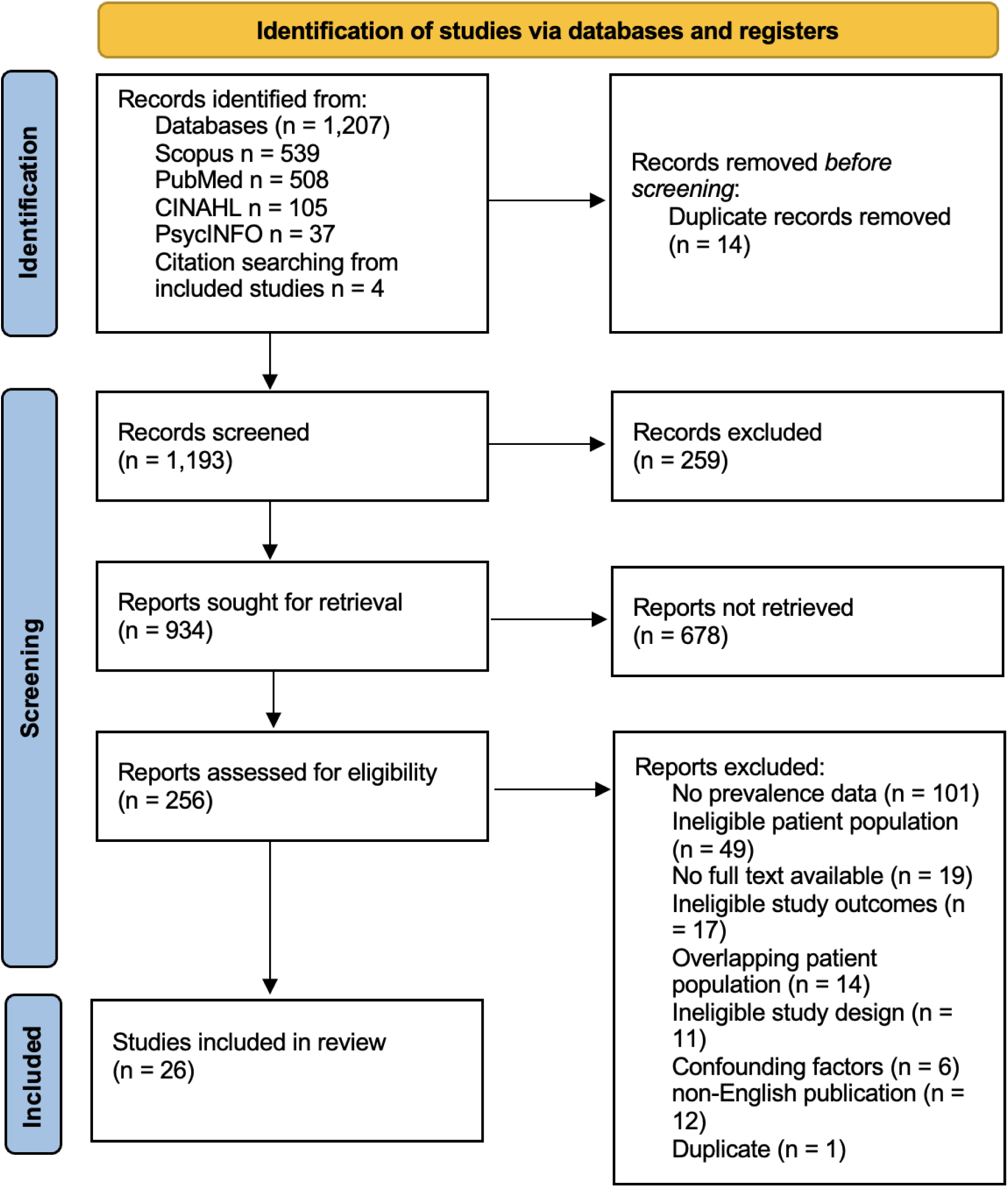
PRISMA 2020 flow diagram for the systematic review detailing the databases searched, the number of abstracts screened and full texts retrieved [35].

### Assessment of methodological quality

Eligible studies were critically appraised by two independent reviewers (IS, ALW) at the study level for methodological quality. The JBI checklist for prevalence studies was utilized for all prevalence data, regardless of study design **(Online Resource 3)** [38]. Conflicts in assessment of methodological quality were resolved through discussion or by consultation with a third independent reviewer. The results are reported narratively and tabulated. A study was rated as high quality if it scored greater than 70%, moderate if it scored between 50-70%, and low if it scored below 50%, as described in Dijkshoorn et al 2021 [39]. All studies, regardless of the results of their methodological quality, underwent data extraction and synthesis where possible.

### Data extraction

Data were extracted from studies included in the review by two independent reviewers (IS, ALW) using a previously modified version of the Covidence data extraction tool 2.0 in Covidence **(Online Resource 4)** [26]. Data extraction consistency between two reviewers was piloted on a randomly selected sample of studies (*n* = 10) prior to full extraction taking place (IS, OJH) (Cohen’s Kappa 0.75). Data extracted included study characteristics and specific details about the condition, population (e.g. sex, age), context, age at study evaluation, type of cancer (e.g. acute lymphoblastic leukemia), age at cancer diagnosis, age at treatment, time since cancer diagnosis, time since cessation of treatment, method of assessment of prevalence, and summary prevalence estimates. Prevalence data were extracted as a percent or converted to this using numerator/denominator information for the number of impacted survivors and sample size.

Confidence intervals (CIs) were extracted where available.

### Data synthesis

An initial descriptive analysis was conducted by tabulating the study characteristics via Microsoft Excel and comparing against the planned sub-groups of cancer type, age at diagnosis (less than 5 versus older than 5), time since diagnosis, time since treatment and/or cessation of treatment, and methods of diagnosis of cognitive impairment (neuropsychological testing versus self-report). Key study characteristics are presented in tabular form and narratively described.

Synthesis was conducted narratively where there was heterogeneity in study characteristics. Where possible, sufficiently homogenous studies were grouped to perform a meta-analysis using the PERSyst-MA tool version 1.0, an online application for R package meta, version 7.0-0.

These data were assimilated separately (i.e. prevalence is presented either narratively or via meta-analysis).

### Meta Analysis

For each available grouping of studies (described above), a simple proportional meta-analysis method was conducted using the PERSyst-MA tool version 1.0, an online application for R package meta (version 7.0-0) to provide an estimate of the prevalence across the included studies [40]. Meta analyses are presented via a forest plot with accompanying narrative synthesis.

Where >5 studies were included in the meta-analysis, overall estimates of effect were calculated with the random effects models employing the DerSimonian & Laird method as a between study variance estimator. Where <5 studies were included in the meta-analysis, overall estimates of effect were calculated with a fixed/common effects model. The meta-analysis employed the inverse of variance method with the logit link function for the analysis of proportion data.

Inverse variance assigns weights to each study based on the inverse of the variance of the effect estimate (or one over the square of its standard error). More weight is assigned to larger studies with lower standard errors compared to smaller studies with larger standard errors to minimise imprecision of pooled effect estimates [41]. Transformation of proportions was done using the Logit transformation. Heterogeneity was quantified using restricted maximum-likelihood as the variance estimator. Cochran’s Q verified the presence of heterogeneity, whilst the I^2^ statistic are presented as the proportion of observed variance that is not due to sampling error [42, 43].

Prediction intervals are additionally presented to inform the range of expected estimates and presented alongside Cochran’s Q, I^2^, and the 95% confidence interval. Overall heterogeneity was assessed according to the Prevalence Estimates Reviews-Systematic Reviews Methodology Group’s (PERSyst) guidelines [43]. Sensitivity analysis, in the form of subgroup analysis, was conducted in accordance with predetermined hypotheses, as stated in the *a priori* protocol. The Q-Profile method was employed to determine significance between subgroups in subgroup analysis, with a *p* value of <0.05 considered significant.

Subgroup analyses were conducted where there was sufficient data to investigate how these may influence prevalence rates. The subgroups identified and assessed included cancer type, CNS-directed versus non-CNS directed therapy, time since cessation of treatment, and outcome severity. Where no subgroups were available, if appropriate, overall prevalence estimates were calculated and reported. Where too few studies remained in each subgroup to use meta-analytic techniques, these studies were synthesized narratively and in tabular form, with simple descriptive statistics conducted such as median/range. These data were presented based on the subgroups described above. Publication bias was assessed using doi plots and the Luis Furuya-Kanamori asymmetry index (LFK index) via the Megastat Excel Add-In (version 10.3, McGraw-Hill Higher Education, New York, NY). Where smaller number of studies are included in a meta-analysis, the doi plot is considered to be more sensitive than the funnel plot [44].

Qualitative considerations were narratively reported including language bias, availability bias, cost bias, familiarity bias and outcome bias.

### Assessing certainty in the evidence

Certainty in the evidence was assessed employing the Grading of Recommendations, Assessment, Development and Evaluation (GRADE) tool for assessment of evidence of prognosis [45]. Each outcome from the included studies was assigned a ranking to reflect the quality of evidence (very low, low, moderate, and high). The assigned ranking considered risk of bias, precision of effect estimates, consistency of individual study outcomes, if the evidence directly answered the question of interest and publication/reporting bias. This process was piloted by assigning an outcome grouping to two independent reviewers (IS, ALW). Consensus between the reviewers was attained prior to undertaking this stage. Where conflicts arose on consensus, it was resolved through discussion, or the consultation of an additional independent reviewer (THB). A Summary of Findings (SoF) table was created through GRADEPro GDT software (McMaster University, ON, Canada) and presented **(Table 1)** [46]. The SoF table reported the review title, population, phenomena of interest, context, outcomes, estimated risk, relative effect, number of participants, quality of the evidence (GRADE) and any relevant comments.

**Table 1.**
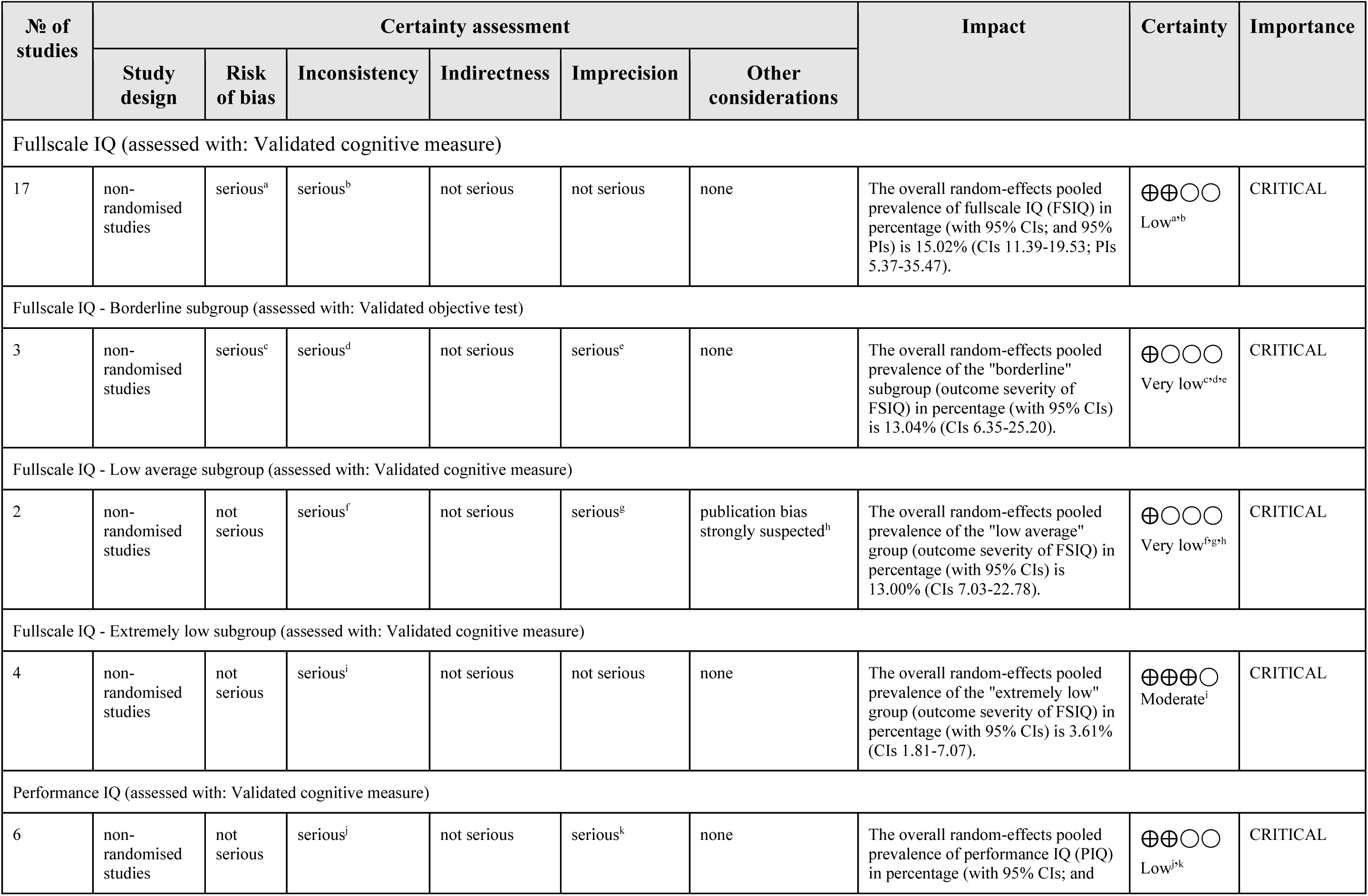
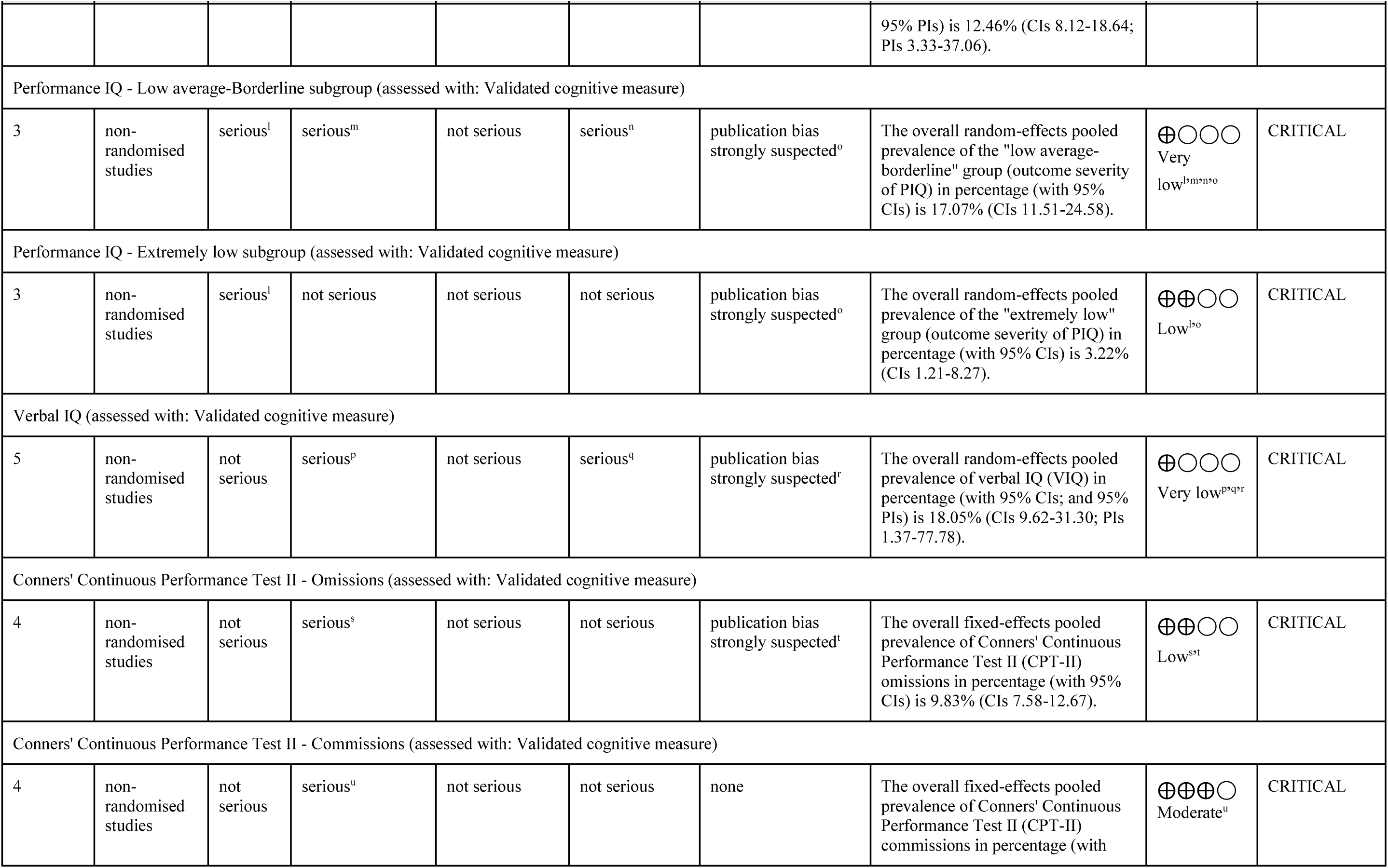
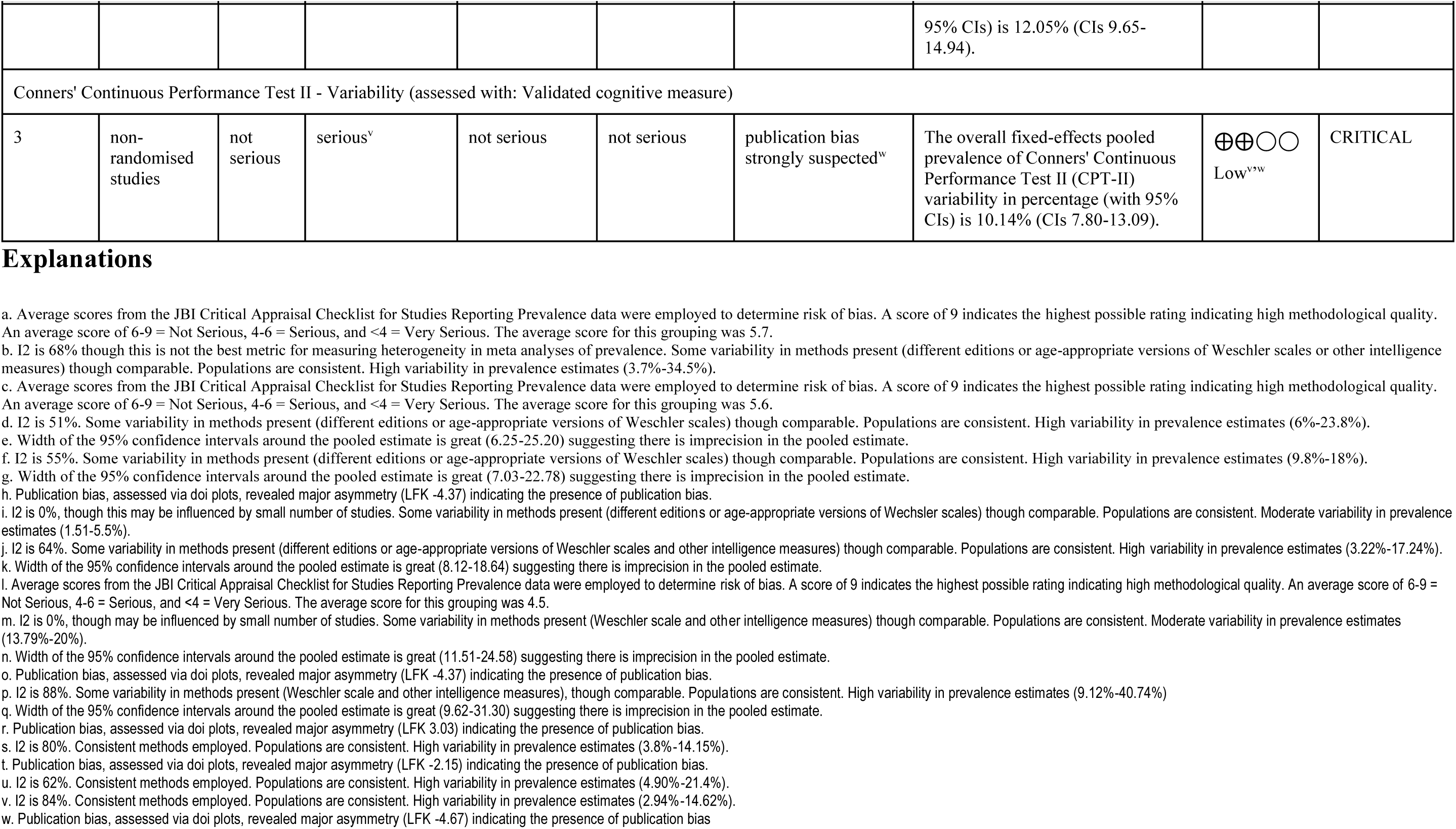
GRADE Summary of Findings table for prevalence as classified by cognitive domain.

| № of studies | Certainty assessment |  |  |  |  |  | Impact | Certainty | Importance |
| --- | --- | --- | --- | --- | --- | --- | --- | --- | --- |
|  | Study design | Risk of bias | Inconsistency | Indirectness | Imprecision | Other considerations |  |  |  |
| Fullscale IQ (assessed with: Validated cognitive measure) |  |  |  |  |  |  |  |  |  |
| 17 | non-randomised studies | serious<sup>a</sup> | serious<sup>b</sup> | not serious | not serious | none | The overall random-effects pooled prevalence of fullscale IQ (FSIQ) in percentage (with 95% CIs; and 95% PIs) is 15.02% (CIs 11.39-19.53; PIs 5.37-35.47). | ⊕⊕○○<br>Low<sup>a,b</sup> | CRITICAL |
| Fullscale IQ - Borderline subgroup (assessed with: Validated objective test) |  |  |  |  |  |  |  |  |  |
| 3 | non-randomised studies | serious<sup>c</sup> | serious<sup>d</sup> | not serious | serious<sup>e</sup> | none | The overall random-effects pooled prevalence of the "borderline" subgroup (outcome severity of FSIQ) in percentage (with 95% CIs) is 13.04% (CIs 6.35-25.20). | ⊕○○○<br>Very low<sup>c,d,e</sup> | CRITICAL |
| Fullscale IQ - Low average subgroup (assessed with: Validated cognitive measure) |  |  |  |  |  |  |  |  |  |
| 2 | non-randomised studies | not serious | serious<sup>f</sup> | not serious | serious<sup>g</sup> | publication bias strongly suspected<sup>h</sup> | The overall random-effects pooled prevalence of the "low average" group (outcome severity of FSIQ) in percentage (with 95% CIs) is 13.00% (CIs 7.03-22.78). | ⊕○○○<br>Very low<sup>f,g,h</sup> | CRITICAL |
| Fullscale IQ - Extremely low subgroup (assessed with: Validated cognitive measure) |  |  |  |  |  |  |  |  |  |
| 4 | non-randomised studies | not serious | serious<sup>i</sup> | not serious | not serious | none | The overall random-effects pooled prevalence of the "extremely low" group (outcome severity of FSIQ) in percentage (with 95% CIs) is 3.61% (CIs 1.81-7.07). | ⊕⊕⊕○<br>Moderate<sup>i</sup> | CRITICAL |
| Performance IQ (assessed with: Validated cognitive measure) |  |  |  |  |  |  |  |  |  |
| 6 | non-randomised studies | not serious | serious<sup>j</sup> | not serious | serious<sup>k</sup> | none | The overall random-effects pooled prevalence of performance IQ (PIQ) in percentage (with 95% CIs; and | ⊕⊕○○<br>Low<sup>j,k</sup> | CRITICAL |
|  | Study design | Risk of bias | Inconsistency | Indirectness | Imprecision | Other considerations |  |  |  |
|  |  |  |  |  |  |  | 95% PIs) is 12.46% (CIs 8.12-18.64; PIs 3.33-37.06). |  |  |
| Performance IQ - Low average-Borderline subgroup (assessed with: Validated cognitive measure) |  |  |  |  |  |  |  |  |  |
| 3 | non-randomised studies | serious<sup>l</sup> | serious<sup>m</sup> | not serious | serious<sup>n</sup> | publication bias strongly suspected<sup>o</sup> | The overall random-effects pooled prevalence of the "low average-borderline" group (outcome severity of PIQ) in percentage (with 95% CIs) is 17.07% (CIs 11.51-24.58). | ⊕○○○<br>Very low<sup>l,m,n,o</sup> | CRITICAL |
| Performance IQ - Extremely low subgroup (assessed with: Validated cognitive measure) |  |  |  |  |  |  |  |  |  |
| 3 | non-randomised studies | serious<sup>l</sup> | not serious | not serious | not serious | publication bias strongly suspected<sup>o</sup> | The overall random-effects pooled prevalence of the "extremely low" group (outcome severity of PIQ) in percentage (with 95% CIs) is 3.22% (CIs 1.21-8.27). | ⊕⊕○○<br>Low<sup>l,o</sup> | CRITICAL |
| Verbal IQ (assessed with: Validated cognitive measure) |  |  |  |  |  |  |  |  |  |
| 5 | non-randomised studies | not serious | serious<sup>p</sup> | not serious | serious<sup>q</sup> | publication bias strongly suspected<sup>r</sup> | The overall random-effects pooled prevalence of verbal IQ (VIQ) in percentage (with 95% CIs; and 95% PIs) is 18.05% (CIs 9.62-31.30; PIs 1.37-77.78). | ⊕○○○<br>Very low<sup>p,q,r</sup> | CRITICAL |
| Conners' Continuous Performance Test II - Omissions (assessed with: Validated cognitive measure) |  |  |  |  |  |  |  |  |  |
| 4 | non-randomised studies | not serious | serious<sup>s</sup> | not serious | not serious | publication bias strongly suspected<sup>t</sup> | The overall fixed-effects pooled prevalence of Conners' Continuous Performance Test II (CPT-II) omissions in percentage (with 95% CIs) is 9.83% (CIs 7.58-12.67). | ⊕⊕○○<br>Low<sup>s,t</sup> | CRITICAL |
| Conners' Continuous Performance Test II - Commissions (assessed with: Validated cognitive measure) |  |  |  |  |  |  |  |  |  |
| 4 | non-randomised studies | not serious | serious<sup>u</sup> | not serious | not serious | none | The overall fixed-effects pooled prevalence of Conners' Continuous Performance Test II (CPT-II) commissions in percentage (with | ⊕⊕⊕○<br>Moderate<sup>u</sup> | CRITICAL |
|  | Study design | Risk of bias | Inconsistency | Indirectness | Imprecision | Other considerations |  |  |  |
|  |  |  |  |  |  |  | 95% CIs) is 12.05% (CIs 9.65-14.94). |  |  |
| Conners' Continuous Performance Test II - Variability (assessed with: Validated cognitive measure) |  |  |  |  |  |  |  |  |  |
| 3 | non-randomised studies | not serious | serious<sup>v</sup> | not serious | not serious | publication bias strongly suspected<sup>w</sup> | The overall fixed-effects pooled prevalence of Conners' Continuous Performance Test II (CPT-II) variability in percentage (with 95% CIs) is 10.14% (CIs 7.80-13.09). | ⊕⊕○○<br>Low<sup>v,w</sup> | CRITICAL |

## Results

### Search results

The original database search conducted in April 2023, with an updated search conducted a year later in August 2024, identified a total of 1,207 studies with 273 studies excluded as duplicates (*n* = 14 manually removed, *n* = 259 removed by Covidence automation). A total of 678 studies were excluded based on title and abstract screening, with 256 remaining for full-text assessment. During the full-text screening, 230 studies were excluded due to the following reasons: no prevalence data reported (*n* = 101), ineligible participant population (*n* = 49), no full-text available (*n* = 19), ineligible study outcomes (*n* = 17), overlapping patient populations/cohort studies (*n* = 14), ineligible study design (*n* = 11), confounding factors (*n* = 6), non-English publications (*n* = 12) and being a duplicate (*n* = 1). In total, 26 studies were deemed eligible for inclusion in this review.

Details of the identification process are described in the Preferred Reporting Items for Systematic Reviews and Meta-Analyses (PRISMA 2020) flow chart **(Figure 1)** [35].

### Included studies

The 26 included studies contributed 217 prevalence estimates. It should be noted that none of the included studies sought to investigate prevalence as a primary aim, but did report prevalence estimates within their reported outcomes. Publication dates ranged from 1981 to 2023. Data were provided from the USA (*n* = 14), Germany (*n* = 2), Finland (*n* = 2), United Kingdom (*n* = 1), China (*n* = 1), Taiwan (*n* = 1), Spain (*n* = 1), Italy (*n* = 1), Poland (*n* = 1), Norway (*n* = 1) and Switzerland (*n =* 1). Included studies were primarily observational in nature, with cross-sectional design (*n =* 7), cohort studies (*n =* 9), case series and case-control studies (*n =* 4), longitudinal (*n =* 4), and correlational studies (*n =* 1). Additionally, one experimental study was included with randomized controlled trial design (*n =* 1). Sample sizes ranged from *n* = 16 to *n* = 350, with a mean sample size of 80. All studies (*n* = 26) employed objective test modalities. The characteristics of the included studies are reported in full (see Table 2).

**Table 2.**
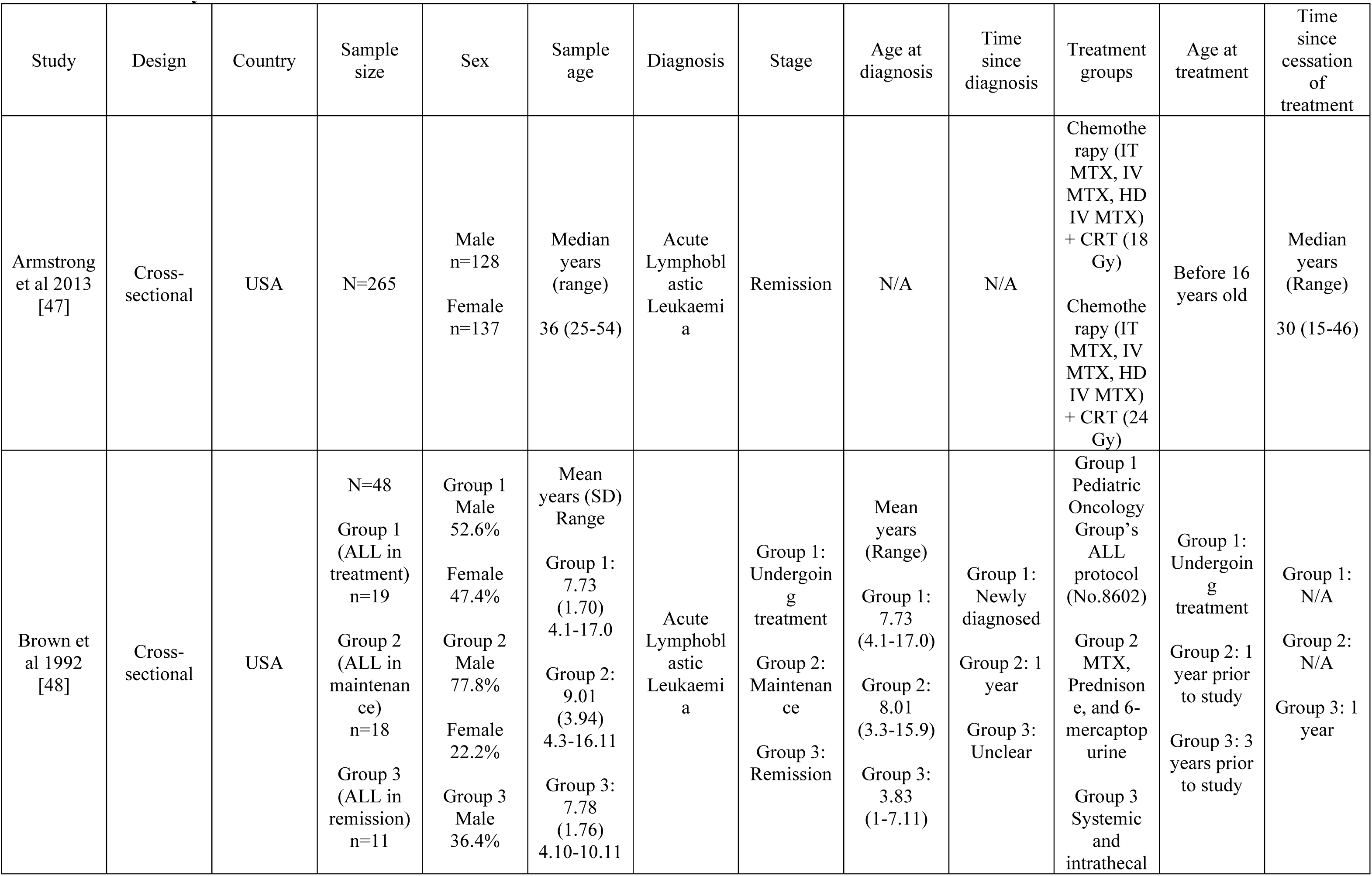
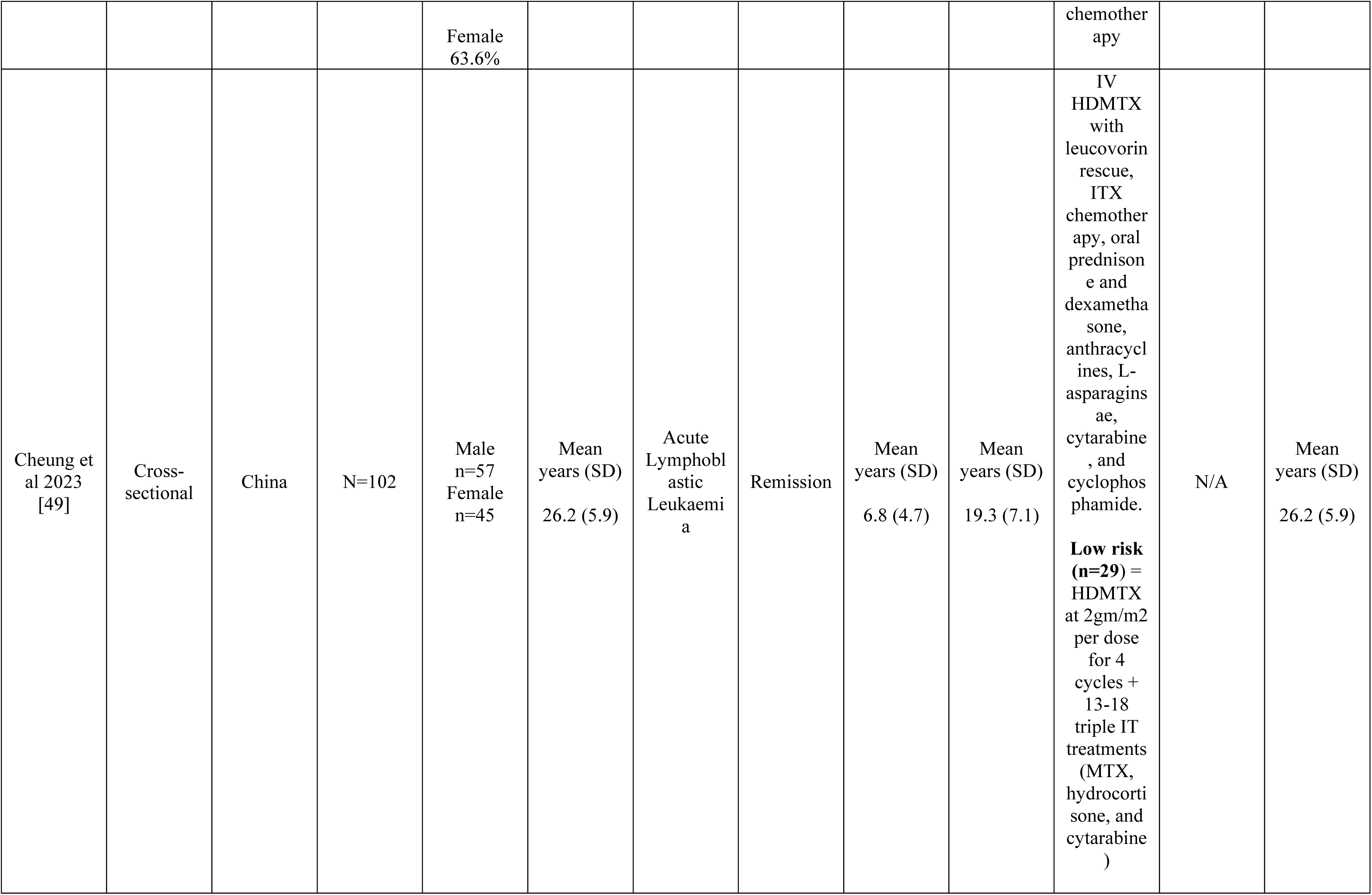
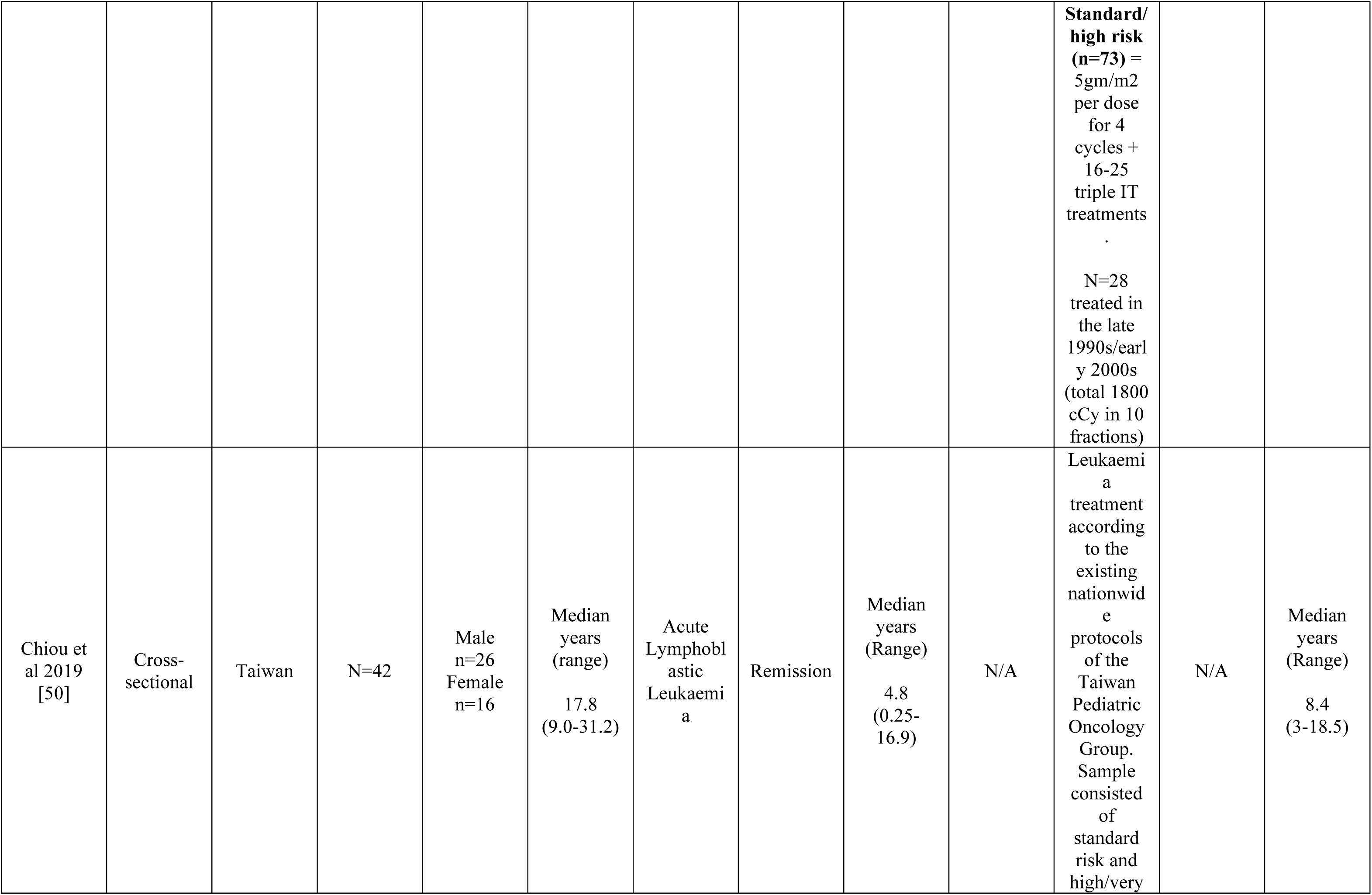
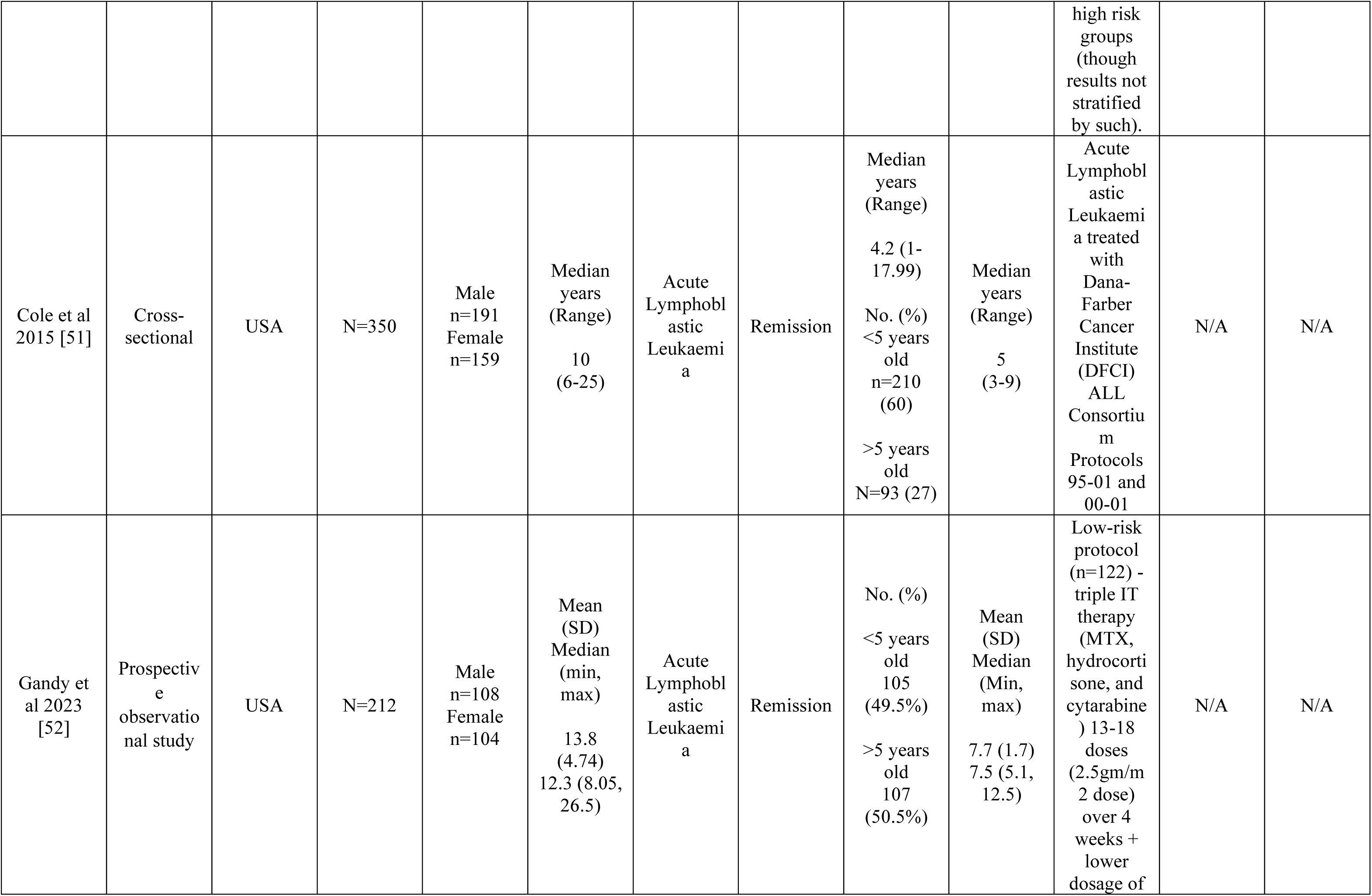
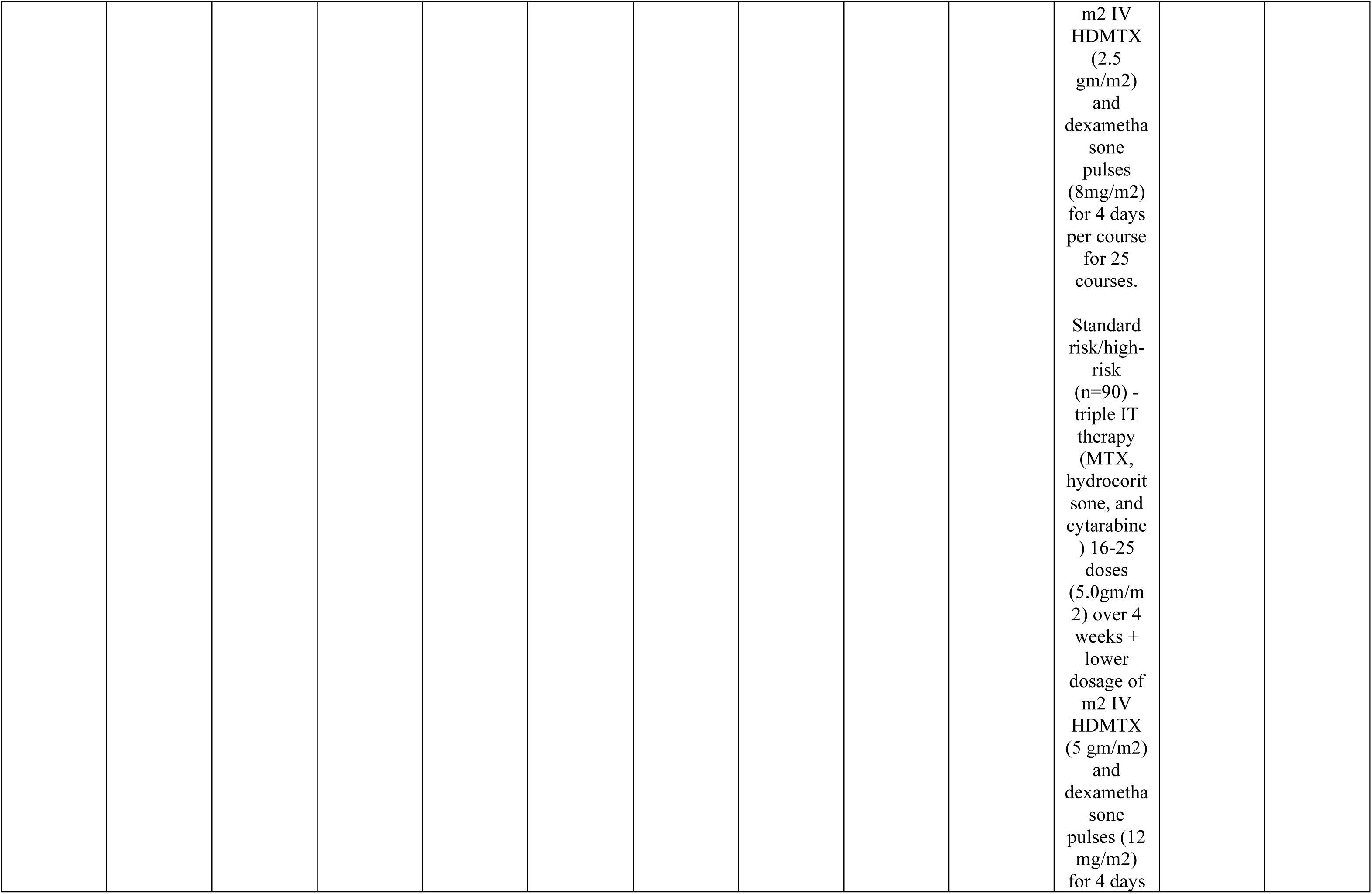
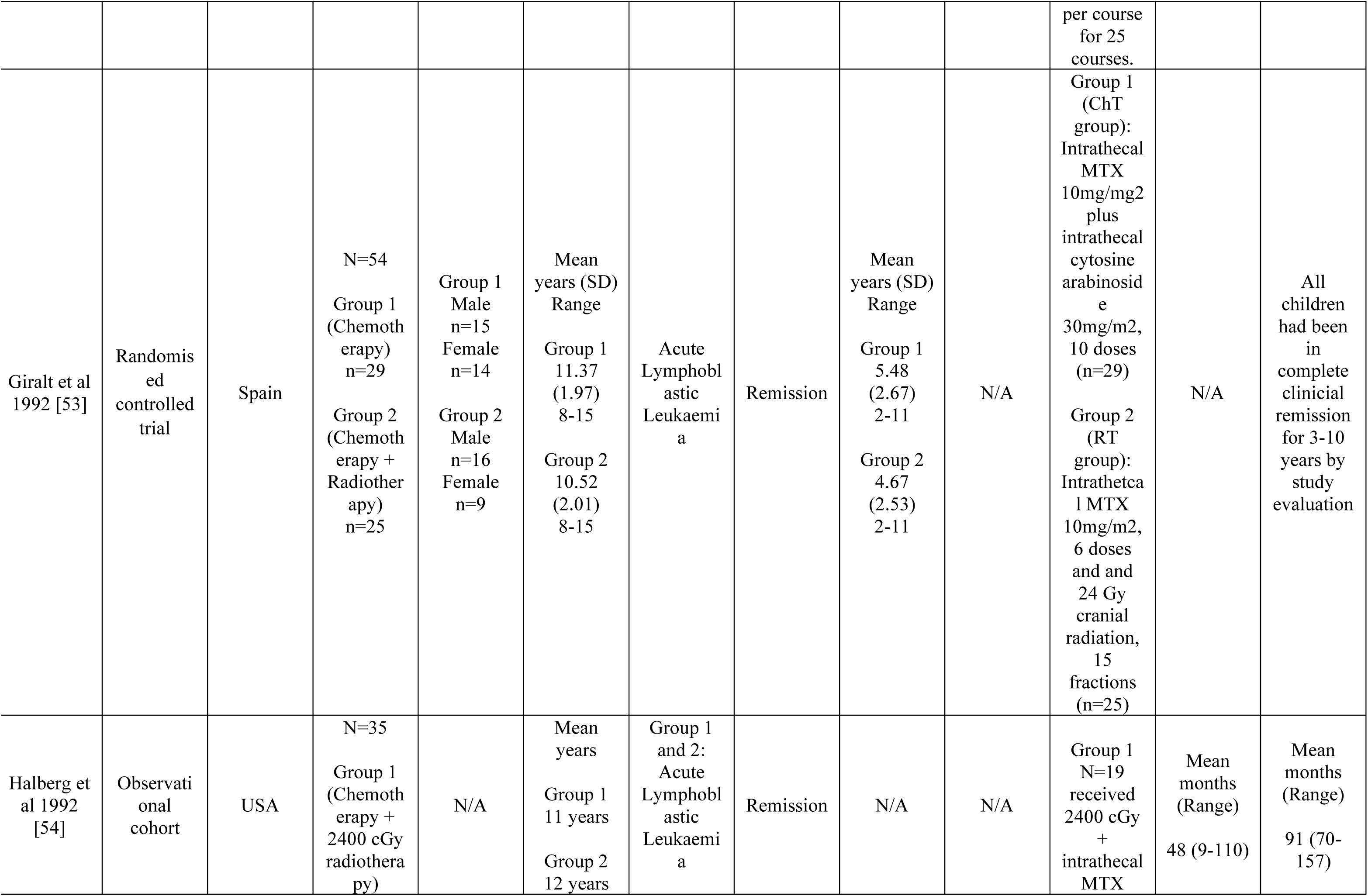
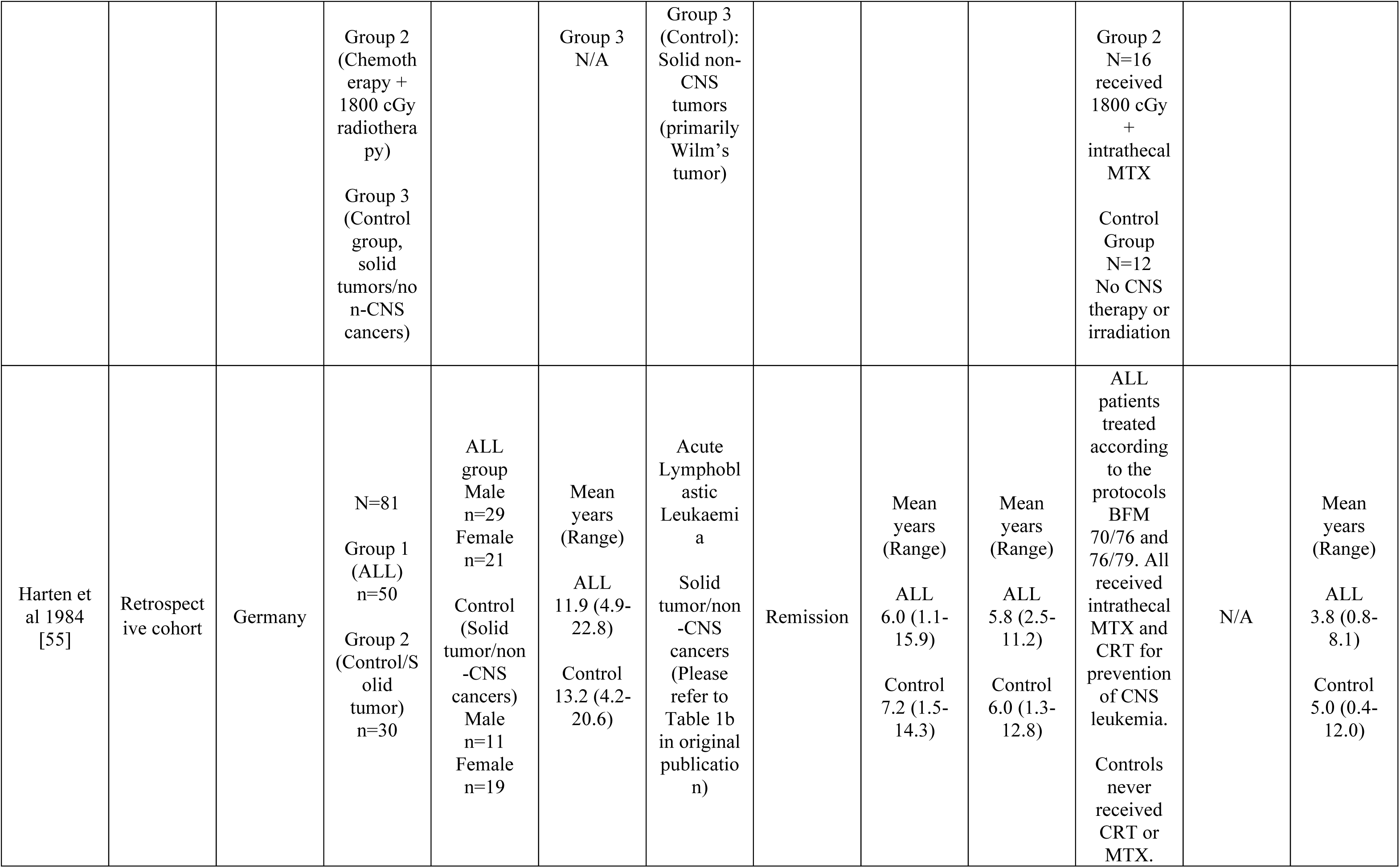
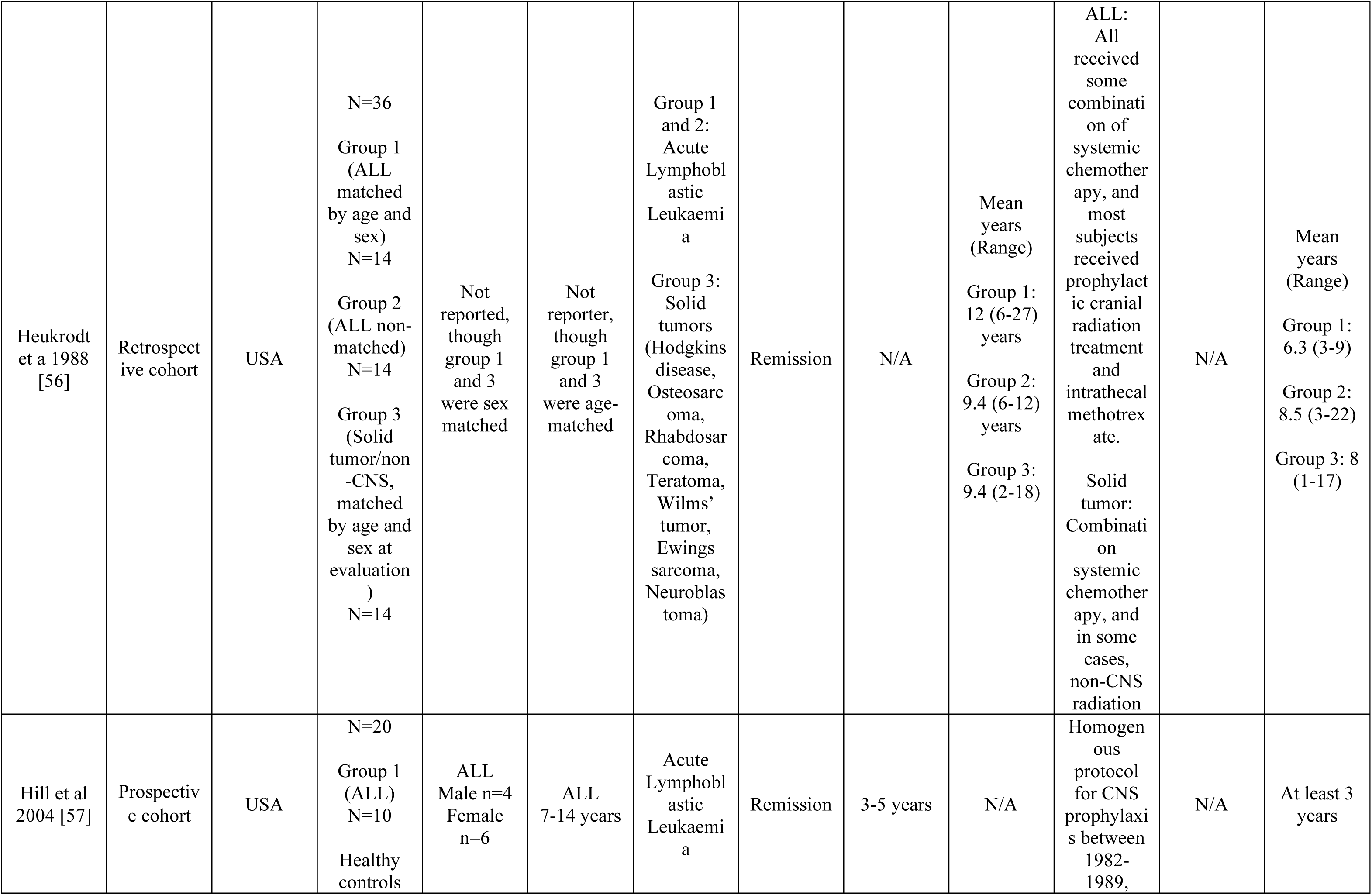
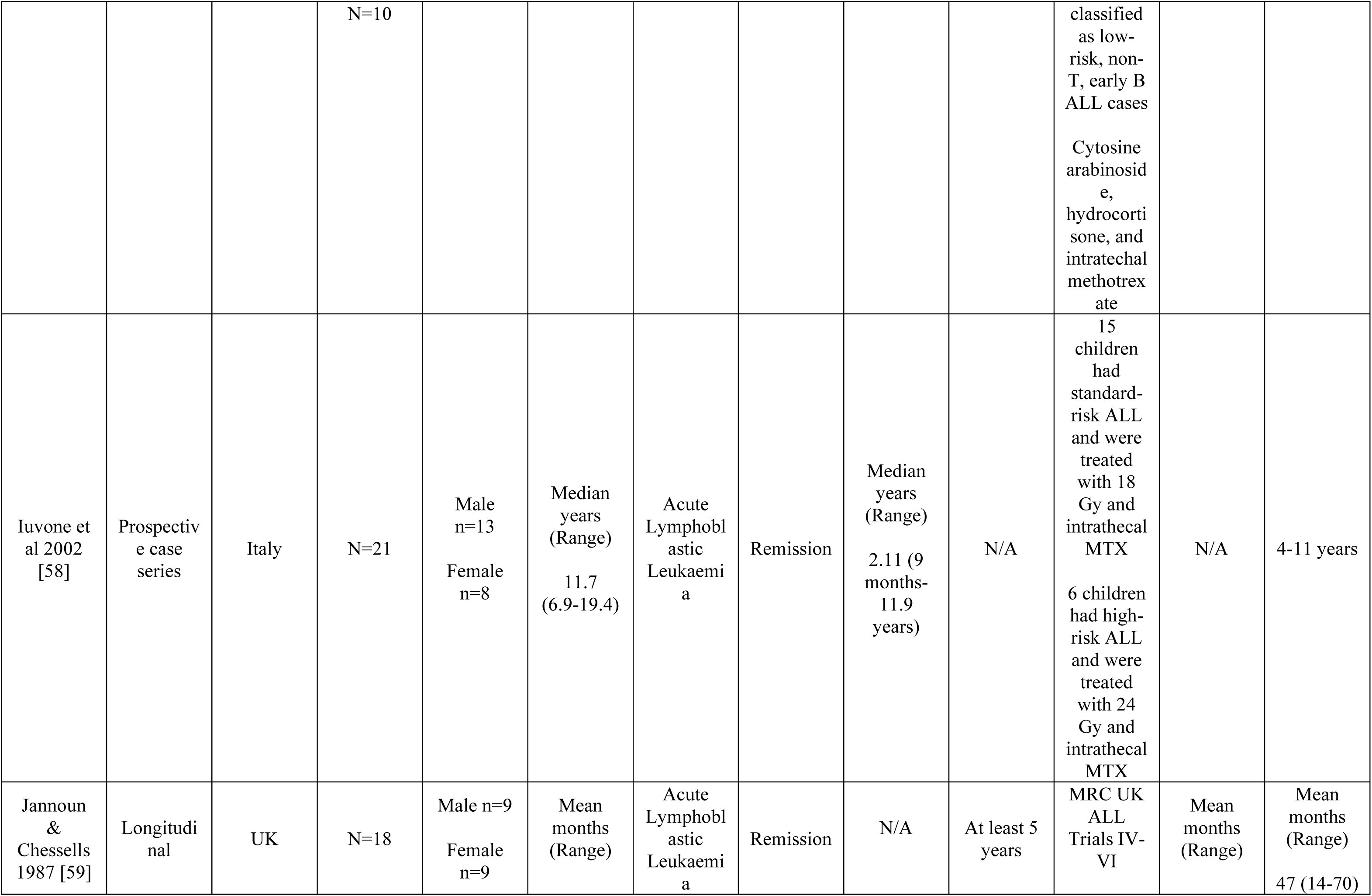
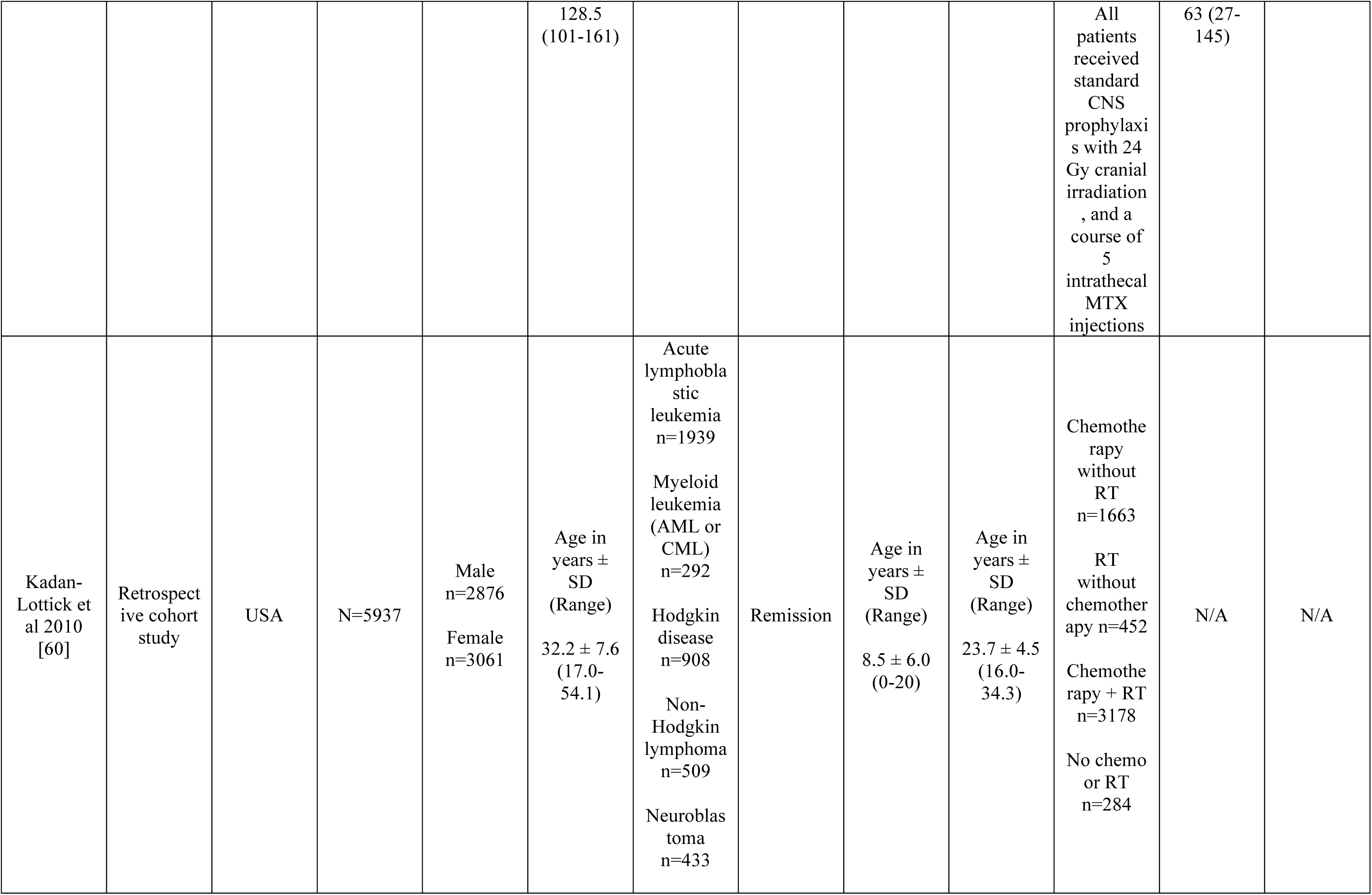
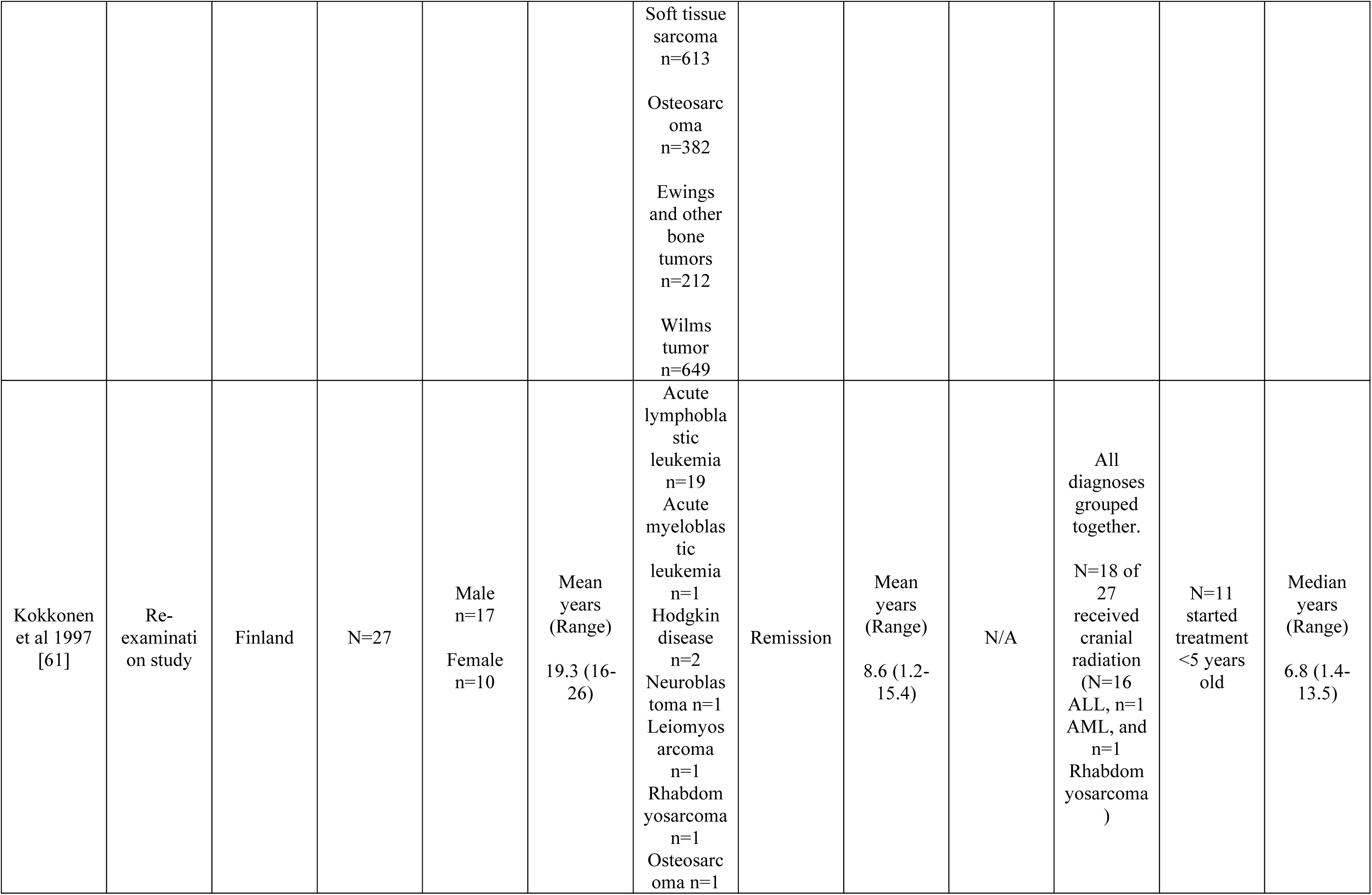
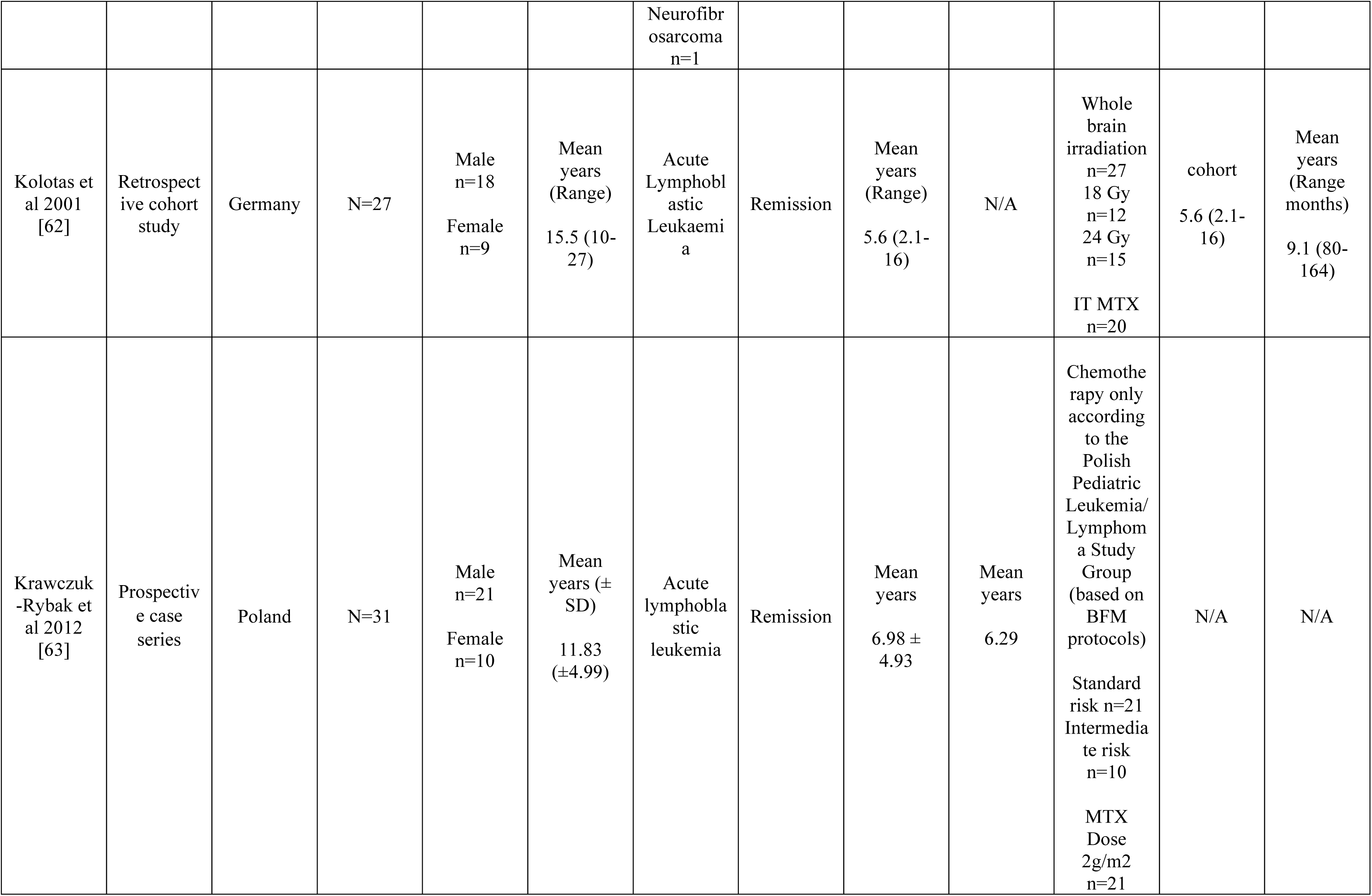
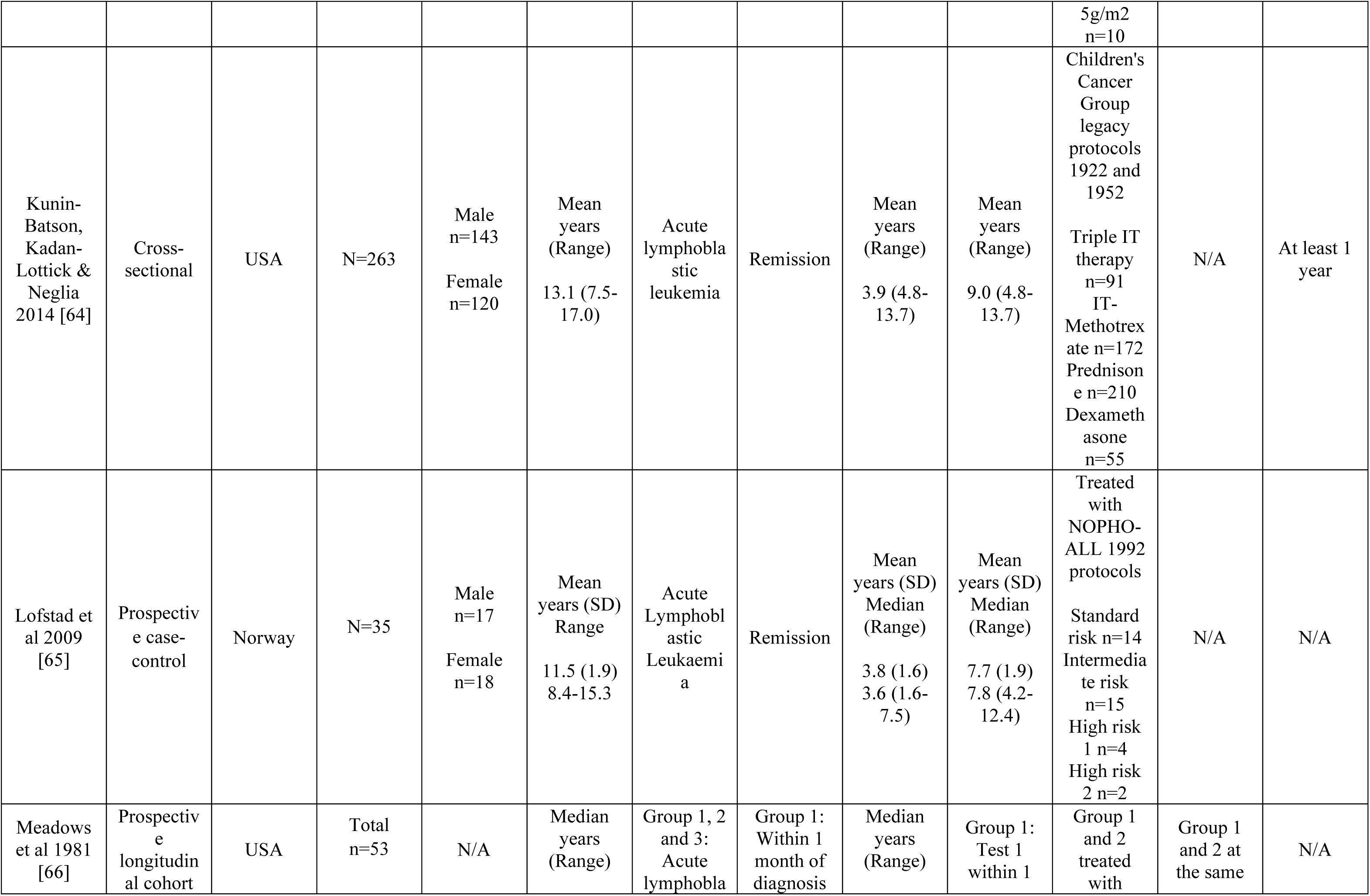
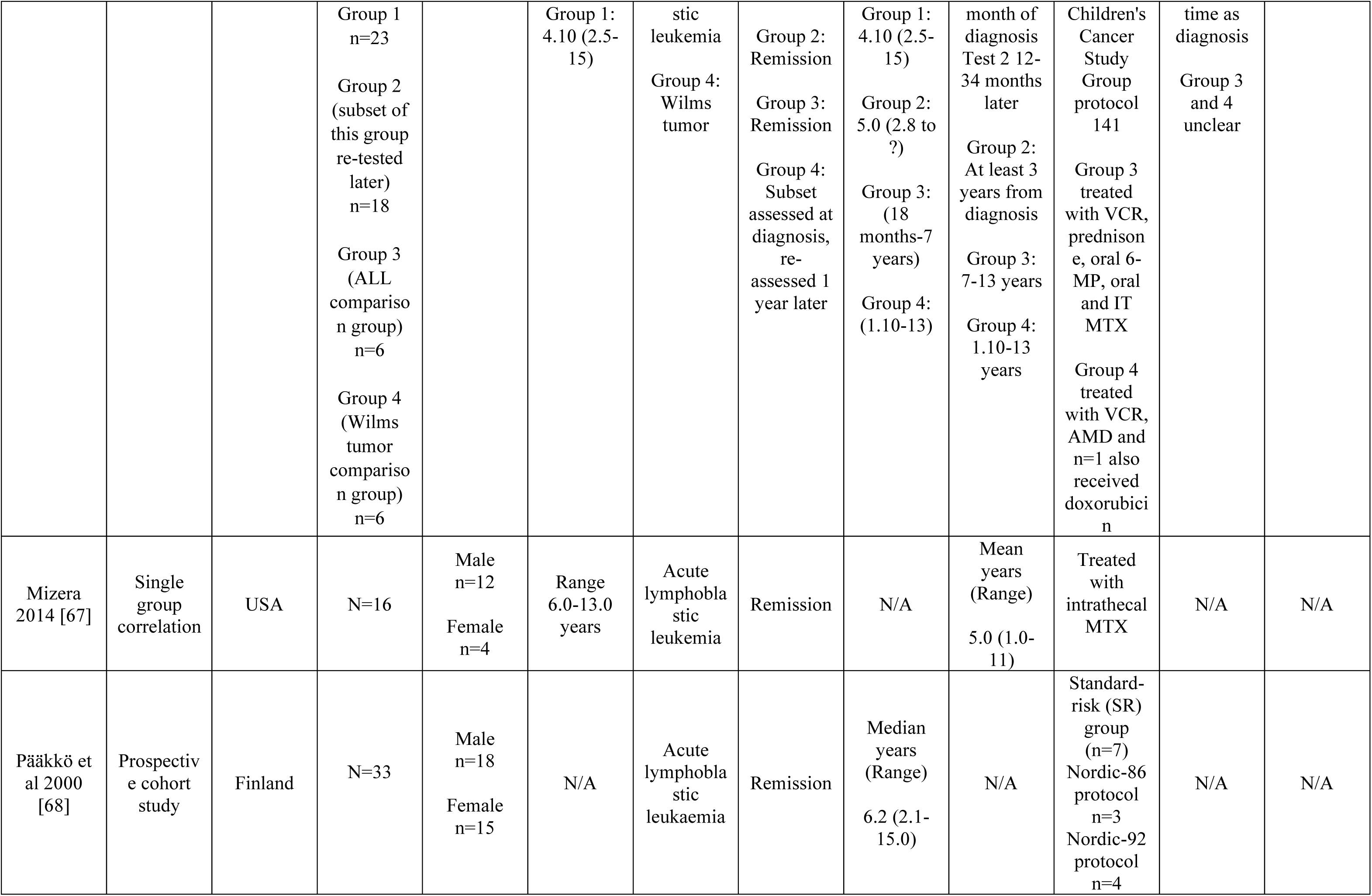
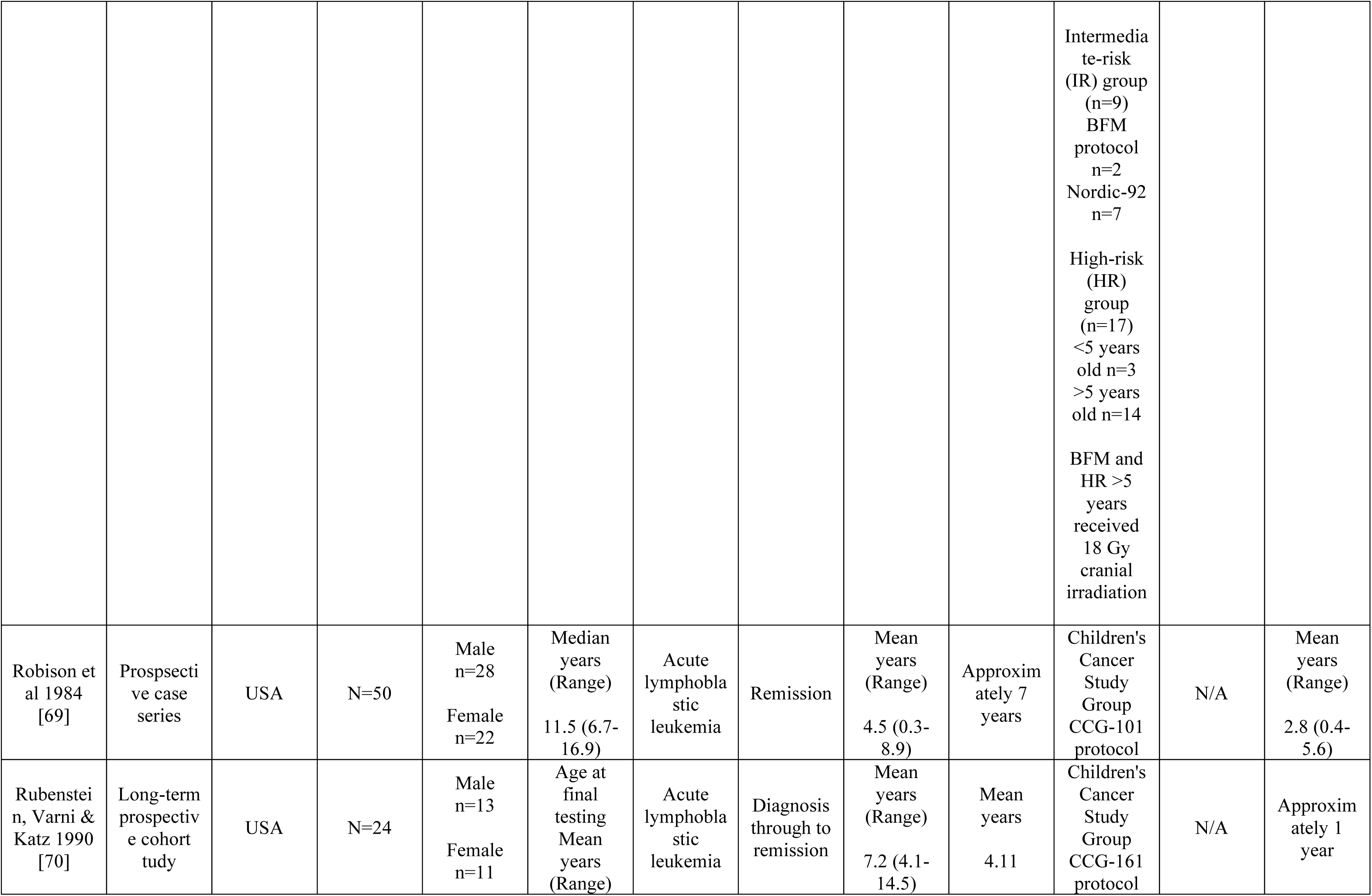
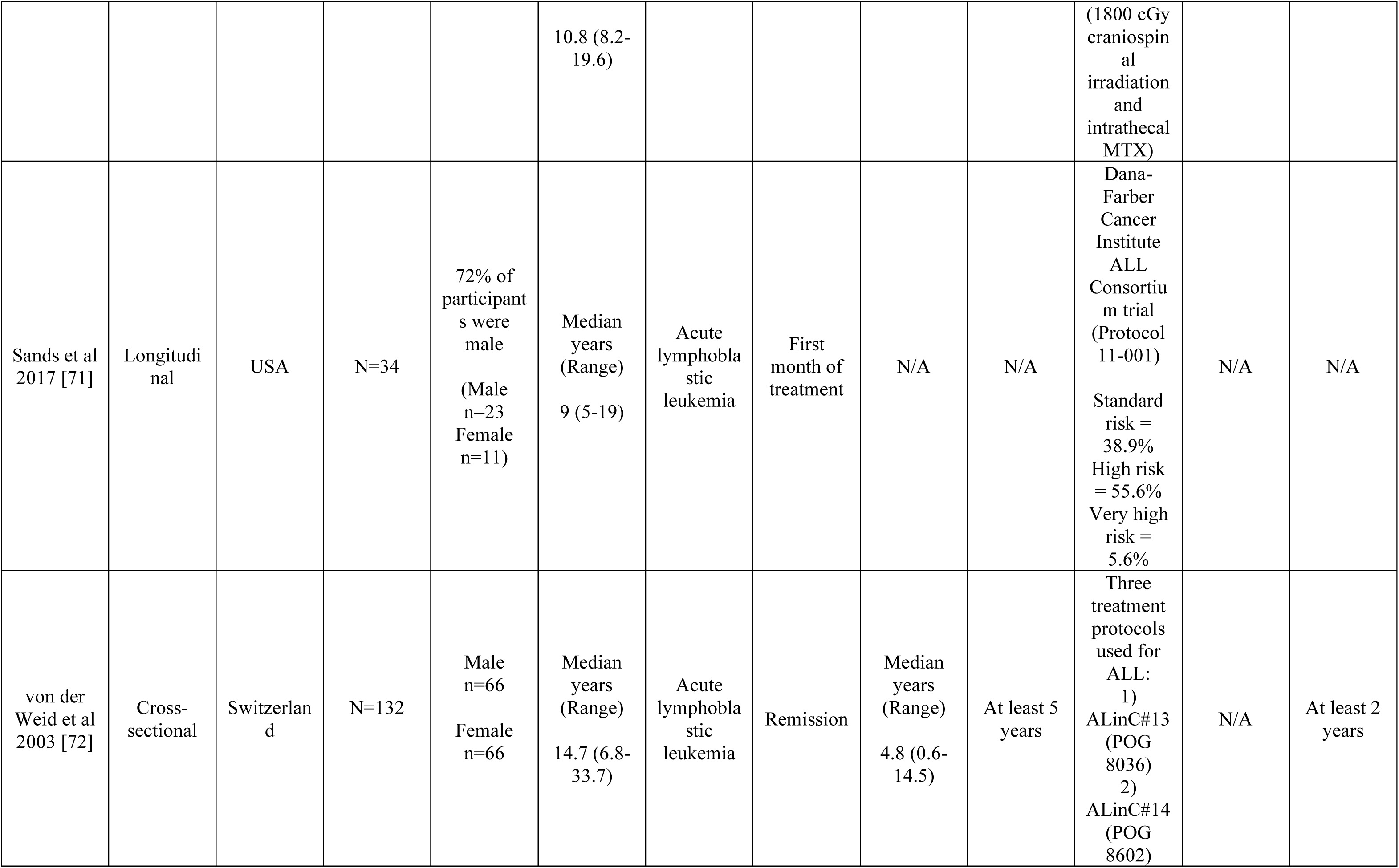
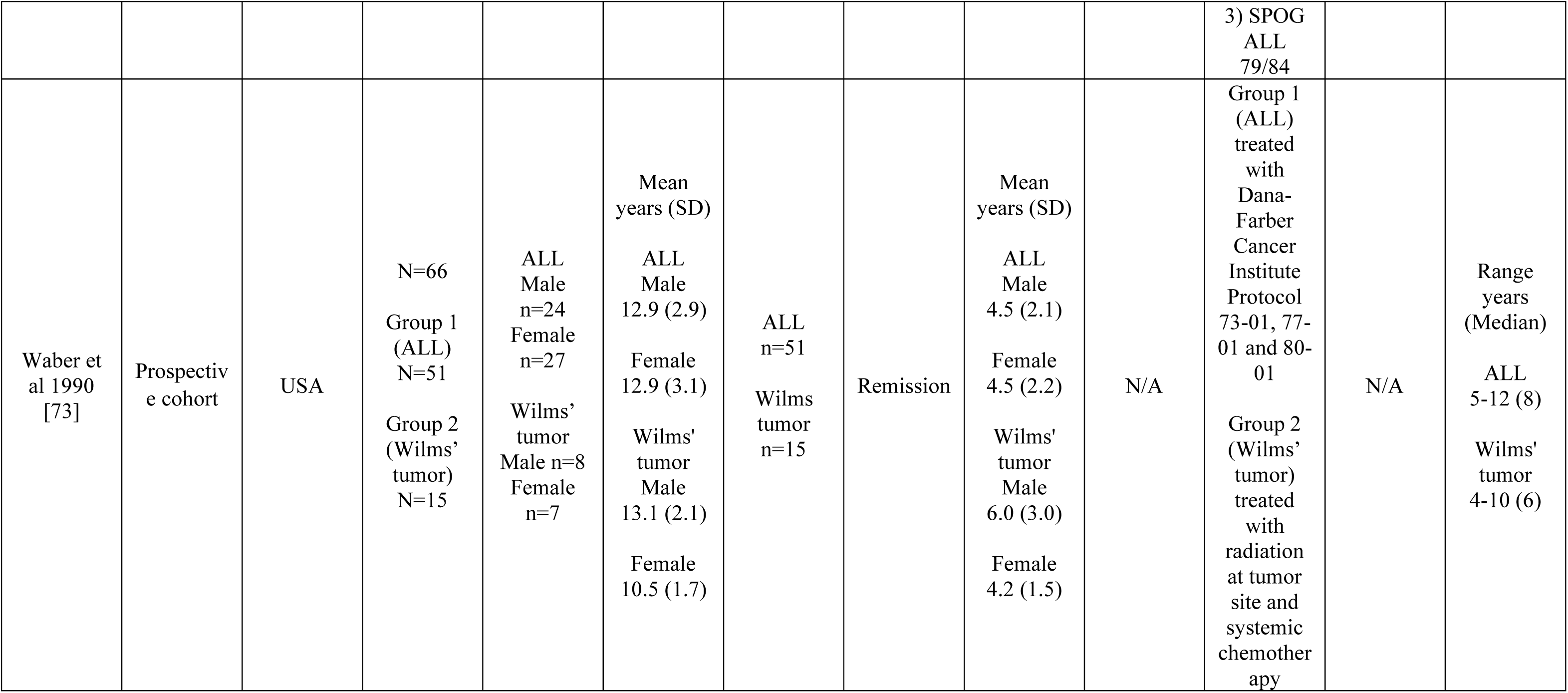
Study characteristics.

Participant characteristics extracted from the included studies comprised: age at evaluation (range: 2.5-54 years, not reported in *n* = 2), age at diagnosis (range: 0.25-20 years, not reported in *n* = 6), age at treatment (range: 9 months-16 years, not reported in *n* = 19), stage of treatment (*n* = 23 in remission, *n* = 3 followed groups of participants from diagnosis through to remission, and *n* = 1 assessed participants within the first month of treatment), treatment details **(Table 2)**, time since diagnosis (range: newly diagnosed to 34.3 years, not reported in *n* = 11), and time since cessation of treatment (range: 0.4-46 years, not reported in *n* = 9). Medical diagnoses included leukemias (acute lymphoblastic leukemia, AML or CML), Wilm’s tumor, Hodgkins disease, non-Hodgkin’s lymphoma, Osteosarcoma, Rhabdosarcoma, Teratoma, Ewings Sarcoma, Neuroblastoma, Histiocytosis, PNET, Germ cell tumor, Retinoblastoma, soft tissue Sarcoma outside head/neck, Leiomyosarcoma, Neurofibrosarcoma, and “miscellaneous”. No participants had CNS involvement of disease or CNS relapses.

Participants received a range of treatment modalities, including intravenous (IV)/intrathecal (IT) chemotherapy, radiotherapy (non-CNS and CNS-directed), or surgery. Often combination therapy was employed (e.g., chemoradiotherapy, IV + IT chemotherapy, chemotherapy + surgery). Commonly employed chemotherapy agents included methotrexate (MTX), 6-mercaptopurine, cytarabine, cyclophosphamide, anthracyclines, L-asparaginase and cytosine arabinoside. MTX was the most commonly employed chemotherapy regimen, in *n* = 25 studies. Within studies, different treatment protocols, typically pertaining to dose and dosing schedule, were employed for low risk, standard risk, high risk or very high-risk patients in order to prevent CNS involvement, with higher-risk patients receiving more aggressive treatment. Supporting treatment was employed in some cases, such as prednisone, dexamethasone and hydrocortisone. Radiotherapy was commonly employed for CNS prophylaxis or at non-CNS tumor sites, at either 18 Gy or 24 Gy doses. Treatment details are reported in full **(Table 2)**.

A wide variety of methods for assessment of cognitive impairment were employed. It was acknowledged that differences in assessment could introduce heterogeneity when assimilating prevalence estimates. Overall, 40 distinct measures were employed to assess cognition, including the same test in different editions, revised versions, and language adaptations (e.g., WISC, WISC-R, WISC-IV). All 26 studies utilized objective cognitive screening tools/cognitive batteries. It is worth noting that whilst studies employing self-report measures were within the inclusion criteria, none were eligible per other criteria. This is in contrast with the adult CRCI literature where self-report is commonly employed [26]. Detailed characteristics of the cognitive tests are reported **(Online Resource 5).**

## Methodological quality

The methodological quality of included studies is reported in **Online Resource 6**. The main domains of concern were adequate sample size (only 26.92% had adequate sample size determined either by reported power calculations or a sample size of *n* =50 per Dijkshoorn et al) [39], whether the condition was measured in a standard, reliable way for all participants (23.07% of studies satisfied this criteria), and whether appropriate statistical analysis was performed (only 15.38% of studies satisfied this criteria). Studies often neglected reporting key details surrounding who administered testing (qualifications, number of observers), order of testing, if multiple cognitive tests were administered, and the impact of extraneous factors that may have influenced results, such as fatigue. Whilst statistical analysis and methods were often well-reported, there was a distinct lack of reporting CIs.

### Prevalence estimates based on global cognitive function and intelligence

Two cognitive domain groupings arose from the objective cognitive measures: (1) global cognitive function and intelligence, and (2) achievement, aptitude and school readiness. A total of 24 studies contributed to the first grouping, with 209 prevalence estimates **(Online Resource 7)**. Of note, one study fit within both groupings, as the cognitive impairment index reported was a culmination of IQ outcomes and achievement measures; however, the prevalence estimates it provided (2 prevalence estimates) and demographic details were only considered within the “achievement, aptitude and school readiness” domain [73].

#### Non-pooled prevalence estimates of global cognitive function and intelligence

For this cognitive domain, sample sizes ranged from 16-350 [51, 67], with a mean sample size of *n =* 81 across studies.

Across studies included within this domain, there was some overlap in the cognitive measures administered and/or outcomes reported between studies, namely for IQ measures, the Wide Range Assessment of Memory and Learning (WRAML), Conners’ Continuous Performance Task (CPT) and the Rey-Osterrieth Complex Figure task. Measures of IQ were the most employed type of assessment (20 of 24 studies), with studies typically employing a form of the Wechsler Intelligence Scale with differences in Editions, age-appropriate versions, and/or language appropriate adaptations between studies. In some studies, alternative tests, such as the Stanford-Binet Intelligence Scale (Form L-M), were utilised. The primary assessed outcomes included fullscale IQ (FSIQ), verbal IQ (VIQ) and performance IQ (PIQ).

In 10 studies, outcomes were classified into severity scores (e.g., average or above, low average, borderline, and extremely low) [47, 53, 55, 57, 58, 66, 69, 71, 72], with 7 of these studies measuring IQ outcomes. The remaining three examined outcomes from the WRAML, Peabody Picture Vocabulary Test, Halstead Category Test, Ravens Progressive Matrices, McCarthy Scales of Children’s Abilities and the Cogstate battery [57, 66, 71]. For the purpose of assimilation, all scores below “average” were aggregated to provide prevalence estimates for meta-analyses where possible. Subgroup analysis was conducted to assess if between group differences were present between the outcome severity classifications.

Notably, there were differences between studies in the criteria used to define cognitive impairment. Generally, impairment was defined by comparing scores to an age-adjusted norm/T-score. Four studies employed >1 SD below the norm [47, 51, 59, 66], one study employed >1.5 SD below the norm [49] and one study employed >2 SD below the norm [66] as the cutoff score for cognitive impairment. One study explored scores <1 SD, <1.5 SD and <2 SD below the population norm for each of the reported outcomes [71]. Four studies employed less than or equal to percentile ranks (one: <5^th^, two: < or equal to 25^th^, one: < 20^th^, and one: <10^th^) as their definition of cognitive impairment [50, 51, 56, 59]. Thirteen studies defined cognitive impairment according to the relevant test criteria (e.g., IQ <80) [52–55, 57, 58, 62–65, 67, 69, 72]. The criteria for defining cognitive impairment were unclear for the remaining studies [61, 68, 70].

One study additionally assessed cognitive outcomes across different stages of treatment. Rubenstein et al assessed the same patients across two time-points – at diagnosis prior to beginning treatment and at 4–5-year follow-up after treatment [70]. Patients did not display any signs of cognitive impairment when newly diagnosed, evidenced by all falling into the normal IQ range. Over time, however, Rubenstein et al. demonstrated a decline in IQ within the same patients from newly diagnosed to 4-5 years post-treatment, with 12.5% of patients scoring low-average and 8.3% scoring borderline, dropping an average of 23 IQ points [70].

Non-pooled prevalence estimates for global cognitive function and intelligence ranged from 0-67%.

#### Meta-analysis of global cognitive function and intelligence

Meta-analysis was appropriate for further investigation of prevalence estimates from studies that employed objective cognitive test measures. A total of 18 studies contributed prevalence data for meta-analysis, contributing 36 data points over 12 meta-analyses [49–55, 58, 59, 61–65, 67, 69, 70, 72]. Five studies, with six data points, were excluded from meta-analysis, as there were not sufficient sample sizes within these studies to justify inclusion (WRAML verbal memory recall and visual memory recall, CPT detectability and Rey-Osterrieth Complex figure, n=1-2 per outcome) [50, 52, 57, 59, 64].

The overall random-effects pooled prevalence in percentage (with 95% CIs; and 95% PIs) of cognitive impairment is reported as follows - 15.02% in fullscale IQ (CIs 11.39-19.53; PIs 5.37-35.47) **(Figure 2)**, 18.05% in verbal IQ (CIs 9.62-31.30; PIs 1.37-77.78) **(Figure 3)**, and 12.46% in performance IQ (CIs 8.12-18.64; PIs 3.33-37.06) **(Figure 4)**. The overall fixed-effects pooled prevalence in percentage (with 95% CIs) of cognitive impairment is as follows - 9.83% in CPT-II omissions (CIs 7.58-12.67) **(Figure 5A)**, 12.05% in CPT-II commissions (CIs 9.65-14.94) **(Figure 5B)**, and 10.14% in CPT-II variability (CIs 7.80-13.09) **(Figure 5C)**. Subgroup analysis was conducted for cancer type (ALL vs other non-CNS cancers), CNS vs non-CNS directed treatment, time since cessation of treatment, and outcome severity, where data were available.

**Figure 2.**
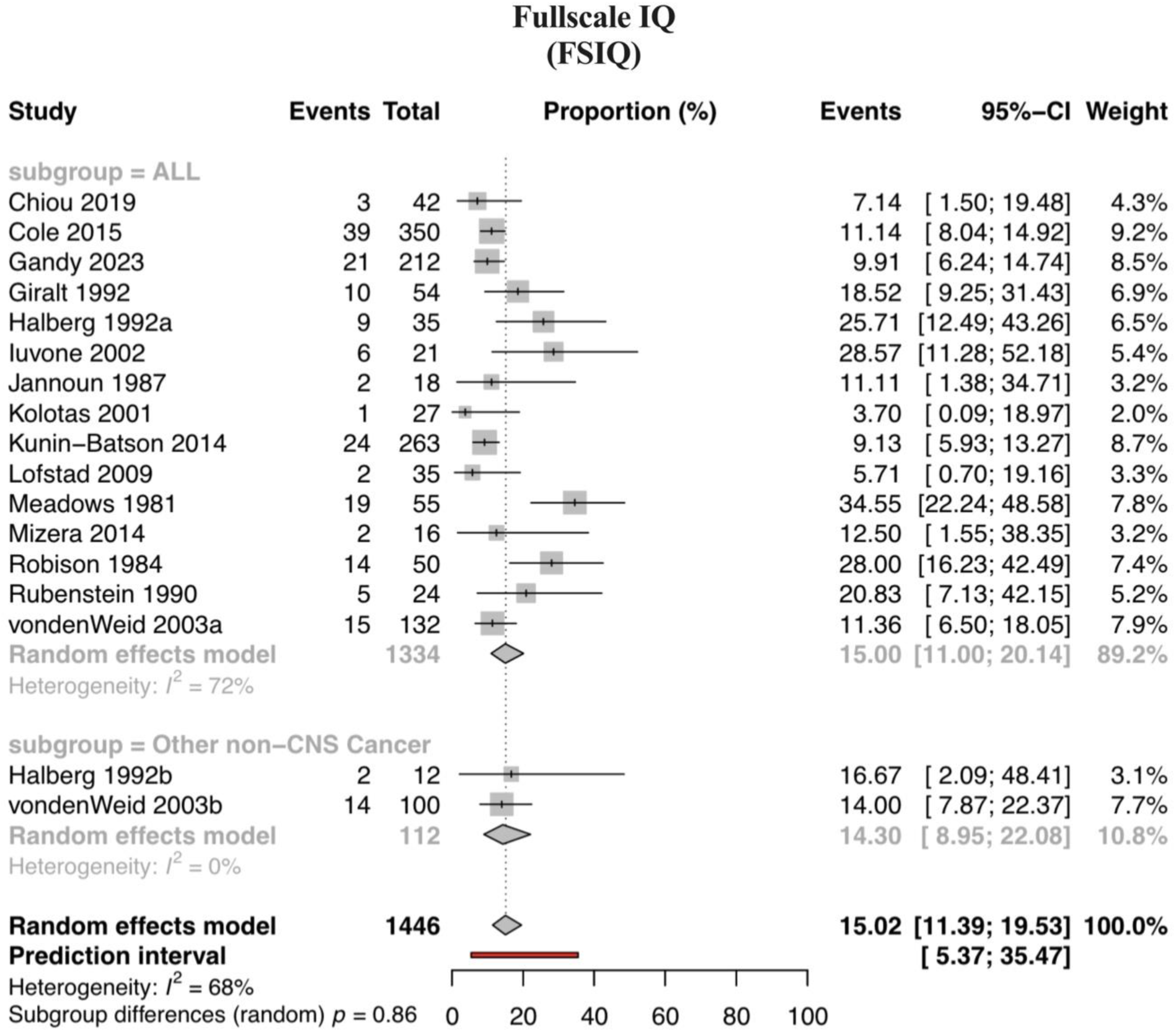
Pooled prevalence estimates (random effects model) of fullscale IQ with subgroup analysis of cancer type (ALL and other non-CNS cancers) (no significance).

**Figure 3.**
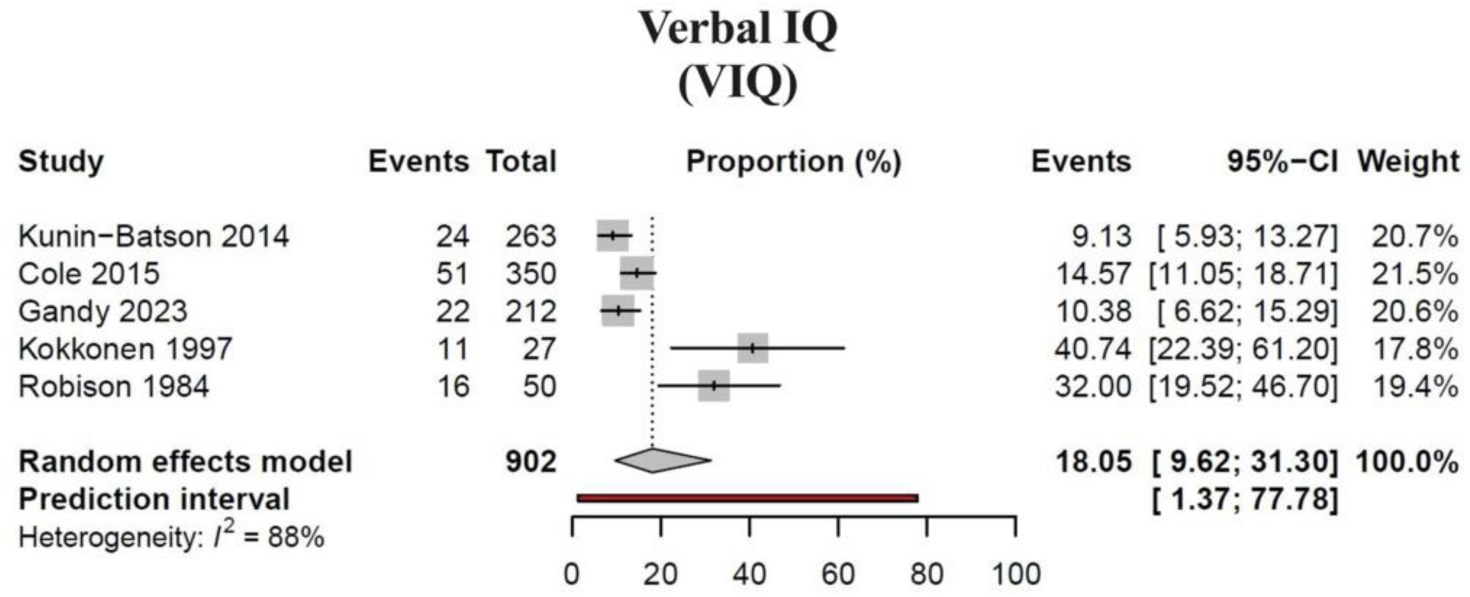
Pooled prevalence estimates (random effects model) of verbal IQ.

**Figure 4.**
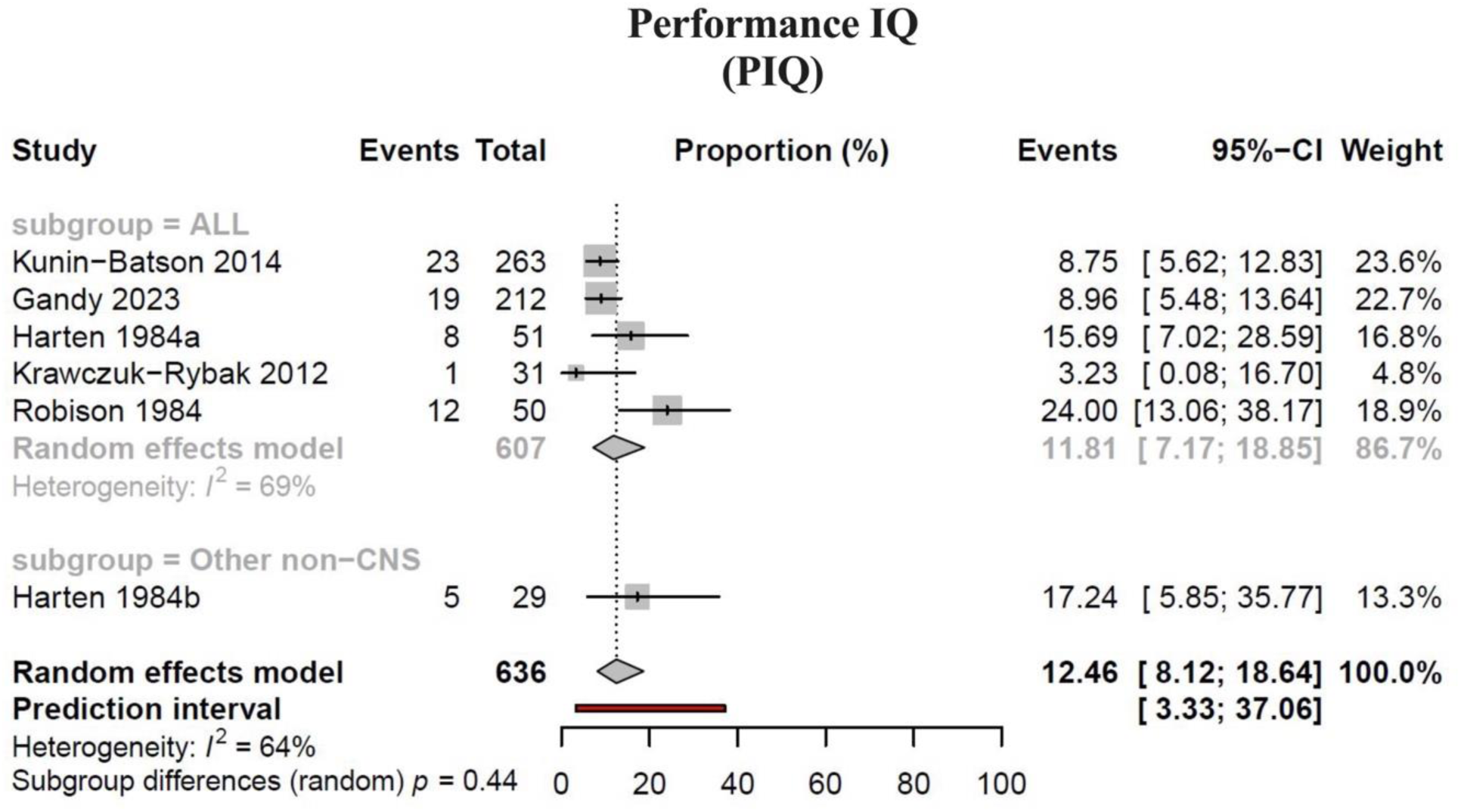
Pooled prevalence estimates (random effects model) of performance IQ with subgroup analysis of cancer type (ALL and other non-CNS cancers) (no significance).

**Figure 5.**
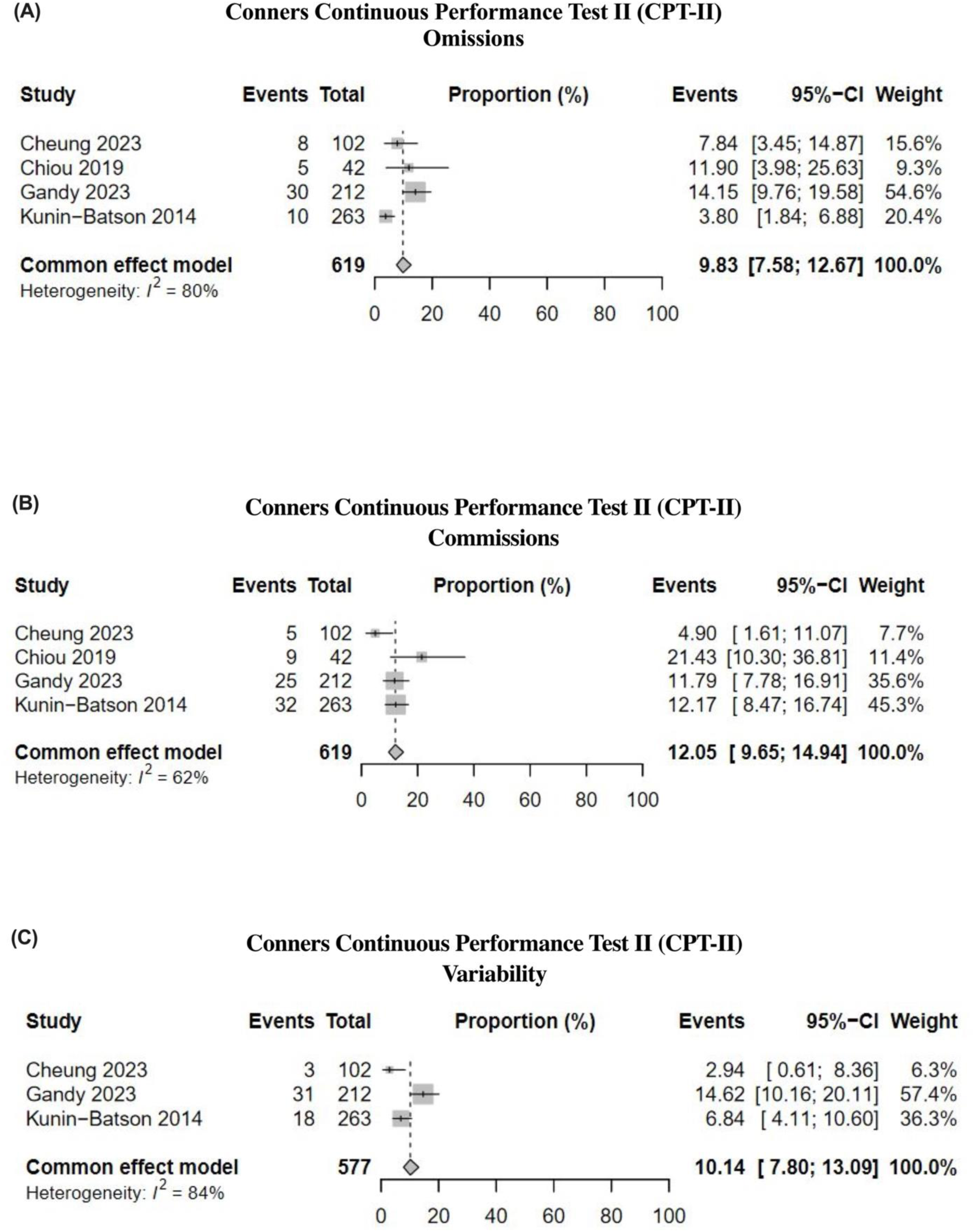
*(A)* Pooled prevalence estimates (common effect model) of CPT-II Omissions, *(B)* Pooled prevalence estimates (common effect model) of CPT-II Commissions, and *(C)* Pooled prevalence estimates (common effect model) of CPT-II Variability.

#### Subgroup analysis of global cognitive function and intelligence

There were no significant subgroup differences for cancer type, CNS vs non-CNS directed treatment, or time since cessation of treatment in either FSIQ or PIQ measures **(Online Resource 8, Supplementary figures 1-6)**. Random effects models were employed for all subgroup analyses, sans time since cessation of treatment for PIQ, which employed a fixed effects model due to the inclusion of <5 studies. There was a significant difference in outcome severity in FSIQ (X^2^(2) = 9.54, *p < 0.01*), with participants primarily falling into the “borderline” (13.04%; CIs 6.35-25.20) or “low average” (13.00%; CIs 7.03-22.78]) classification, rather than “extremely low” (3.61%; CIs 1.81-7.07). The overall prevalence estimate for FSIQ impairment reported with outcome severity classification was 8.54% (Cis 4.98-14.24; PIs 1.58-35.13) **(Figure 6A)**. Similarly, there was a significant difference in outcome severity in PIQ (X^2^(1) = 10.59, *p < 0.01*), with participants primarily falling in the “low average-borderline” (17.07%; CIs 11.51-24.58) classification, rather than “extremely low” (3.22%; CIs 1.21-8.27). The overall prevalence for PIQ impairment reported with outcome severity classification was 9.65% (CIs 4.78-18.52; PIs 1.14-49.78) **(Figure 6B)**.

**Figure 6.**
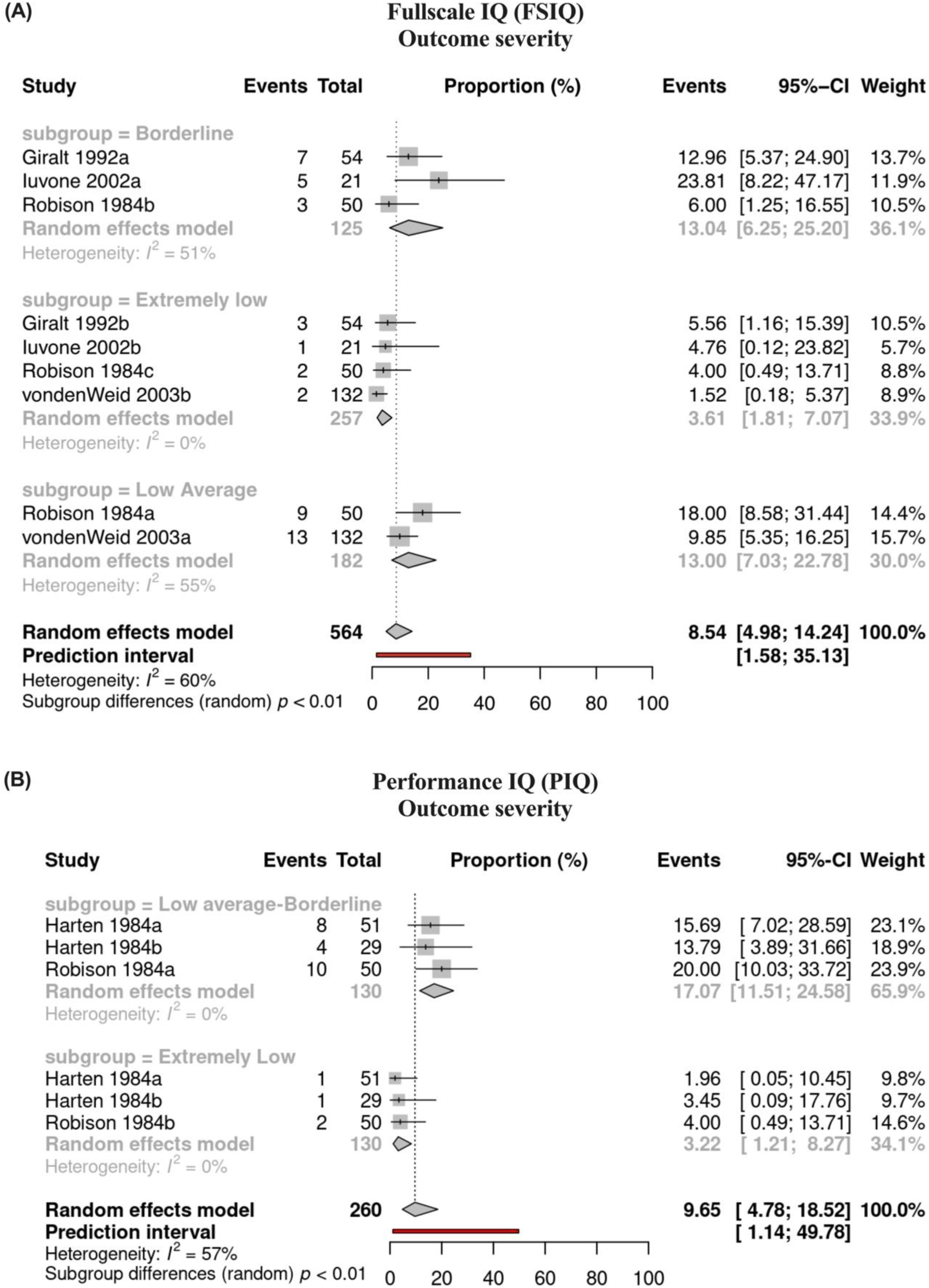
*(A)* Pooled prevalence estimates (random effects model) of fullscale IQ with subgroup analysis of outcome severity (borderline, low average, and extremely low). Significant differences with participants primarily falling in borderline and low average, compared to extremely low (X^2^(2) = 9.54, p < 0.01), and *(B)* Pooled prevalence estimates (random effects model) of performance IQ with subgroup analysis of outcome severity (low average-borderline and extremely low). Significant differences with participants primarily falling in the low average-borderline range compared to extremely low (X^2^(1) = 10.59, p < 0.01).

### Prevalence estimates based on achievement, aptitude and school readiness

A total of four studies contributed to the achievement, aptitude and school readiness domain, with 6 prevalence estimates **(Online Resource 9)** [48, 59, 64, 73]. Each study employed validated objective measures of cognition, including the Detroit Test of Learning Aptitude, BAS Word Reading Test, Wechsler Individual Achievement Test 2^nd^ Edition Abbreviated, and a combination of the Wechsler Intelligence Scale Revised and the Wide Range Achievement Test Revised (culminating in a “cognitive impairment index” score). There was no overlap in the tests administered between studies.

Sample sizes ranged from 18-263 [59, 64], with a mean sample size of *n =* 99 across studies. One study assessed cognitive outcomes across different stages of treatment [48]. Brown et al assessed three separate groups of patients that were either newly diagnosed, 1 year into treatment, or 1-year post-treatment [48]. Patients did not display any signs of cognitive impairment when newly diagnosed, as evidenced by 0% learning disabilities within the cohort. In contrast, patients 1 year into treatment and 1-year post-treatment demonstrated greater impairment, as evidenced through an increase in the number of diagnosed learning disabilities (15.4% and 60%, respectively), although it is important to note that these were not the same patients followed across time [48].

As noted above, for global cognitive function and intelligence, there were differences between studies in the criteria used to define cognitive impairment. In one study, the definition was unclear [48]. Two studies compared scores to age-related norms, in one case defining impairment as scores below the 25^th^ percentile [59], and another with scores falling >1 SD below the population mean [64]. Waber et al employed a composite score, called the “Cognitive Impairment Index”, which incorporated both IQ and academic achievement outcomes into a 5-scale index, with 0 being the norm and 1-4 suggesting impairment of increasing severity (IQ >90, 1 SD below mean on 1-2 tests versus IQ <90, 1 SD below mean on all four tests) [73].

Non-pooled prevalence estimates for achievement, aptitude and school readiness ranged from 0-73%. Meta-analysis was not appropriate for further investigation, due to heterogeneity in assessment methods and outcomes between the small number of studies investigating this domain.

### Heterogeneity and publication bias

Heterogeneity was assessed by interpreting PIs, where available. Across the included meta-analyses, the PIs suggest that future studies conducted in this population may result in a wide and varied dispersion of effect sizes around the mean estimate effect we have calculated, in both directions (lower or higher than our estimate), reflecting heterogeneity **(Figures 3-6)**.

The doi plots to assess publication bias revealed some asymmetry, implying the presence of potential bias. There was no evidence of publication bias for FSIQ cancer type, FSIQ treatment type, PIQ treatment type, PIQ time since cessation of treatment, and CPT Commissions. Minor asymmetry was reported in FSIQ outcome severity and PIQ cancer type. Major asymmetry was reported in FSIQ time since cessation of treatment, VIQ overall, PIQ outcome severity, CPT Omissions and CPT Variability **(Online Resource 10)**. It is notable that there are no formal guidelines for assessment of publication bias when data is proportional [40]. Further, while doi plots are being promoted as a more reliable alternative to funnel plots, which are imprecise at extremes of proportion, there may be similar concerns for doi plots [40, 74, 75].

### GRADE certainty assessment and results

The evidence presented was assessed using the GRADE approach (GRADEpro GDT: GRADEpro Guideline Development Tool [Software]) in two groupings – (1) global cognitive function and intelligence and (2) achievement, aptitude and school readiness. Global cognitive function and intelligence included 11 outcomes – 4 graded as very low, 5 graded as low and 2 graded as moderate. Concerns commonly stemmed from risk of bias, inconsistency (variable prevalence) and imprecision (wide confidence intervals). However, there is a lack of formal guidance from the GRADE working group on the use of methodology for reviews of prevalence. As such, the results from this review should be interpreted with caution and conclusions should be made tentatively. Results are presented in the Summary of Findings Table **(Table 1)**.

## Discussion

To our knowledge, this is the first systematic review investigating the prevalence of CRCI in non-CNS cancers in paediatric patients with a meta-analytical assimilation. Previous publications in this area have provided foundational insights into these concerns via both a scoping review and a targeted review encouraging a call to action from the European Cancer and Cognition Consortium (ECCC) [76, 77]. Our primary findings highlight evidence of impairments across the two cognitive domains assessed - (1) global cognitive function and intelligence, and (2) achievement, aptitude and school readiness. Notably, pooled prevalence estimates suggest that survivors of non-CNS cancers experience impairments in measures of intelligence (12.46%-18.05%) and attention (9.83%-12.05%). Non-pooled prevalence estimates ranged from 0-67% in global cognitive function and intelligence, and 0-73% in achievement, aptitude and school readiness. Whilst highly variable, this is comparable to reports of CRCI across other populations, e.g., a systematic review and meta-analysis of breast cancer survivors yielding 0-83% [26], and reviews reporting figures such as the commonly cited 30-75% [78], or “up to 75%”, both derived from breast cancer survivors [78, 79]. Further, interestingly, the range of cognitive impairment that we report here for non-CNS cancers does not differ substantially from that previously reported in a systematic review of pediatric CNS cancer survivors (i.e. 0-75%, with a mean of 30.3%) [80]. The current review elected to focus specifically on non-CNS cancers, in order to minimize the potential for CNS involvement and CNS diagnoses to conflate prevalence estimates, given the higher risk for developing cognitive impairment in this population due to the impact of the tumour itself, as well as direct injury to the CNS via concentrated radiation and/or chemotherapy doses [81, 82].

ALL is the most diagnosed non-CNS cancer in children; therefore, it is not surprising that the included studies primarily focused on ALL, compared to other non-CNS cancers [1]. Though no subgroup differences were revealed in the outcomes measured between ALL and other non-CNS cancers, there was a significant lack of studies that either assessed cancers other than ALL, or clearly delineated the data from ALL and non-ALL participants to allow for comparison between cancer types. Of those that did present the latter, the sample sizes were exceptionally small in the meta-analysis subgroup analyses compared to ALL populations. Overall, non-CNS cancers other than ALL were not well represented and need to be investigated further.

Regardless of cancer type, whilst investigating outcome severity of intelligence measures, most participants fell within the milder impairment category, rather than the severely impaired category, aligning with our theory that there is impairment seen in pediatric cancer patients, but that its presentation may be more subtle rather than pervasive [83, 84]. Interestingly, a majority of the included studies employed measures of IQ. IQ tests measure both fluid intelligence (g_f_), reflecting the ability to navigate and solve new problems with no prior learning or experiences to call on, and crystallised intelligence (g_c_), reflecting the knowledge we accumulate through life, learning, and experiences [85]. IQ subtests further investigate verbal IQ and performance IQ, reflecting verbal skills versus visual, speed, and perceptual abilities, respectively, as well as full scale IQ (combining verbal and performance IQ scores) [86]. Whilst IQ performance has been commonly investigated and is a standard approach for identifying difficulties in childhood cancer survivors (such as learning disabilities), it offers a limited view of CRCI, failing to capture performance in specific cognitive domains, such as memory, processing speed and executive function [36]. Neuropsychological testing, which can be used to evaluate domain-specific impairments, would provide a more comprehensive assessment than IQ measures alone can capture, and might be better able to capture subtle changes in function [36]. Even with the use of neuropsychological testing, it is worth noting that some tests may not be sensitive enough to pick up on the potentially more subtle changes that occur in the context of CRCI, particularly when considering that the pediatric population is currently underrepresented within existing literature [83, 87, 88]. Therefore, more sensitive neuropsychological measures, validated for use in pediatric cohorts, may be needed to truly assess the presentation of cognitive change within this population. Further, objective assessments such as those used in the included studies may fail to account for individuals who still score within the “average” range, but who have experienced a significant drop in performance (e.g., IQ points, lower attentional function) compared to their pre-cancer baseline, with such changes still resulting in an impact on quality of life and functional performance for the individual [87–89]. In order to assess individual impacts, self-report measures of perceived changes in cognitive function may be needed, which, at least based on this review, appear to be less frequently employed in this population, perhaps due to their age. In line with this, despite commonly held concerns regarding the reliability and validity of self-report over objective measures [36, 90], prevalence estimates among survivors in large scale studies employing the Childhood Cancer Survivor Study Neurocognitive Questionnaire (CCS-NCQ) have demonstrated great reliability, construct validity and discriminative validity within outcome measures of memory, task efficiency and organisation validated in childhood cancer survivors specifically [60, 91, 92]. There are certainly good arguments for giving more weight to outcomes from such validated self-report assessments compared to the range of objective, but perhaps overly gross, measures employed within this review, at least whilst a gold standard neuropsychological test(s) is developed specifically for CRCI and subsequently validated in children.

It is additionally noteworthy that the achievement, aptitude and school readiness domain was not well-represented, particularly in comparison to the global cognitive function and intelligence domain, with only four studies included, each employing varied methodologies and thus providing no means to employ meta-analytical methods. This is in stark contrast to the short- and long-term quality of life impacts that have been widely demonstrated across childhood cancer survivors in the literature including, but not limited to, struggling with school reintegration, impaired academic performance, lower levels of education attainment, and higher need for specialized education and additional support, with subsequent impacts on self-esteem, confidence and mental health [19–22, 93]. Whilst changes in global cognitive function and intelligence are certainly evidenced to occur, it is limiting to focus solely on these aspects, without considering the real-life impact CRCI symptoms have on quality of life and tangible outcomes [20, 22]. Just as IQ alone offers a limited view of CRCI, failing to capture more subtle, domain-specific changes, investigating CRCI without considering tangible day-to-day outcomes further limits our understanding of its full impact.

Only two studies included in this review assessed impairment over time [48, 70]. Both demonstrated that impairments were non-existent at diagnosis, but that individuals declined both throughout and following treatment. Brown et al assessed participants longitudinally but did not assess this trajectory past 1-year post-cessation of treatment (time-points assessed: newly diagnosed, 1-year into treatment, 1-year post-treatment) [48], whilst Rubenstein examined different participants cross sectionally at diagnosis and at 4-5-year follow-up after treatment [70]. To date, no studies have looked longer than 4-5 years following treatment cessation. This is noteworthy, as time since cessation of treatment may further influence cognitive impacts; to date, however, the literature is unclear as to whether cognitive impairments improve, decline, or plateau over time following cancer treatment, even in adult populations. For example, in breast cancer populations, cognitive impairments are seen to peak during treatment and improve over time, which may occur due to reduced stress and worry, learning coping mechanisms, cognitive training and/or healing processes (such as white matter recovery) [94–96]. Other studies of breast cancer populations, however, have demonstrated no changes, or even a slight decline, in impairments at one-year post-treatment [39, 97, 98]. In the current study, where it was possible to investigate, meta-analytical methods demonstrated no difference in the outcomes of participants who were <5 years since cessation of therapy compared to >5 years since cessation of therapy (FSIQ and PIQ), although it should be noted that the included studies compared participants cross sectionally, rather than longitudinally following participants over time, where decline has previously been reported [48, 70]. There was also a very small number of studies that specifically investigated the effect of time post-treatment on cognitive outcomes within the pediatric population, highlighting important areas for future research.

In assessing the impact of treatment type on cognitive outcomes, it was expected that greater impairment, arising from central neurotoxicity, would be apparent in those with a history of CNS-directed treatment [99–102]. Central toxicity results in significant structural and functional changes in the brain, such as direct injury due to radiation or chemotherapy, neuroinflammation, oxidative stress, vascular changes, and white matter damage [99, 100]. These changes can disrupt key cell types (like astrocytes, microglia, and oligodendrocytes) and affect neuronal signaling, essential for cognitive functioning [103–105]. As expected, ALL was the most represented non-CNS cancer within this review and the use of intrathecal chemotherapy and/or cranial irradiation is commonly employed in ALL to prevent CNS relapse; therefore, we would expect this group to experience greater impairment compared to other non-CNS cancers with non-CNS directed treatment [106]. Contrary to our expectations, however, no significant differences were discovered when investigating participants who received non-CNS directed treatment compared to those that received CNS-directed treatment, although this may be due, at least in part, to smaller sample sizes in the meta-analysis for this specific subgroup investigation. Further, the timing of cognitive assessments may have influenced these findings, as cognitive function has been shown to change over time following cessation of treatment [107]. Indeed, a previous systematic review looking at cognitive impairment in survivors of pediatric central nervous system tumours highlighted that the frequency of cognitive impairments in paediatric CNS cancer survivors treated with CNS-directed therapy substantially increased with time since cessation of treatment, with mean frequencies of 30.9% within the first 5 years, 35.8% between 5 and 10 years, 44.8% over 10 years post-treatment, respectively, although how much of this was due to the presence of the tumour itself versus the administration of CNS-directed therapy, or what mechanisms may account for this increasing frequency with longer time following treatment, are difficult to disentangle [80]. Unfortunately, it is not possible to assess whether this also holds true in non-CNS cancer populations or for non-CNS directed treatment, as longitudinal studies were not well represented in this review.

Of note, the heterogeneity of measures of cognition and population sampling in the current study was significant and this poses a barrier to providing estimates of CRCI prevalence in the pediatric non-CNS cancer population with a high degree of confidence. Of note, the vast majority of measures employed in the included studies were not domain-specific, but rather general IQ measures, which offer a limited view of CRCI and fail to capture core reported impacts on memory, processing speed and executive function. Given this, future use of more comprehensive neuropsychological testing, which can be used to evaluate domain-specific impairments, should be promoted. In 2011, the International Cognition and Cancer Task Force (ICCTF) developed recommendations for a core set of neuropsychological tests, common criteria for defining cognitive impairment and cognitive change in cancer survivors and common methods to homogenize study methodology, including study design and analytical methods [36]. To date, in studies of CRCI employing neuropsychological testing, there has been some uptake in the use of the definition of cognitive impairment encouraged by the ICCTF (i.e. >2 SDs below the norm in one test, or 1.5 SDs below the norm in two tests, in a neuropsychological battery of recommended measures), with uptake increasing to 56.2% of studies in 2022, but declining to 40% of studies in 2023 [36, 108]. ICCTF recommendations have primarily been implemented in breast cancer patient populations and have not seen wide uptake in other types of cancer, particularly in the pediatric population [108]. Also of note, the definition of cognitive impairment here varied significantly across the included studies, with six studies employing >1 SD from the norm in a single test, which may be considered less stringent than the ICCTF recommendation [47, 51, 59, 64, 66, 73]. Taken together, these considerations emphasize the importance of implementing standardized guidelines, such as the ICCTF recommendations, in both the selection of cognitive assessment tasks utilized and the criteria employed for determining the presence of cognitive impairment, to order improve the reproducibility and reliability of findings between studies of CRCI in the pediatric population.

In addition to study design considerations, study reporting across the included studies was also an area of concern noted in the current work. Specifically, quality appraisal of the included studies revealed poor representation of appropriate sample size per treatment group (or at least justification of it), reliable measurements (ascertained via transparency in methodology) and appropriate reporting of the statistical analysis methods (i.e. reporting CIs). Poor, or complete lack of, reporting such important variables hindered the ability to synthesize all the data meaningfully or to provide a high degree of certainty in the evidence. Additionally, reporting around demographic and clinical characteristics meant additional risk factors, such as age at diagnosis, gender differences, and racial/ethnic differences, were not able to be explored. As a result, there were significant hurdles around harmonizing study designs and reporting of outcomes. Further, there are many confounding variables that may have further impacted the results, including cancer type, treatment type, dose and duration, time of assessment, hormones, and common health implications [107, 109–111]. Taken together, this leads to difficulty in developing meaningful evidence-based guidelines and recommendations around assessment, monitoring, and management of CRCI in survivors of non-CNS pediatric cancer.

### Conclusion

This review is the first systematic review investigating the prevalence of CRCI in non-CNS peadiatric cancer patients and survivors with meta-analytical assimilation. Results suggest clinically significant impairment may be present in 0-73% of patients across non-pooled data (0-67% for global cognitive function and intelligence and 0-73% for achievement, aptitude and school readiness, respectively), with pooled prevalence estimates of 12.46%-18.05% in measures of intelligence and 9.83%-12.05% in measures of attention. Certainty of evidence, however, was generally rated as low or very low. Therefore, these results should be interpreted with caution, as estimated prevalence rates may differ from actual prevalence rates. Future research should employ the ICCTF’s recommendations to harmonise studies of cognitive function, enabling more accurate estimates of prevalence and severity to be made. It is also crucial to address the gaps in the literature identified, including methodological heterogeneity, inconsistent criteria for defining cognitive impairment and a lack of longitudinal studies. This is essential for overcoming current barriers faced in the development of meaningful evidence-based guidelines and recommendations regarding assessment, monitoring, and management of CRCI in both patients and survivors of non-CNS pediatric cancer.

### Recommendations for practice

Children who have had childhood cancer, and have subsequently suffered cognitive changes, find their lives impacted on a personal, interpersonal, and academic level [93]. Of note, the data reported here emphasise that a significant number of survivors of non-CNS pediatric cancer may experience changes in measures of intelligence, attention and achievement/aptitude/school readiness These impacts may persist long-term, with many survivors experiencing slower developmental trajectories, lower levels of education attainment, higher risk of job discrimination, and lower levels of career success [20, 21, 112–115]. Despite this, there is a significant area of unmet need surrounding support and resources for the child, their families, and those in support positions, such as allied health and healthcare providers. There is a fundamental need to advocate for this population to ensure that they are protected and provided with resources.

Healthcare providers should be cognizant that survivors may experience cognitive changes in survivorship that can have long-lasting impacts on quality of life. Whilst there is a lack of standardized guidelines for both diagnosis and management, healthcare teams are encouraged to monitor and assess for changes in cognition (measured by objective tools or self-report) and provide resources/support where needs are identified. Within a paediatric population, particular considerations should be made towards identifying and aiding educational challenges and advocacy needs. Healthcare providers may like to promote awareness and discuss some non-therapeutic interventions that are commonly recommended (e.g., diet, exercise, memory aids, cognitive training, or occupational therapy). Better recognition of these impacts by healthcare professionals may aid in shifting the overall narrative for these survivors and encourage supportive legislation and the provision of needed supportive care resources, such as access to disability support, neuropsychological evaluation, occupational therapy and educational and workplace accomodations.

## Author Contributions

Authors Ines Semendric, Timothy H. Barker, Lyndsey E. Collins-Praino, and Alexandra L. Whittaker contributed to study conception and design. The search strategy was designed by Ines Semendric, Alexandra L. Whittaker, and library liaison Vikki Langton. Screening was conducted by Ines Semendric, Olivia J. Haller, and Alexandra L. Whittaker. Data curation was maintained by Ines Semendric, and data analysis was conducted by Ines Semendric, Timothy H. Barker, and Alexandra L. Whittaker. Original draft preparation was conducted by Ines Semendric, with critical revisions provided by Timothy H. Barker, Danielle Pollock, Lyndsey E. Collins-Praino, and Alexandra L. Whittaker. All authors read and approved the final manuscript.

## Data Availability

The datasets generated and analysed during the current study are available from the corresponding author on reasonable request.

---

**PRISMA 2020 Checklist**

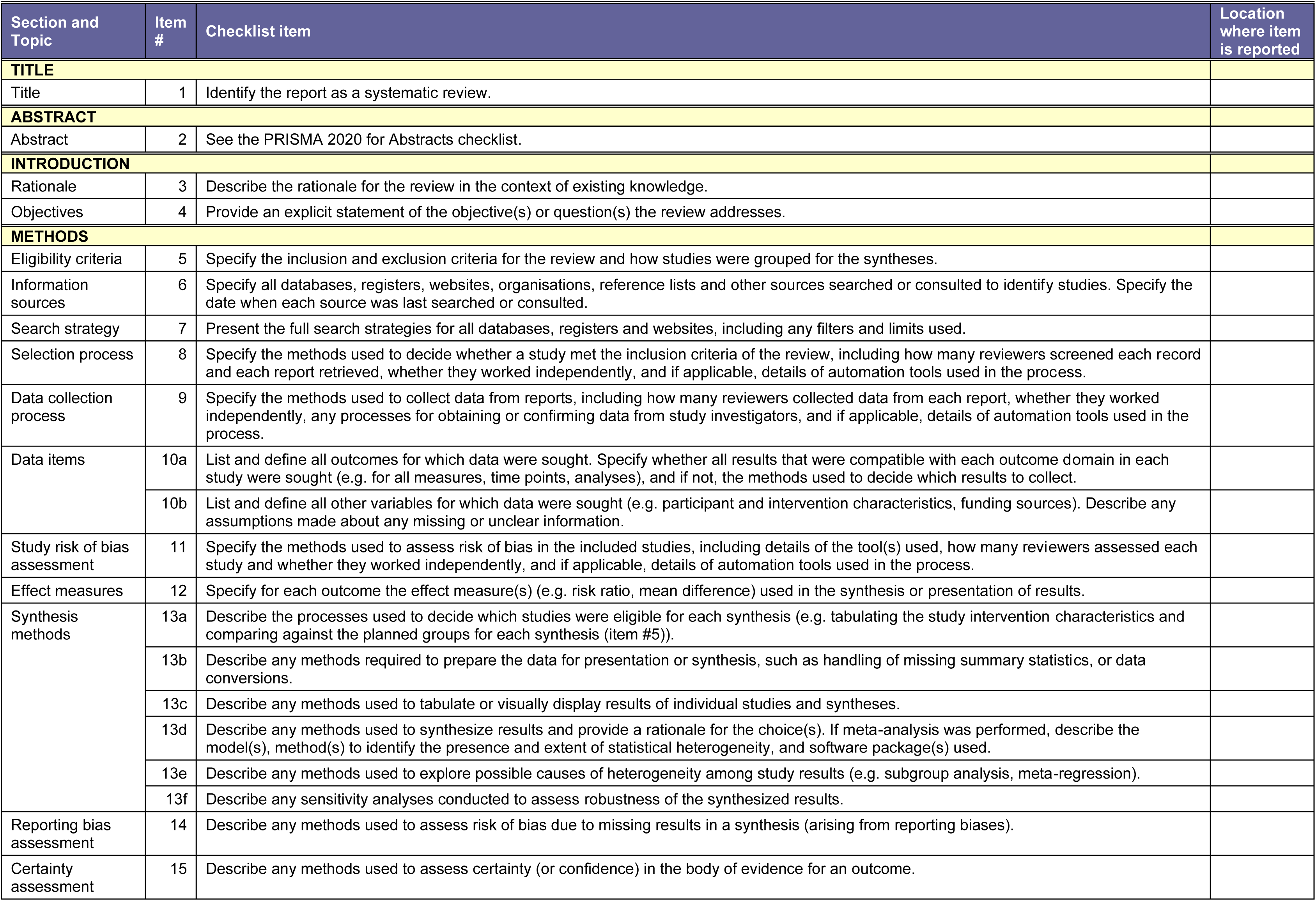
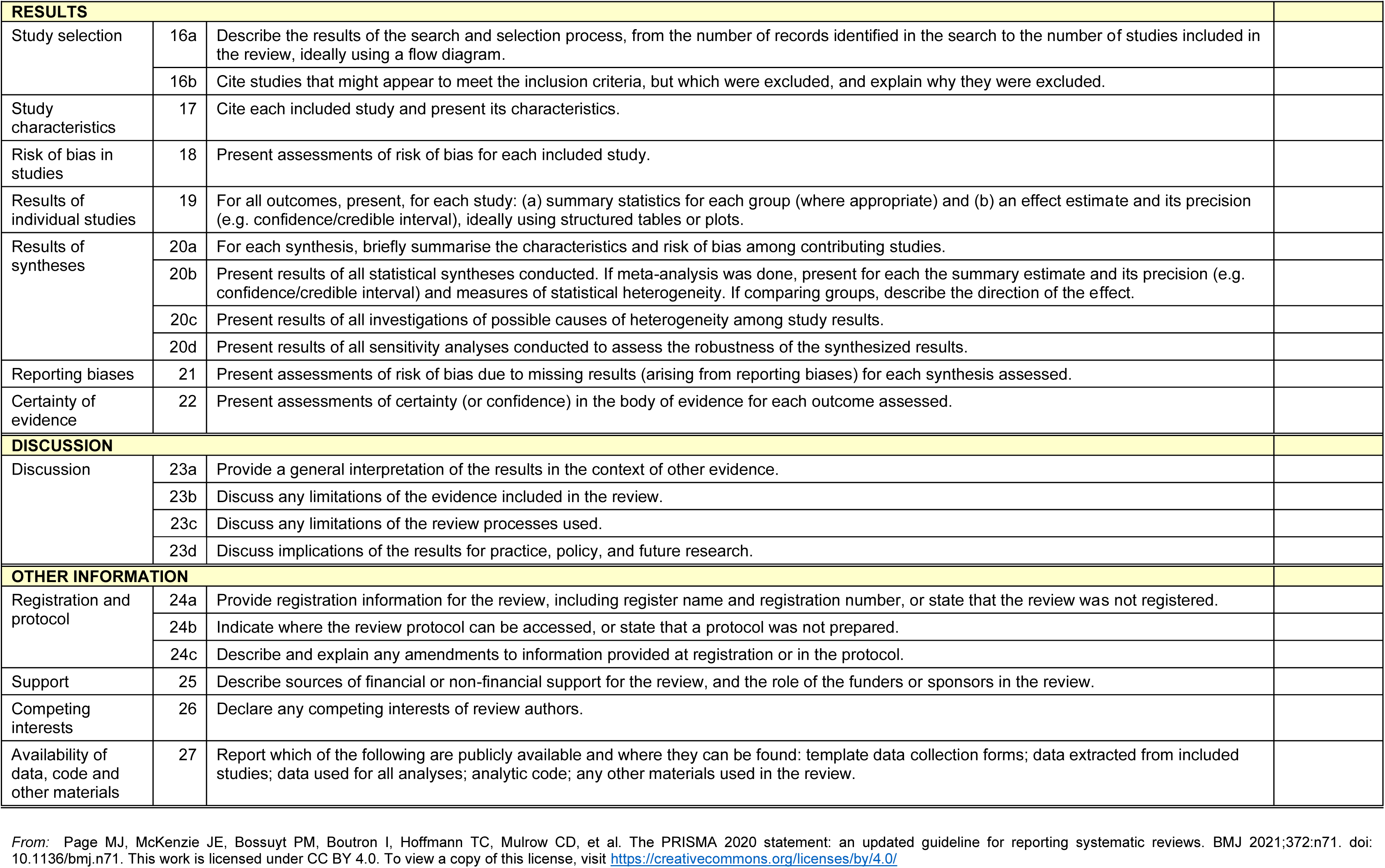

Online Resource 2: Search strategy

**Original search**

PubMed via MEDLINE

Date searched: 21/04/2023

| Search | Query | Records retrieved |
| --- | --- | --- |
| #1 | “Neoplasms”[mh] OR “Cancer Survivors”[mh] OR “Cancer*”[tiab] OR “Neoplas*”[tiab] OR “Patient”[tiab] OR “Survivor”[tiab] OR “Childhood cancer”[tiab] OR “Noncentral nervous system cancer*”[tiab] OR “Non-CNS cancer*”[tiab] OR “Leukemia”[tiab] OR “Lymphoma”[tiab] | 6,721,189 |
| #2 | “Antineoplastic combined chemotherapy protocols”[mh] OR “Neoadjuvant therapy”[mh] OR “Chemoradiotherapy”[mh] OR “Radioimmunotherapy”[mh] OR “Chemo*”[tiab] OR “Radiotherapy”[tiab] | 1,003,161 |
| #3 | “Neuropsychological tests”[mh] OR “Chemobrain”[tiab] OR “Chemofog”[tiab] OR “Chemotherapy induced cogn*”[tiab] OR “CICI”[tiab] OR “CRCI”[tiab] OR “Chemotherapy related cognitive changes”[tiab] OR “Cancer related cognitive impairment”[tiab] OR “Neuropsychol*”[tiab] OR “Cognitive dysfunction”[all] OR “Cognition disorders”[all] OR “Memory disorders”[all] OR “Language disorders”[all] OR “Neuropsychology”[all] | 324,932 |
| #4 | “Child”[mh] OR “Child*”[tiab] OR “Youth”[tiab] OR “Young adult”[tiab] | 2,762,440 |
| #5 | “Brain tumors”[all] OR “Brain tumours”[all] OR “CNS tumor”[all] OR “CNS tumour”[all] | 33,694 |
| #8 | #1 AND #2 AND #3 AND #4 NOT #5 | 490 |

Scopus

Date searched: 21/04/2023

Scopus Date searched: 21/04/2023
| Search | Query | Records retrieved |
| --- | --- | --- |
| #1 | ( TITLE-ABS ( neoplasms ) OR TITLE-ABS ( “cancer survivors” ) OR TITLE-ABS ( cancer* ) OR TITLE-ABS ( patient ) OR TITLE-ABS ( survivor ) OR TITLE-ABS ( “childhood cancer” ) OR TITLE-ABS ( “noncentral nervous system cancer*” ) OR TITLE-ABS ( “non-cns cancer*” ) OR TITLE-ABS ( leukemia ) OR TITLE-ABS ( lymphoma ) ) | 11,216,232 |
| **#2** | ( TITLE-ABS ( “antineoplastic combined chemotherapy protocols” ) OR TITLE-ABS ( “neoadjuvant therapy” ) OR TITLE-ABS ( chemoradiotherapy ) OR TITLE-ABS ( radioimmunotherapy ) OR TITLE-ABS ( chemo* ) OR TITLE-ABS ( radiotherapy ) ) | 1,129,863 |
| **#3** | ( TITLE-ABS ( “neuropsychological tests” ) OR TITLE-ABS ( chemobrain ) OR TITLE-ABS ( chemofog ) OR TITLE-ABS ( “chemotherapy induced cogn*” ) OR TITLE-ABS ( cici ) OR TITLE-ABS ( crci ) OR TITLE-ABS ( “chemotherapy related cognitive changes” ) OR TITLE-ABS ( “cancer related cognitive impairment” ) OR TITLE-ABS ( neuropsychol* ) OR ALL ( “cognitive dysfunction” ) OR ALL ( “cognition disorders” ) OR ALL ( “memory disorders” ) OR ALL ( “language disorders” ) OR ALL ( neuropsychology ) ) | 587,522 |
| **#4** | ( TITLE-ABS ( child* ) OR TITLE-ABS ( youth ) OR TITLE-ABS-KEY ( "Young Adult" ) ) | 3,532,086 |
| **#5** | ( ALL ( "Brain tumour*" ) OR ALL ( "Brain tumor*" ) OR ALL ( "CNS tumor*" ) OR ALL ( "CNS tumours*" ) ) | 371,576 |
| **#8** | **#1 AND #2 AND #3 AND #4 AND NOT #5** | 514 |

CINAHL Complete via EBSCO Date searched: 21/04/2023

| **Search** | **Query** | **Records retrieved** |
| --- | --- | --- |
| **#1** | ( (MH "Neoplasms") OR (MH "Cancer Survivors") ) OR TI ( Cancer* OR Neoplasm* OR Patient OR Survivor OR "Childhood Cancer" OR "Noncentral Nervous System Cancer" OR "Non-CNS Cancer*" OR "Leukemia" OR "Lymphoma" ) OR AB ( Cancer* OR Neoplasm* OR Patient OR Survivor OR "Childhood Cancer" OR "Noncentral Nervous System Cancer" OR "Non-CNS Cancer*" OR "Leukemia" OR "Lymphoma" ) | 2,415,460 |
| **#2** | ( (MH "Antineoplastic Agents, Combined") OR (MH "Chemotherapy, Cancer") OR (MH "Neoadjuvant Therapy") OR (MH “Radiotherapy”) ) OR TI ( Chemo * OR Radiotherapy ) OR AB ( Chemo * OR Radiotherapy ) | 104,296 |
| **#3** | ( MH “Neuropsychological tests” OR MH “Chemotherapy-Related Cognitive Impairment” ) OR TI ( Chemobrain OR Chemofog OR CICI or CRCI OR Neuropsychol* ) OR AB ( Chemobrain OR Chemofog OR CICI or CRCI OR Neuropsychol* ) OR TX ( "Cognitive dysfunction" OR "Cognitive impairment" OR "Memory Disorders" OR "Language Disorders" OR "Neuropsychol*" ) | 141,518 |
| **#4** | ( (MH “Child”) OR (MH "Adolescence") ) OR TI ( Child* OR Youth OR "Young Adult" ) OR AB ( Child* OR Youth OR "Young Adult" ) | 1,152,951 |
| **#5** | (MH "Brain Neoplasms") OR TX ( "Brain tumor*" OR "Brain tumour*" OR "CNS tumor*" OR "CNS tumour*" ) | 28,873 |
| **#8** | **#1 AND #2 AND #3 AND #4 NOT #5** | 101 |

PsycINFO via OvidSP

Date searched: 21/04/2023

PsycINFO via OvidSP Date searched: 21/04/2023
| **Search** | **Query** | **Records retrieved** |
| --- | --- | --- |
| **#1** | exp Neoplasms/ or ("cancer survivors" or patient or survivor or "childhood cancer" or "noncentral nervous system cancer" or "non-CNS cancer" or leukemia or lymphoma).ti. or ("cancer survivors" or patient or survivor or "childhood cancer" or "noncentral nervous system cancer" or "non-CNS cancer" or leukemia or lymphoma).ab. | 319,849 |
| **#2** | exp Antineoplastic Drugs/ or exp Chemotherapy/ or exp Radiation Therapy/ or (chemo* or radiotherapy).ti. or (chemo* or radiotherapy).ab. | 16,933 |
| **#3** | Exp Neuropsychological Assessment/ or exp Psychometric/ or exp Cognitive Impairment/ or exp Neuropsychology/ or (Chemobrain or chemofog or CICI or CRCI or neuropsychol*).ti. or (Chemobrain or chemofog or CICI or CRCI or neuropsychol*).ab. or (memory disorders or language disorders).mp. | 344,908 |
| **#4** | (child or adolescen* or youth or "young adult").ti. or (child or adolescen* or youth or "young adult").ab. | 568,664 |
| **#5** | Exp Brain Neoplasms/ or ("brain tumor*" or "brain tumour" or "CNS tumor" or "CNS tumour").mp. | 5,018 |
| **#8** | **#1 AND #2 AND #3 AND #4 NOT #5** | 35 |

**Updated search**

PubMed via MEDLINE Date searched: 03/06/2024

| **Search** | **Query** | **Records retrieved** |
| --- | --- | --- |
| **#1** | "Neoplasms"[mh] OR "Cancer Survivors"[mh] OR "Cancer*"[tiab] OR "Neoplas*"[tiab] OR "Patient"[tiab] OR "Survivor"[tiab] OR "Childhood cancer"[tiab] OR "Noncentral nervous system cancer*"[tiab] OR "Non-CNS cancer*"[tiab] OR "Leukemia"[tiab] OR "Lymphoma"[tiab] | 7,128,772 |
| **#2** | "Antineoplastic combined chemotherapy protocols"[mh] OR "Neoadjuvant therapy"[mh] OR "Chemoradiotherapy"[mh] OR "Radioimmunotherapy"[mh] OR "Chemo*"[tiab] OR "Radiotherapy"[tiab] | 1,068,230 |
| **#3** | "Neuropsychological tests"[mh] OR "Chemobrain"[tiab] OR "Chemofog"[tiab] OR "Chemotherapy induced cogn*"[tiab] OR "CICI"[tiab] OR "CRCI"[tiab] or "Chemotherapy related cognitive changes"[tiab] OR "Cancer related cognitive impairment"[tiab] OR "Neuropsychol*"[tiab] OR "Cognitive dysfunction"[all] OR "Cognition disorders"[all] OR "Memory disorders"[all] OR "Language disorders"[all] OR "Neuropsychology"[all] | 339,555 |
| **#4** | "Child"[mh] OR "Child*"[tiab] OR "Youth"[tiab] OR "Young adult"[tiab] | 2,880,472 |
| **#5** | "Brain tumors"[all] OR "Brain tumours"[all] OR "CNS tumor"[all] OR "CNS tumour"[all] | 36,101 |
| **#8** | **#1 AND #2 AND #3 AND #4 NOT #5 (filter date 21/04/2023-03/06/2024)** | 18 |

Scopus

Date searched: 03/04/2024

Scopus Date searched: 03/04/2024
| **Search** | **Query** | **Records retrieved** |
| --- | --- | --- |
| #1 | ( TITLE-ABS ( neoplasms ) OR TITLE-ABS ( “cancer survivors” ) OR TITLE-ABS ( cancer* ) OR TITLE-ABS ( patient ) OR TITLE-ABS ( survivor ) OR TITLE-ABS ( “childhood cancer” ) OR TITLE-ABS ( “noncentral nervous system cancer*” ) OR TITLE-ABS ( “non-cns cancer*” ) OR TITLE-ABS ( leukemia ) OR TITLE-ABS ( lymphoma ) ) | 12,022,335 |
| #2 | ( TITLE-ABS ( “antineoplastic combined chemotherapy protocols” ) OR TITLE-ABS ( “neoadjuvant therapy” ) OR TITLE-ABS ( chemoradiotherapy ) OR TITLE-ABS ( radioimmunotherapy ) OR TITLE-ABS ( chemo* ) OR TITLE-ABS ( radiotherapy ) ) | 1,206,454 |
| #3 | ( TITLE-ABS ( “neuropsychological tests” ) OR TITLE-ABS ( chemobrain ) OR TITLE-ABS ( chemofog ) OR TITLE-ABS ( “chemotherapy induced cogn*” ) OR TITLE-ABS ( cici ) OR TITLE-ABS ( crci ) OR TITLE-ABS ( “chemotherapy related cognitive changes” ) OR TITLE-ABS ( “cancer related cognitive impairment” ) OR TITLE-ABS ( neuropsychol* ) OR ALL ( “cognitive dysfunction” ) OR ALL ( “cognition disorders” ) OR ALL ( “memory disorders” ) OR ALL ( “language disorders” ) OR ALL ( neuropsychology ) ) | 640,641 |
| #4 | ( TITLE-ABS ( child* ) OR TITLE-ABS ( youth ) OR TITLE-ABS-KEY ( "Young Adult" ) ) | 3,760,804 |
| #5 | ( ALL ( "Brain tumour*" ) OR ALL ( "Brain tumor*" ) OR ALL ( "CNS tumor*" ) OR ALL ( "CNS tumours*" ) ) | 411,498 |
| #8 | #1 AND #2 AND #3 AND #4 AND NOT #5 (filter year 2023-2024) | 31 |

CINAHL Ultimate via EBSCO Date searched: 03/06/2024

| Search | Query | Records retrieved |
| --- | --- | --- |
| #1 | ( (MH "Neoplasms") OR (MH "Cancer Survivors") ) OR TI ( Cancer* OR Neoplasm* OR Patient OR Survivor OR "Childhood Cancer" OR "Noncentral Nervous System Cancer" OR "Non-CNS Cancer*" OR "Leukemia" OR "Lymphoma" ) OR AB ( Cancer* OR Neoplasm* OR Patient OR Survivor OR "Childhood Cancer" OR "Noncentral Nervous System Cancer" OR "Non-CNS Cancer*" OR "Leukemia" OR "Lymphoma" ) | 2,462,224 |
| #2 | ( (MH "Antineoplastic Agents, Combined") OR (MH "Chemotherapy, Cancer") OR (MH "Neoadjuvant Therapy") OR (MH "Radiotherapy") ) OR TI ( Chemo * OR Radiotherapy ) OR AB ( Chemo * OR Radiotherapy ) | 109,598 |
| #3 | ( MH "Neuropsychological tests" OR MH "Chemotherapy-Related Cognitive Impairment" ) OR TI ( Chemobrain OR Chemofog OR CICI or CRCI OR Neuropsychol* ) OR AB ( Chemobrain OR Chemofog OR CICI or CRCI OR Neuropsychol* ) OR TX ( "Cognitive dysfunction" OR "Cognitive impairment" OR "Memory Disorders" OR "Language Disorders" OR "Neuropsychol*" ) | 184,897 |
| #4 | ( (MH "Child") OR (MH "Adolescence") ) OR TI ( Child* OR Youth OR "Young Adult" ) OR AB ( Child* OR Youth OR "Young Adult" ) | 1,194,390 |
| #5 | (MH "Brain Neoplasms") OR TX ( "Brain tumor*" OR "Brain tumour*" OR "CNS tumor*" OR "CNS tumour*" ) | 34,481 |
| #8 | **#1 AND #2 AND #3 AND #4 NOT #5 (filter date April 2023-June 2024)** | 6 |

PsycINFO via OvidSP

Date searched: 03/06/2024

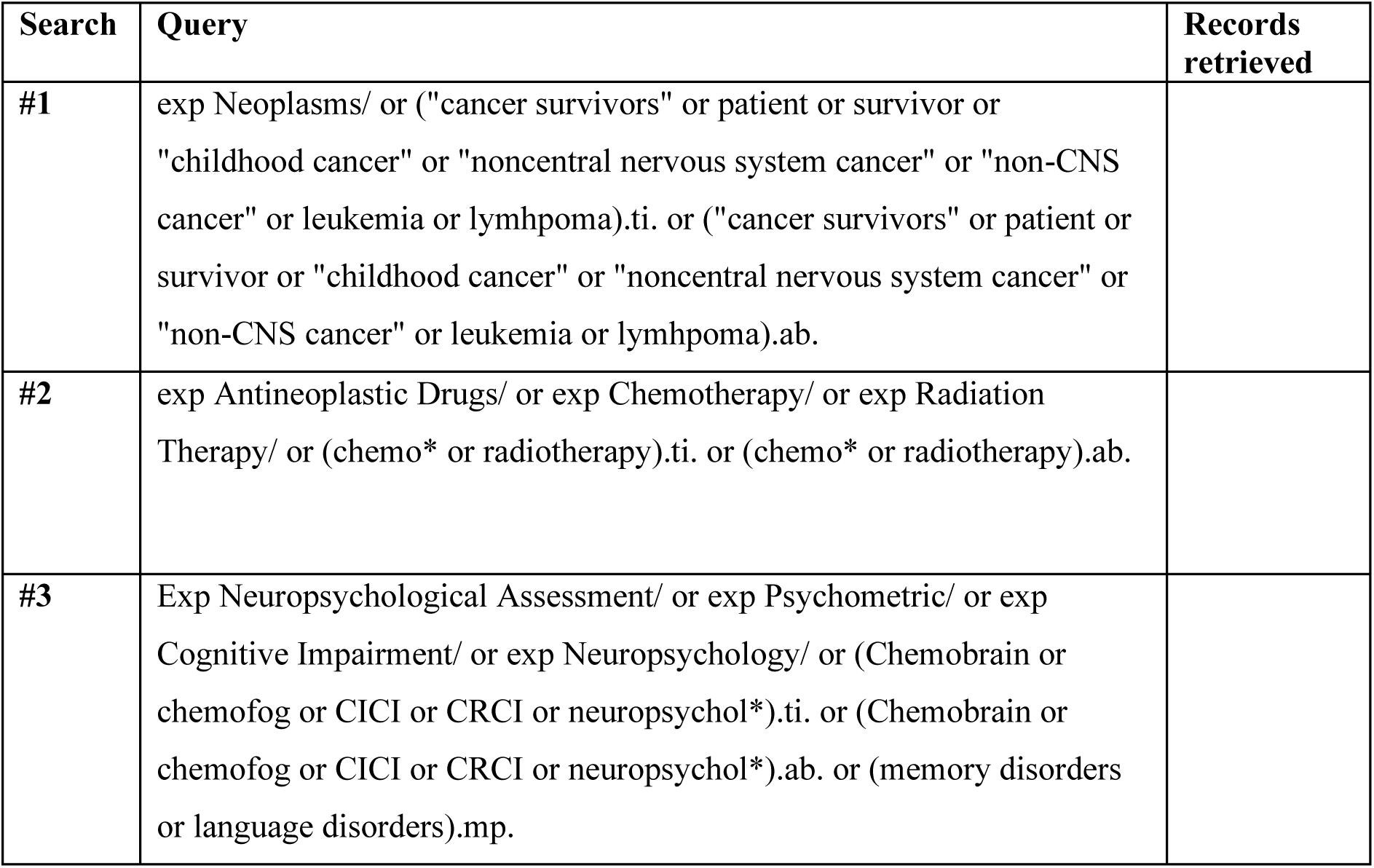
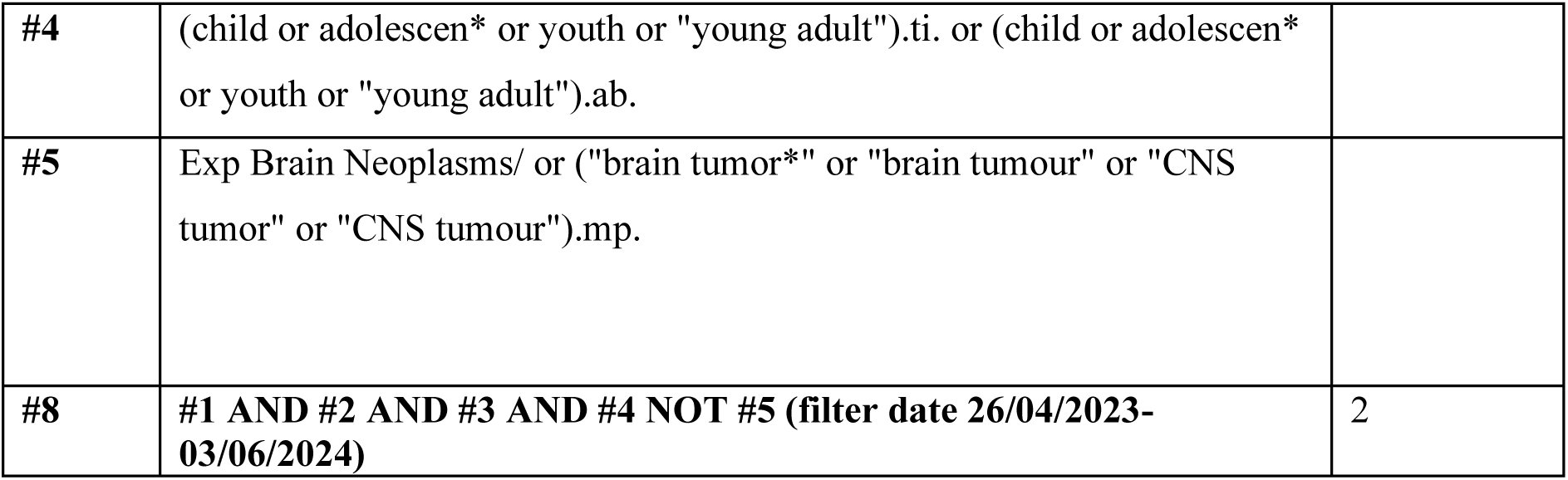

**Online Resource 3: Critical appraisal tool**

JBI CRITICAL APPRAISAL CHECKLIST FOR STUDIES REPORTING PREVALENCE DATA

|  | Yes | No | Unclear | Not applicable |
| --- | --- | --- | --- | --- |
| 1. Was the sample frame appropriate to address the target population? | ☐ | ☐ | ☐ | ☐ |
| 2. Were study participants sampled in an appropriate way? | ☐ | ☐ | ☐ | ☐ |
| 3. Was the sample size adequate? | ☐ | ☐ | ☐ | ☐ |
| 4. Were the study subjects and the setting described in detail? | ☐ | ☐ | ☐ | ☐ |
| 5. Was the data analysis conducted with sufficient coverage of the identified sample? | ☐ | ☐ | ☐ | ☐ |
| 6. Were valid methods used for the identification of the condition? | ☐ | ☐ | ☐ | ☐ |
| 7. Was the condition measured in a standard, reliable way for all participants? | ☐ | ☐ | ☐ | ☐ |
| 8. Was there appropriate statistical analysis? | ☐ | ☐ | ☐ | ☐ |
| 9. Was the response rate adequate, and if not, was the low response rate managed appropriately? | ☐ | ☐ | ☐ | ☐ |

Overall appraisal: Include ☐ Exclude ☐ Seek further info ☐
Comments (Including reason for exclusion)

**JBI CRITICAL APPRAISAL CHECKLIST FOR STUDIES REPORTING PREVALENCE DATA**

*Adaptions included from* Whittaker, A. L., George, R. P., & O’Malley, L. (2022). Prevalence of cognitive impairment following chemotherapy treatment for breast cancer: a systematic review and meta-analysis. *Scientific reports*, *12*(1), 2135. https://doi.org/10.1038/s41598-022-05682-1

Answers: Yes, No, Unclear or Not/Applicable

1. **Was the sample frame appropriate to address the target population?**

This question relies upon knowledge of the broader characteristics of the population of interest and the geographical area. For survivors of non-CNS childhood cancers, the characteristics, demographics and medical history is needed. Particular consideration will be towards reporting of diagnosis, treatment regime, age at diagnosis, age at treatment, age at testing, time since cessation of treatment (if applicable), and time since commencing treatment (if applicable).

1. **Were study participants recruited in an appropriate way?**

Studies may report random sampling from a population, and the methods section should report how sampling was performed. Random probabilistic sampling from a defined subset of the population (e.g., hospital/region) should be employed in most cases, however, random probabilistic sampling is not needed when everyone in the sampling frame will be included/ analysed. Convenience samples, such as a street survey or interviewing lots of people at public gatherings are not considered to provide a representative sample of the base population. Studies deemed to be low risk of bias clearly describe how participants were sourced, e.g., through out-patient records or support groups, and the manner of sampling technique to reach the required sample size, for instance consecutive sampling of all patients that met the inclusion criteria.

If a study reports how participants were sourced (e.g., hospital outpatient centre) AND manner of sampling (e.g., purposive sampling), they would receive a “YES”. If a study reports one, but not the other, they receive an “Unclear”, if they do not report either they receive a “No”.

1. **Was the sample size adequate?**

The larger the sample, the narrower will be the confidence interval around the prevalence estimate, making the results more precise. An adequate sample size is important to ensure good precision of the final estimate. Ideally we are looking for evidence that the authors conducted a sample size calculation to determine an adequate sample size. This will estimate how many subjects are needed to produce a reliable estimate of the measure(s) of interest.

For conditions with a low prevalence, a larger sample size is needed. Also consider sample sizes for subgroup (or characteristics) analyses, and whether these are appropriate.

Sometimes, the study will be large enough (as in large national surveys) whereby a sample size calculation is not required. In these cases, sample size can be considered adequate.

When there is no sample size calculation, an arbitrary sample size of 50 (per treatment group) was considered adequate based on the value suggested by Dijkshoorn et al 2021.

1. **Were the study subjects and setting described in detail?**

Certain diseases or conditions vary in prevalence across different geographic regions and populations. The study sample should be described in sufficient detail so that other researchers can determine if it is comparable to the population of interest to them.

Particularly, it is important that patient demographics are included, treatment details, and the setting (e.g., hospital/out-patient).

1. **Was data analysis conducted with sufficient coverage of the identified sample?**

Coverage bias can occur when not all subgroups of the identified sample respond at the same rate. For instance, you may have a very high response rate overall for your study, but the response rate for a certain subgroup (i.e. older adults) may be quite low. Factors considered were bias of age/racial status and differences in mean income/education status. This is particularly relevant where self-report was used as a method of assessment.

1. **Were valid methods used for the identification of the condition?**

This criterion is looking at measurement or classification bias. If a validated assessment tool was used, a “yes” answer would be selected. However, self-report methods may lead to a risk of over-reporting or under-reporting and non-validated self-report methods would lead to a “no” answer. Whilst there is some controversy over the use of short cognitive screening tools for CRCI, due to their inability to detect subtle cognitive decline, for the purposes of this question they were regarded as validated tests.

1. **Was the condition measured in a standard, reliable way for all participants?**

Considerable judgment is required to determine the presence of some health outcomes. Having established the validity of the outcome measurement instrument (see item 6 of this scale), it is important to establish how the measurement was conducted. How many observers performed the tests, and if there were multiple observers was there a comparison of results between the observers? Were those involved in collecting data trained or educated in the use of the instrument/s, which may be particularly vital for administering neuropsychological tests? If there was more than one data collector, were they similar in terms of level of education, clinical or research experience, or level of responsibility in the piece of research being appraised? When there was more than one observer or collector, was there comparison of results from across the observers? Was the condition measured in the same way for all participants? Given the nature of patients potentially being sick and experiencing fatigue from treatment, consideration was given to when these tests were performed e.g., directly after treatment, and whether the location of testing may have influenced the test oucomes.

1. **Was there appropriate statistical analysis?**

Importantly, the numerator and denominator should be clearly reported, and percentages should be given with confidence intervals. The methods section should be detailed enough for reviewers to identify the analytical technique used and how specific variables were measured. These criteria were applied strictly with a failure to report confidence intervals around prevalence estimates leading to a high-risk rating.

1. **Was the response rate adequate, and if not, was the low response rate managed appropriately?**This criterion considered the dropout rate from study start to completion. Consideration was however given to the fact that dropout is inevitable particularly in longer term studies where patients may have died or been lost to follow up. If attrition was evident, an examination of the attrition rates for each group was performed. If rates were fairly similar, then a low risk of bias rating was awarded. Authors should also discuss the response rate in the study and reasons for non-response. If this was done well with the conclusion that reasons for non-response are unrelated to the outcome measured, and the characteristics of non-responders are comparable to those measured, then a low risk of bias was awarded.

**Online Resource 4: Data extraction template**

**General information**

Title

Title of paper / abstract / report that data are extracted from

Country in which the study conducted United States

UK

Canada Australia Other

Notes

**Characteristics of included studies**

Methods Aim of study Study design

Randomised controlled trial

Non-randomised experimental study Cohort study

Cross sectional study Case control study Systematic review Qualitative research Prevalence study Case series

Case report

Diagnostic test accuracy study Clinical prediction rule Economic evaluation

Text and opinion Other

Cohort study (if applicable)

**Participants**

Population description

Sex

Diagnosis

Age at diagnosis Stage of treatment

Currently undergoing treatment In remission

Time since cessation of treatment Identified confounds

Total number of participants Timepoint of cognitive report Method of cognitive assessment

Self-report

Objective Neuropsychological testing

Imaging

Notes on specifics of cognitive assessment

Author’s criteria for determining cognitive impairment

Prevalence report for chemobrain (% or number to convert, standard deviation, confidence intervals)

**Online Resource 5 Table 1.** Cognitive classification of tests employed

*\*Please refer to main manuscript for references*

| Cognitive test | Cognitive domain(s) | Outcomes reported | Study* |
| --- | --- | --- | --- |
| ***Global cognitive function and intelligence*** |  |  |  |
| Wechsler Scales<br>Wechsler Memory Scale IV<br>Wechsler Intelligence Scale for Children (WISC, WISC Revised, WISC-III, WISC-IV, Wechsler Adult Intelligence Scale (WAIS, WAIS Revised) Wechsler Abbreviated Scale of Intelligence (WASI)<br>Hamburg-Wechsler Intelligence Scale for Adults | General cognitive function and intelligence | Fullscale IQ, verbal IQ, performance IQ, immediate and delayed memory, logical memory, design memory, visual reproduction and verbal paired association, digit span (forward and backwards), vocabulary, block design, matrix reasoning, processing speed index, digit symbol, symbol search, spatial span (forward and backwards), verbal cognition, perceptual reasoning, working memory | Armstrong et al 2013 [47]; Chiou et al 2019 [50]; Cole et al 2015 [51]; Gandy et al 2023 [52]; Giralt et al 1992 [53]; Halberg et al 1992 [54]; Iuvone et al 2002 [58]; Jannoun & Chessells 1987 [59]; Kokkonen et al 1997 [61]; Kolotas et al 2001 [62]; Krawczuk-Rybak et al 2012 [63]; Kunin-Batson, Kadan-Lottick & Neglia 2014 [64]; Lofstad et al 2009 [65]; Meadows et al 1981 [66]; Mizera 2014 [67]; Robison et al 1984 [69]; Rubenstein, Varni & Katz 1990 [70]; von der Weid et al 2003 [73]; Waber et al 1990 [47] |
| Snijders-Oomen Nonverbal Intelligence Scale | General intelligence | Fullscale IQ | Harten et al 1984 [55] |
| Stanford-Binet Form L-M | General cognitive ability including visual preference, attention, memory, sensorimotor, exploration and manipulation, concept formation | Fullscale IQ | Meadows et al 1981 [66] |
| HAWIK-R (German or French) | General intelligence | Fullscale IQ | von der Weid et al 2003 [72] |
| Illinois Test of Psycholinguistic Abilities (PET) | Spoken and written language abilities in children | Verbal expression | Harten et al 1984 [55] |
| McCarthy Scales of Children's Abilities | Cognitive ability in children in 6 domains: Verbal, Perceptual-Performance, Quantitative, General Cognitive, Memory, and Motor | Fullscale IQ | Meadows et al 1981 [66] |
| Halstead Category Test | Abstract reasoning | Abstract reasoning | Meadows et al 1981 [66] |
| Peabody Picture Vocabulary Test | Receptive vocabulary | Receptive vocabulary | Meadows et al 1981 [66] |
| Ravens Progressive Matrices | Non-verbal abstract reasoning, proxy for general cognitive ability | Abstract reasoning | Meadows et al 1981 [66] |
| Brief Cognitive Status Exam (BCSE) | Global cognitive function | Scored as average, low average, borderline, low, and very low | Armstrong et al 2013 [47] |
| Continuous Performance Task (CPT, CPT-II, CPT-III) | Attention, primarily used in ADHD research or clinically where attention deficits are expected | HRT ISI, HRT SD, variability, omissions, detectability, perseverations, commissions, HIT reaction time, HIT SD, D-Prime, Beta, percentile, response time | Cheung et al 2023 [49]; Chiou et al 2019 [50]; Gandy et al 2023 [52]; Kunin-Batson, Kadan-Lottick & Neglia 2014 [64] |
| Wisconsin Card Sorting Test (WCST) | Set shifting, cognitive flexibility, executive function | Error, perseverative responses, perseverative error, non-perseverative error, conceptual level response | Chiou et al 2019 [50]; Gandy et al 2023 [52] |
| Rey Osterrieth Complex Figure | Visuo-constructional ability and visual memory | Overall score, copy (immediate and delayed score) | Cole et al 2015 [51]; Gandy et al 2023 [52] |
| Rey Auditory Verbal Learning | Attention, working memory, verbal learning, memory | Overall score | Jannoun & Chessells 1987 [59] |
| Delis-Kaplan Executive Function System (DKEFS) | Higher cognitive function including verbal and non-verbal executive functions, verbal/spatial fluency, problem solving, verbal and spatial concept formation, cognitive flexibility, verbal inhibition, planning and reasoning, impulsivity, verbal and spatial abstract thinking, and metaphorical and creative thinking | 20 question initial abstraction, 20 question total, color word read, color word inhibit, color word inhibit switch<br>Tower test<br>Trail making task (TMT) number sequence, TMT letter sequence, TMT number-letter switch<br>Verbal fluency (VF) letter fluency, VF category fluency, VF category switch | Gandy et al 2023 [52] |
| Wide Range Assessment of Memory and Learning (WRAML) | Short and long-term memory (immediate and delayed), ability to learn new material in both children in adults | Story memory recall, verbal learning memory recall, visual learning memory recall, visual memory index, verbal memory index | Hill et al 2004 [57] |
| NEPSY | Assesses neuropsychological development across six functional domains:<br>1) Attention and Executive Functioning,<br>2) Language,<br>3) Memory and Learning,<br>4) Sensorimotor,<br>5) Social Perception, and<br>6) Visuospatial Processing | Attention, language, visuospatial function, memory | Pääkkö et al 2000 [68] |
| Token Test | Verbal comprehension of commands of increasing complexity | Overall score | Iuvone et al 2002 [58] |
| Modified Bells test | Visuospatial analysis and attention | Rapidity, accuracy | Iuvone et al 2002 [58] |
| 16 Word | Auditory memory | Short-term and long-term memory | Iuvone et al 2002 [58] |
| Cogstate battery | Visual memory, processing speed, psychomotor function, executive function (delayed recall for visual memory, delayed reverse recall for spatial working memory), attention, working memory | Continuous Paired Associate Learning (CPAL), detection (DET), Groton Maze Learning (GML), identification (IND), one back (ONB) | Sands et al 2017 [71] |
| ***Achievement, aptitude and school readiness*** |  |  |  |
| Wide Range Achievement Test Revised (WRAT-R) | Academic skills, ability to read words, comprehend sentences, spell, and compute solutions to math problems | Cognitive impairment index | Waber et al 1990 [73] |
| Wechsler Individual Achievement Test 2<sup>nd</sup> Edition Abbreviated (WIAT) | Academic skills/achievement in reading, maths, written language, oral language | Word reading, numerical operations, spelling | Kunin-Batson, Kadan-Lottick & Neglia 2014 [64] |
| Detroit Test of Learning Aptitude | Global cognitive function | Learning disability | Armstrong et al 2013 [47] |
| Test of Reading Comprehension (TORC) | Relational vocabulary, sentence completion, paragraph construction, text comprehension, contextual fluency | Cognitive Impairment Index | Waber et al 1990 [73] |
| BAS Word Reading Test | Word decoding, recognition of printed words, visual and auditory working memory, word analysis, vocabulary knowledge | Overall score | Jannoun & Chessells 1987 [59] |

**Online Resource 6 Table 1.** JBI Critical Appraisal Checklist for studies reporting prevalence data outcomes

| Citation | Q1<br>Sample<br>frame | Q2<br>Appropriate<br>sampling | Q3<br>Sample<br>size | Q4 Study<br>subjects/setting | Q5<br>Sufficient<br>coverage | Q6<br>Valid<br>methods | Q7<br>Measured<br>reliably | Q8<br>Appropriate<br>statistical<br>analysis | Q9<br>Response<br>rate | Methodological<br>quality |
| --- | --- | --- | --- | --- | --- | --- | --- | --- | --- | --- |
| Armstrong<br>2013 | Y | U | Y | Y | Y | Y | U | Y | Y | High |
| Brown<br>1992 | Y | Y | N | Y | Y | Y | U | N | Y | Moderate |
| Cheung<br>2023 | Y | Y | Y | Y | Y | Y | U | Y | Y | High |
| Chiou 2019 | Y | Y | N | Y | U | Y | U | N | Y | Moderate |
| Cole 2015 | Y | Y | Y | Y | Y | Y | U | Y | Y | High |
| Gandy<br>2023 | Y | Y | Y | Y | Y | Y | Y | Y | Y | High |
| Giralt 1992 | U | Y | N | Y | Y | Y | U | N | Y | Moderate |
| Halberg<br>1992 | Y | U | N | N | U | Y | Y | N | Y | Low |
| Harten<br>1984 | Y | U | U | Y | U | Y | U | N | U | Low |
| Heukrodt<br>1988 | Y | N | N | N | U | Y | U | N | Y | Low |
| Hill 2004 | Y | Y | N | N | U | Y | U | N | Y | Low |
| Iuvone<br>2002 | Y | Y | N | Y | Y | Y | U | N | Y | Moderate |
| Jannoun<br>1987 | Y | Y | N | U | Y | Y | U | N | Y | Moderate |
| Kokkonen<br>1997 | Y | Y | N | U | U | Y | U | N | Y | Low |
| Kolotas<br>2001 | Y | Y | N | U | Y | Y | U | N | Y | Moderate |
| Krawczuk-Rybak 2012 | Y | U | N | Y | Y | Y | Y | N | Y | Moderate |
| Kunin-Batson 2014 | Y | Y | Y | Y | Y | Y | Y | N | Y | High |
| Lofstad 2009 | Y | Y | N | Y | Y | Y | U | N | Y | Moderate |
| Meadows 1981 | Y | U | N | N | U | Y | U | N | Y | Low |
| Mizera 2014 | Y | Y | N | U | U | Y | U | N | Y | Low |
| Pääkkö 2000 | Y | Y | N | U | U | Y | Y | N | Y | Moderate |
| Robison 1984 | Y | Y | Y | Y | Y | Y | U | N | N | Moderate |
| Rubenstein 1990 | Y | U | N | Y | Y | Y | U | N | Y | Moderate |
| Sands 2017 | Y | Y | N | Y | Y | Y | Y | N | Y | High |
| vonderWeid 2003 | Y | Y | Y | Y | Y | Y | U | N | Y | High |
| Waber 1990 | Y | Y | U | U | Y | Y | U | N | Y | Moderate |
| % | 96.15 | 73.07 | 26.92 | 61.53 | 65.38 | 100 | 23.07 | 15.38 | 92.30 |  |

**Online Resource 7 Table 1.** Global cognitive function and intelligence outcomes

*\*Please refer to main manuscript for references*

| Study* | Assessment method | Methodological quality | Definition of cognitive impairment | Timepoint(s) | Prevalence % (95% CIs where available) |
| --- | --- | --- | --- | --- | --- |
| Armstrong et al 2013 [47] | Wechsler Memory Scale IV<br>Brief Cognitive Status Exam (BCSE)<br>Wechsler Abbreviation Scale of Intelligence (WASI) | High | z-score more than 1 SD below the normal mean of zero<br>Compared with age-adjusted normative rates | One timepoint assessing survivors | **Group 1 (18Gy):**<br>**Wechsler Memory Domains**<br>Immediate 17.5% (11.2-25.2)<br>Delayed 15.1% (9.3-22.5)<br>Auditory 16.7% (10.6-24.3)<br>Visual 13.5% (8.1-20.7)<br>**Logical memory**<br>Immediate 15.0% (9.2-22.4)<br>Delayed n=15 11.9% (6.8-18.9)<br>**Design memory**<br>Immediate 8.7% (4.4-15.1)<br>Delayed 3.2% (0.9-7.9)<br>**Verbal paired association**<br>Immediate 10.3% (5.6-17)<br>Delayed 9.5% (5.0-16.0)<br>**Visual reproduction**<br>Immediate n=16 12.6% (7.4-19.6)<br>n=14 Delayed 11.0% (6.2-17.8)<br>**Brief Cognitive Status Exam**<br>Low Average 16.5%<br>Borderline 5.5% |
|  |  |  |  |  | Low 8.7%<br>Very low 0.0%<br>**Group 2 (24Gy):**<br>**Wechsler Memory Domains**<br>Immediate 33.8% (25.9-42.4)<br>Delayed 30.2% (22.6-38.6)<br>Auditory 30.7% (23.1-39.1)<br>Visual 21.3% (14.8-29.2)<br>**Logical memory**<br>Immediate 23.2% (16.4-31.1)<br>Delayed 28.3% (20.9-36.6)<br>**Design memory**<br>Immediate 14.7% (9.2-21.8)<br>Delayed 13.2% (8.0-20.1)<br>**Verbal paired association**<br>Immediate 20.4% (14.0-28.2)<br>Delayed 19.0% (12.8-26.6)<br>**Brief Cognitive Status Exam**<br>Low Average 15.2%<br>Borderline 8.7%<br>Low 11.6%<br>Very low 6.5% |
| Cheung et al<br>2023 [49] | Conners'<br>Continuous<br>Performance<br>Test III (CPT-III) | High | A higher T-score<br>indicative of more<br>severe<br>impairment/problems.<br>Compared to age- | One<br>timepoint<br>assessing<br>survivors | CPT HRT ISI Change (sustained attention) 2.9<br>(0.0-6.0)<br>HRT SD (sustained attention) 7.8 (2.6-13.0)<br>Variability (inattentiveness) 2.9 (0.0-6.2)<br>Omissions (inattentiveness) 7.8 (2.6-13.0) |
|  |  |  | adjusted T-scores (mean = 50, SD = 10).<br>Impairment % was defined as a score poorer than 1.5 standard deviation of age-adjusted T-scores of reference norms. |  | Detectability (inattentiveness) 12.8 (6.2-19.2)<br>Perseverations (impulsivity) 2.9 (0.0-6.0)<br>Commissions (impulsivity) 4.9 (0.7-9.0) |
| Chiou et al 2019 [50] | Weschler Intelligence Scale for Children-IV (WISC-IV)<br>Conners' Continuous Performance Test (CPT-II) version 5<br>Wisconsin card sorting test (WCST) | Moderate | WISC-IV: fullscale IQ score <70<br>CPT-II: Test scores compared with population norms by computer algorithm<br>WCST = less than five percentile rank considered atypical | One timepoint assessing survivors | **WISC-IV**<br>7.1%<br>**CPT-II**<br>Omission rate (ie, problem in attention) 10%<br>Comission rate (ie, problem in impulsivity control) 20%<br>Detectability 22.5%<br>**WCST**<br>42.8% |
| Cole et al 2015 [51] | Weschler Intelligence Scale for Children-III or Wechsler Adult Intelligence Scale | High | WISC-III or WAIS: scores more than one SD from the reference population (IQ <85, Digit Span, Vocabulary, Block Design or Matrix Reasoning <7) | One timepoint assessing survivors | **WISC-III or WAIS**<br>Fullscale IQ 11% (8 to 15)<br>Wechsler Digit Span 30% (25 to 35)<br>Wechsler Vocabulary 15% (11 to 19)<br>Wechsler Block Design 17% (13 to 23)<br>Wechsler Matrix Reasoning 18% (12 to 25)<br>**Rey-Osterrieth Complex Figure Organization** |
|  | Ray-Osterrieth Complex Figure |  | Ray-Osterrieth Complex Figure: scores < or equal to 25<sup>th</sup> percentile for age |  | 57% (51 to 62) |
| Gandy et al 2023 [52] | Rey-Osterrieth Complex Figure (Rey)<br>Wechsler Abbreviated Scale of Intelligence (WASI)<br>Continuous Performance Task (CPT)<br>Wechsler Intelligence Scale for Children (WISC)<br>Delis-Kaplan Executive Function System (DKEFS) | High | Normative, age-standardized Z scores calculated. Expected impairment rate in normative sample 10%. | One timepoint assessing survivors | **Rey Complex Figure**<br>Copy 120 (58.25%; 51.46-65.0)<br>Immediate copy 101 (49.27%; 42.37-56.17)<br>Delayed copy 106 (51.96%; 45.05-58.87)<br>**WASI**<br>Verbal IQ 22 (10.73%; 6.46-15.0)<br>Performance IQ 19 (9.18%; 5.21-13.14)<br>Fullscale IQ 21 (10.24%; 6.06-14.43)<br>Vocabulary 28 (13.66%; 8.92-18.40)<br>Block design 32 (15.46; 10.49-20.42)<br>Similarities 22 (10.73%; 6.46-15.00)<br>Matrix reasoning 22 (10.63%; 6.39-14.86)<br>**WCST**<br>Error 22 (10.68%; 6.43-14.93)<br>Perseverative response 28 (13.59%; 8.87-18.31)<br>Perseverative error 29 (14.08%; 9.29-18.87)<br>Nonperseverative error 16 (7.77%; 4.08-11.45)<br>Conceptual level response 18 (8.74%; 4.85-12.63)<br>**CPT**<br>Omissions 30 (14.78%; 9.85-19.70)<br>Commissions 25 (12.32; 7.76-16.87)<br>Hit reaction time 26 (12.81%; 8.17-17.44)<br>Hit standard error 32 (15.76%; 10.71-20.82) |
|  |  |  |  |  | Variability of standard error 31 (15.27%; 10.28-20.26)<br>D-Prime 21 (10.34%; 6.12-14.57)<br>Beta 17 (8.37%; 4.53-12.22)<br>Percentile 30 (14.78%; 9.85-19.70)<br>**Wechsler**<br>Digit span forward 43 (20.77%; 15.20-26.35)<br>Digit span backward 36 (17.39%; 12.18-22.60)<br>Processing Speed Index 31 (14.90%; 10.02-19.78)<br>Digit symbol 37 (17.87%; 12.61-23.14)<br>Symbol search 28 (13.46%; 8.78-18.14)<br>Spatial span forward 35 (16.83%; 11.70-21.95)<br>Spatial span backward 25 (12.08%; 7.60-16.55)<br>**DKEFS**<br>20 question initial abstraction 37 (18.05%;12.74-23.36)<br>20 question total 30 (14.63%; 9.76-19.51)<br>Color word read 23 (11.27%; 6.90-15.65)<br>Color word inhibit 33 (16.26%; 11.14-21.37)<br>Color word inhibit switch 29 (14.36%; 9.48-19.23)<br>Tower test 21 (10.10%; 5.97-14.22)<br>TMT number sequence 36 (17.39; 12.18-22.60)<br>TMT letter sequence 43 (20.77%; 15.20-26.35)<br>TMT number-letter switch 54 (26.09%; 20.06-32.12)<br>VF letter fluency 50 (24.27%; 18.37-30.18)<br>VF category fluency 30 (14.56%; 9.71-19.42)<br>VF category switch 22 (10.68%; 6.43-14.93) |
| Giralt et al<br>1992 [53] | Wechsler<br>Intelligence | Moderate | IQ <80 | One<br>timepoint | **WISC** |
|  | Scale for Children (WISC) |  |  | assessing survivors | **Chemotherapy group**<br>IQ <80 10.3%<br>IQ of 50 3.4%<br>**Chemotherapy + radiotherapy group**<br>IQ <80 16%<br>IQ of 50 8% |
| Halberg et al 1992 [54] | Wechsler Intelligence Scale for Children-Revised (WISC-R) | Low | IQ <90 | One timepoint assessing survivors | **WISC-R**<br>**Group 1 (2400 cGy)**<br>10.54%<br>**Group 2 (1800 cGy)**<br>43.75%<br>**Control Group (Wilms)**<br>16.66%<br>*Cumulative (Group 1+2+Control)*<br>23.40% |
| Harten et al 1984 [55] | (PET) Illinois Test of Psycholinguistic Abilities (GB: Verbal expression)<br>For ages 4-5, 5-9<br>(SON) Snijders-Oomen Nonverbal Intelligence Scale | Low | Grade 1 = Average or above<br>Grade 2 = Low average range<br>Grade 3 = Dysfunctional<br>Grade 4 = Severe changes | One timepoint assessing survivors | **Group 1 (ALL)**<br>**Verbal expression**<br>Grade 1: 47%<br>Grade 2: 37%<br>Grade 3: 16%<br>Grade 4: 0%<br>**IQ nonverbal**<br>Grade 1: 84%<br>Grade 2: 8%<br>Grade 3: 8%<br>Grade 4: 1.9% |
|  | For ages 4-5, 5-9, 9-14, 14-18<br>Hamburg-Wechsler Intelligence Scale for Adults<br>For ages 18+ |  |  |  | **Group 2 (Control)**<br>**Verbal expression**<br>Grade 1: 29%<br>Grade 2: 59%<br>Grade 3: 12%<br>Grade 4: 0%<br>**IQ nonverbal**<br>Grade 1: 83%<br>Grade 2: 14%<br>Grade 3: 0%<br>Grade 4: 3.4% |
| Heukrodt et al 1988 [56] | Wide Range Achievement Test (WRAT) | Low | Scores below the 20<sup>th</sup> percentile considered learning disability | One timepoint assessing survivors | **Group 1 (ALL)**<br>50%<br>**Group 2 (Solid tumors)**<br>36% |
| Hill et al 2004 [57] | Wide Range of Assessment and Learning (WRAML) | Low | Atypical<br>Borderline<br>Low Average<br>Average<br>Bright Average | One timepoint assessing survivors | *NR = not reported*<br>*Data transformed from no. to %*<br>**WRAML**<br>**Story memory recall difference score**<br>Atypical 0%<br>Borderline 20%<br>Low Average NR<br>**Verbal learning memory recall**<br>Atypical NR<br>Borderline NR<br>Low Average NR<br>**Visual learning memory recall** |
|  |  |  |  |  | Atypical NR<br>Borderline NR<br>Low Average NR |
| Iuvone et al<br>2002 [58] | Wechsler's Intelligence Scale for Children-Revised (WISC-R) and Adult Intelligence Scale-Revised (WAIS-R) for >16 years<br>Token Test<br>16-word test<br>Modified Bells Test | Moderate | Total IQ <70 = mild mental retardation<br>>70 but <80 = Borderline intelligence<br>"Low scores" for the remainder. | One timepoint assessing survivors | *Data transformed from no. to %*<br>**WISC-R or WAIS-R**<br>Mild mental retardation 4.7%<br>Borderline Intelligence 23.8%<br>**Token test** 4.7%<br>**16-word test**<br>Short-term memory 4.7%<br>Long-term memory 9.5%<br>**Modified Bells Test**<br>Rapidity 61.9%<br>Accuracy 61.9% |
| Jannoun & Chessells<br>1987 [59] | Wechsler Intelligence Scale for Children-Revised (WISC-R)<br>Rey Auditory Verbal Learning | Moderate | WISC-R: 1 SD lower than average<br>Rey Auditory Verbal Learning: score below the 10<sup>th</sup> percentile compared to age-related norms | One timepoint assessing survivors | **WISC-R**<br>42%<br>**Rey Auditory Verbal Learning**<br>44% |
| Kokkonen et al 1997 [61] | Wechsler Adult Intelligence Scale | Low | Not reported | One timepoint assessing survivors | *Data transformed from no. to %*<br>**WAIS**<br>Verbal IQ 40.7%<br>Performance IQ 11.1% |
| Kolotas et al 2001 [62] | Hamburg Wechsler Intelligence Test for Adults | Moderate | IQ <70 corresponds to clinical designation of severe mental retardation | One timepoint assessing survivors | **Hamburg**<br>3.7% |
| Krawczuk-Rybak et al 2012 [63] | Wechsler Intelligence Scale for Children Revised (WISC-R) for ages 6-16<br>Wechsler Adult Intelligence Scale Revised (WAIS-R) for ages >16 | Moderate | Score <85 | One timepoint assessing survivors | *Data transformed from no. to %*<br>**WISC-R or WAIS-R**<br>Performance IQ 3.2% |
| Kunin-Batson, Kadan-Lottick & Neglia 2014 [64] | Wechsler Intelligence Scale for Children-IV (WISC-IV)<br>Conners Continuous Performance | High | Norm-referenced | One timepoint assessing survivors | **WISC-IV**<br>Verbal Cognitive 9.1%<br>Perceptual Reasoning 8.7%<br>Working Memory 12.5%<br>Processing Speed 24.7%<br>Full scale IQ 9.2%<br>**Conners CPT-II**<br>Omission errors 3.8% |
|  | Test 2<sup>nd</sup> Ed<br>(CPT-II) |  |  |  | Commission errors 12.2%<br>Response time 9.5%<br>Variability 6.8% |
| Lofstad et al<br>2009 [65] | Wechsler<br>Intelligence<br>Scale for<br>Children 3rd Ed<br>(WISC-III)<br>Swedish version<br>with Norwegian<br>translation | Moderate | Compared to<br>performance of control<br>group and WISC-III<br>test norms | One<br>timepoint<br>assessing<br>survivors | **WISC-III**<br>Fullscale IQ 5.7% (*data transformed from no. to*<br>*%*)<br>**Long-term Follow-up**<br>Verbal working memory 12.8%<br>Spatial working memory 10.9%<br>Spatial construction 15.4%<br>Verbal span 18.6%<br>Spatial span 14.6%<br>Visual-motor speed 15.4%<br>Visual speed 11.5% |
| Meadows et al<br>1981 [66] | Stanford-Binet<br>Form L-M<br>Wechsler<br>Intelligence<br>Scale for<br>Children-Revised<br>Peabody Picture<br>Vocabulary Test<br>Halstead<br>Category Test<br>Ravens | Low | Impaired score <1 SD<br>(expected 16%)<br>Moderate to severe<br>impairment <2 SD<br>(expected 2%) | One<br>timepoint<br>assessing<br>survivors | **Wechsler Intelligence Scales/Stanford-**<br>**Binet/McCarthy Scales**<br>Impaired 33%<br>Moderately to severely impaired 11%<br>**Other measures**<br>Impaired<br>Receptive vocabulary 0%<br>Expressive vocabulary 28%<br>Abstract reasoning 33%<br>Academic achievement 39%<br>General memory 67%<br>Auditory sequence 39%<br>Visual perception 40%<br>Moderately to severely impaired |
|  | Progressive<br>Matrices<br>McCarthy Scales<br>of Children's<br>Abilities |  |  |  | Receptive vocabulary 0%<br>Expressive vocabulary 0%<br>Abstract reasoning 17%<br>Academic achievement 11%<br>General memory 22%<br>Auditory sequence 17%<br>Visual perception 0% |
| Mizera 2014<br>[67] | Wechsler<br>Intelligence<br>Scale for<br>Children 4th Ed<br>(WISC-IV)<br>or Wechsler<br>Abbreviated<br>Scale of<br>Intelligence<br>(WASI) | Low | WISC-IV and WASI<br>test norms | One<br>timepoint<br>post-status of<br>treatment | **WISC-IV or WAIS**<br>12% |
| Pääkkö et al<br>2000 [68] | NEPSY | Moderate | Not reported | One<br>timepoint<br>assessing new<br>survivors at<br>the end of<br>treatment/in<br>remission | **NEPSY**<br>**Irradiated patients**<br>Attention 14%<br>Language 36%<br>Visuospatial functions 57%<br>Memory 64%<br>**Non-irradiated patients**<br>Attention 43%<br>Language 29%<br>Visuospatial functions 21%<br>Memory 36%<br>**Cumulative**<br>Attention 29% |
|  |  |  |  |  | Language 32%<br>Visuospatial functions 39%<br>Memory 50% |
| Robison et al 1984 [69] | Wechsler Intelligence Scale for Children Revised (WISC-R) | Moderate | Not defined by authors but standard score range for WISC-IV considers<br>90-109 = Average<br>80-89 = Low Average<br>70-79 = Borderline<br><69 = Extremely Low | One timepoint assessing survivors | *Data transformed from no. to %*<br>**WISC-R Verbal IQ**<br>Low Average 22%<br>Borderline 6%<br>Extremely Low 4%<br>**Performance IQ**<br>Low Average 14%<br>Borderline 6%<br>Extremely Low 4%<br>**Full-scale IQ**<br>Low Average 18%<br>Borderline 6%<br>Extremely Low 4% |
| Rubenstein, Varni & Katz 1990 [70] | Age-appropriate Wechsler Intelligence Scale | Moderate | Not reported | Two timepoints<br>Timepoint 1 (at diagnosis)<br>Timepoint 2 (4 to 5-year follow-up) | *Data transformed from no. to %*<br>**Timepoint 1:**<br>**Wechsler**<br>All scored within average range<br>**Timepoint 2:**<br>**Wechsler**<br>Fullscale IQ |
|  |  |  |  |  | Low-average 12.5%<br>Borderline 8.3% |
| Sands et al<br>2017 [71] | Cogstate battery:<br>Continuous Paired Associate Learning (CPAL)<br>Detection (DET)<br>Groton Maze Learning (GML)<br>Identification (IND)<br>One Back (ONB) | High | Scores of 2 or more SD below the population mean but investigated more liberal criteria and presented impairment rates for <1 SD and <1.5 SD as well. | One timepoint assessing patients at baseline (first 4 weeks of treatment) | **CPAL**<br>6% <2 SD<br>12.5% <1.5 SD<br>18.8% <1 SD<br>**DET**<br>3% <2 SD<br>12.5% <1.5 SD<br>21.9% <1 SD<br>**GML**<br>0% <2 SD<br>6.3% <1.5 SD<br>12.5% <1 SD<br>**IND**<br>6% <2 SD<br>9.4% <1.5 SD<br>15.6% <1 SD<br>**ONB**<br>0% <2 SD<br>0% <1.5 SD<br>3.1% <1 SD |
| von der Weid et al<br>2003 [72] | Wechsler's Intelligence Test (WISC-R) | High | Borderline <85 score<br>Mental retardation <70 score | One timepoint | **WISC-R, HAWIK-R, or HAWIE-R:** |
|  | German version<br>HAWIK-R<br>French version<br>HAWIE-R |  |  | assessing<br>survivors | **Fullscale IQ**<br>**ALL**<br>Borderline 10%<br>Mental retardation 2%<br>**Other non-CNS cancers**<br>Borderline 13%<br>Mental retardation 1% |
| Waber et al<br>1990 [73] | *See ESM 7 Table 1 “Achievement, aptitude and school readiness” as Cognitive Impairment Index presented incorporates both IQ and academic achievement thus fits both criteria. For the sake of statistical analysis, the prevalence estimates from this study will only be incorporated in the “Achievement, aptitude and school readiness” domain.* |  |  |  |  |

**Online Resource 8: Meta-analyses**

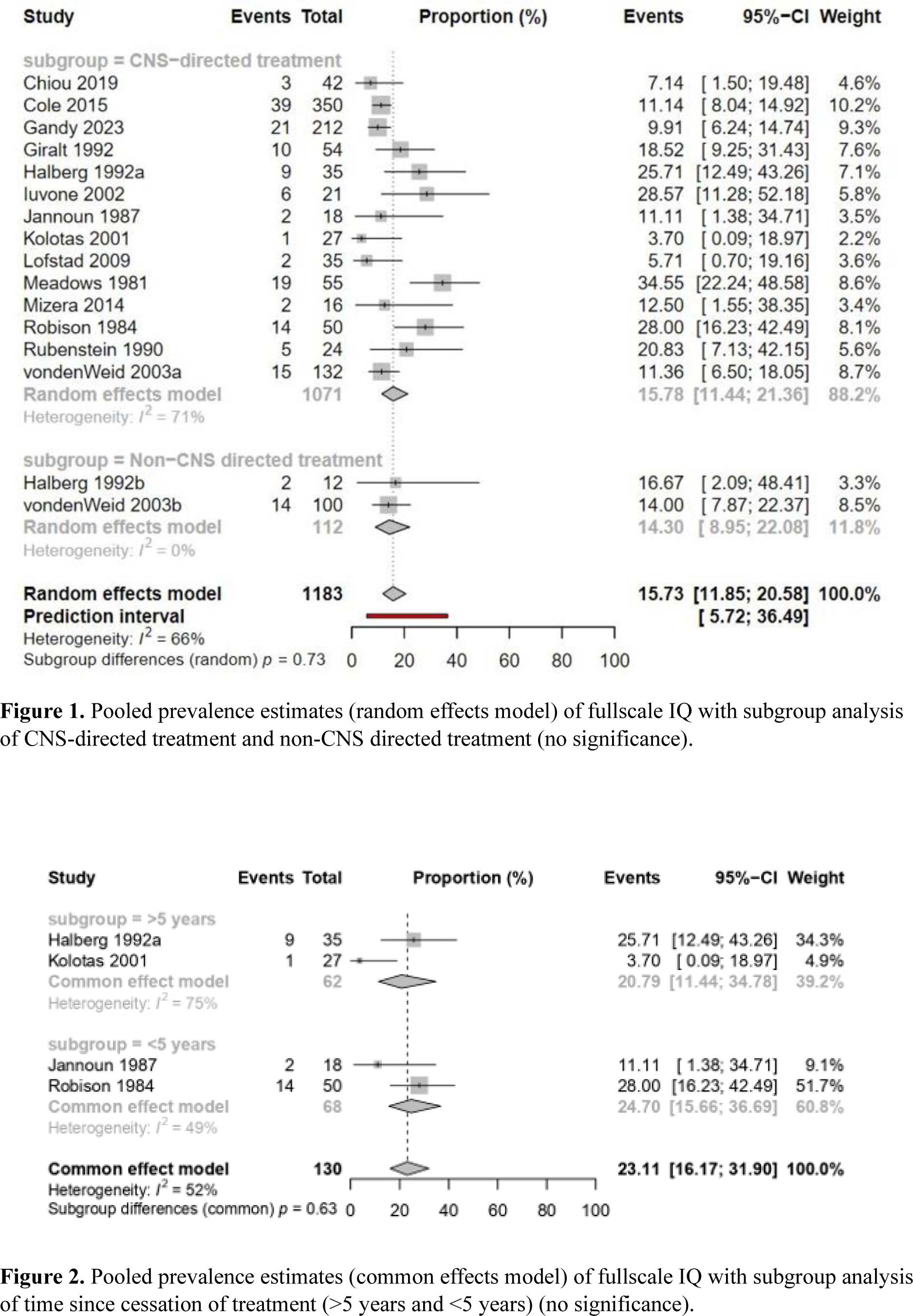
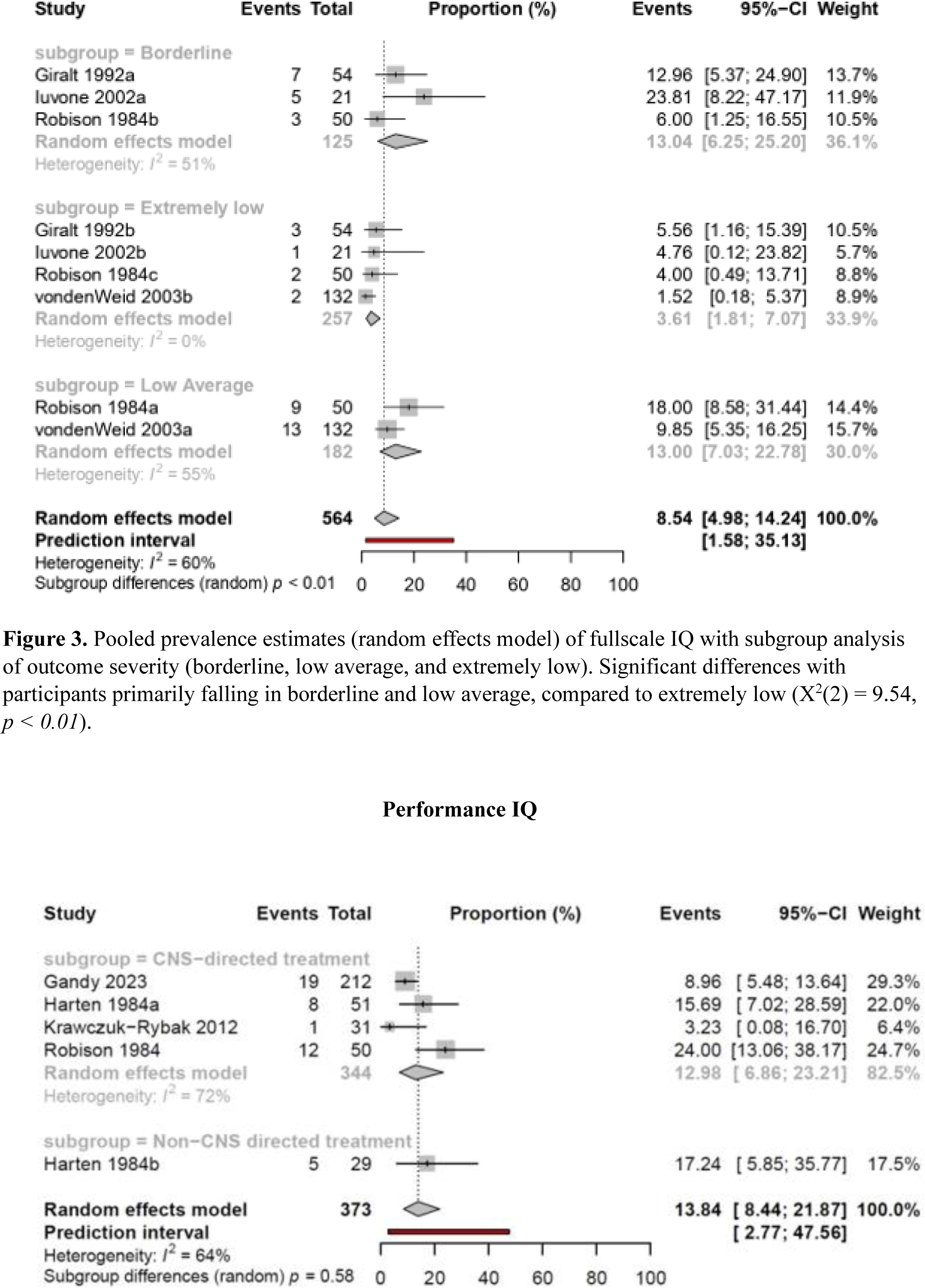
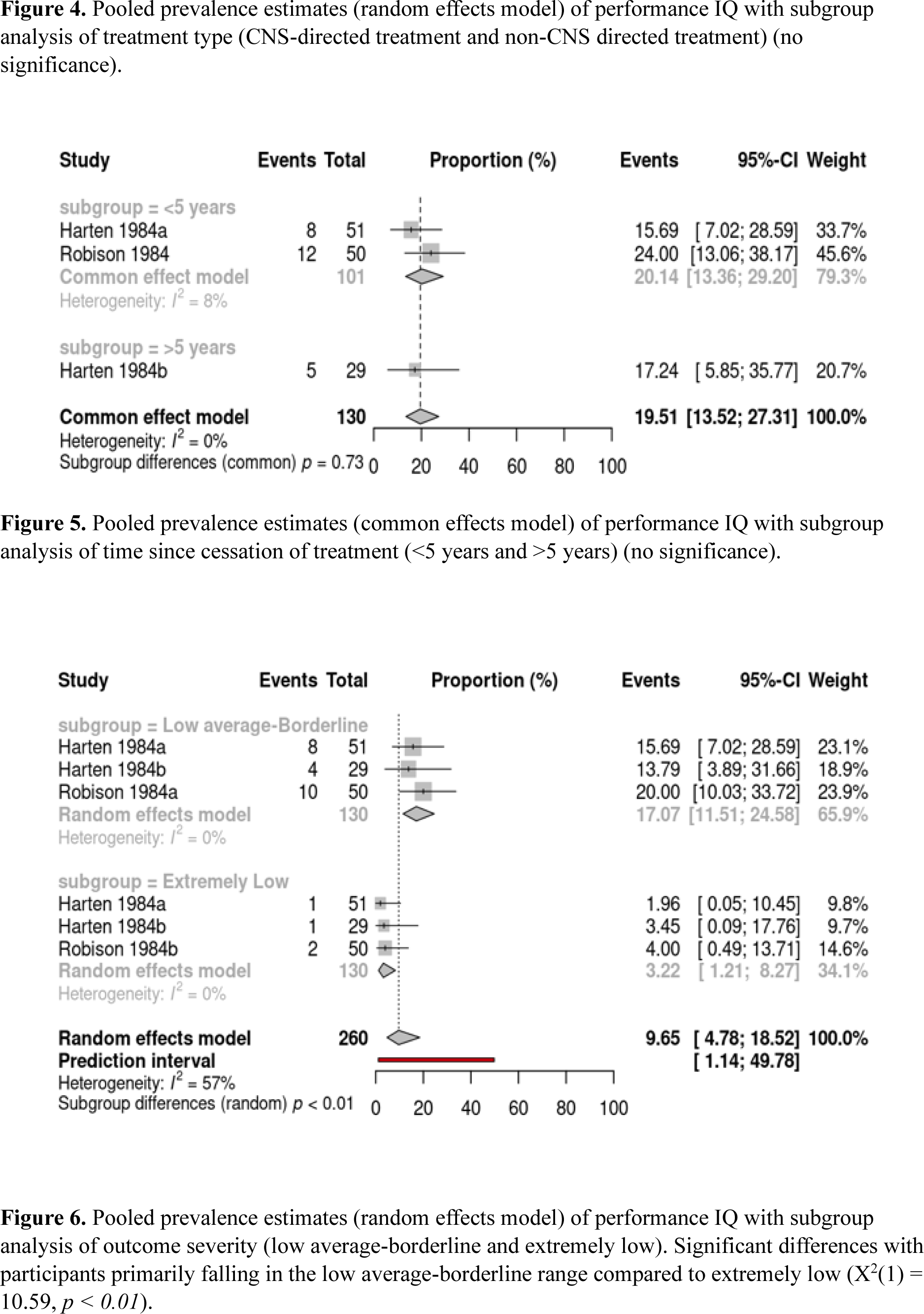

**Online Resource 9 Table 1**. Achievement, aptitude and school readiness outcomes

*\*Please refer to main manuscript for references*

| Study* | Assessment method | Methodological quality | Definition of cognitive impairment | Timepoint(s) | Prevalence % (95% CIs where available) |
| --- | --- | --- | --- | --- | --- |
| Brown et al 1992 [48] | Detroit Test of Learning Aptitude | Moderate | Not reported | One timepoint assessed per group<br>Group 1 was newly diagnosed, Group 2 was assessed 1 year into treatment, and Group 3 was assessed 1-year post-treatment | **Group 1 (newly diagnosed):**<br>Learning disability 0%<br>**Group 2 (1-year into treatment):**<br>Learning disability 15.4%<br>**Group 3 (1-year post-treatment):**<br>Learning disability 60% |
| Jannoun & Chessells 1987 [59] | BAS Word Reading Test | Moderate | Score below the 25<sup>th</sup> percentile compared to age-related norms | One timepoint assessing survivors | **Word Reading Test**<br>44% |
| Kunin-Batson, Kadan-Lottick & Neglia 2014 [64] | Wechsler Individual Achievement Test 2<sup>nd</sup> Ed Abbreviated (WIAT-II) | High | Score falling more than one standard deviation below the population mean (norm-referenced) | One timepoint assessing survivors | **WIAT-II**<br>Word reading 8.7%<br>Numerical operations 15.2%<br>Spelling 12.2% |
| Waber et al 1990 [73] | Wechsler Intelligence Scale (WISC-R) | Moderate | Cognitive Impairment Index incorporating both IQ | One timepoint assessing survivors | **Group 1 (ALL): Cognitive Impairment Index** |
|  | Wide Range<br>Achievement Test<br>(WRAT-R) |  | and academic<br>achievement<br>0 = IQ >90, within 1<br>SD of all academic<br>tests<br>1 = IQ >90, 1 SD<br>below mean on 1/2<br>tests<br>2 = IQ >90, 1 SD<br>below mean on <3<br>tests<br>3 = IQ <90, 1 SD<br>below mean on <3<br>tests<br>4 = IQ <90, 1 SD<br>below mean on all<br>four tests |  | 73%<br>**Group 2 (Wilms’):**<br>**Cognitive**<br>**Impairment Index**<br>26% |

**Online Resource 10: Publication bias**

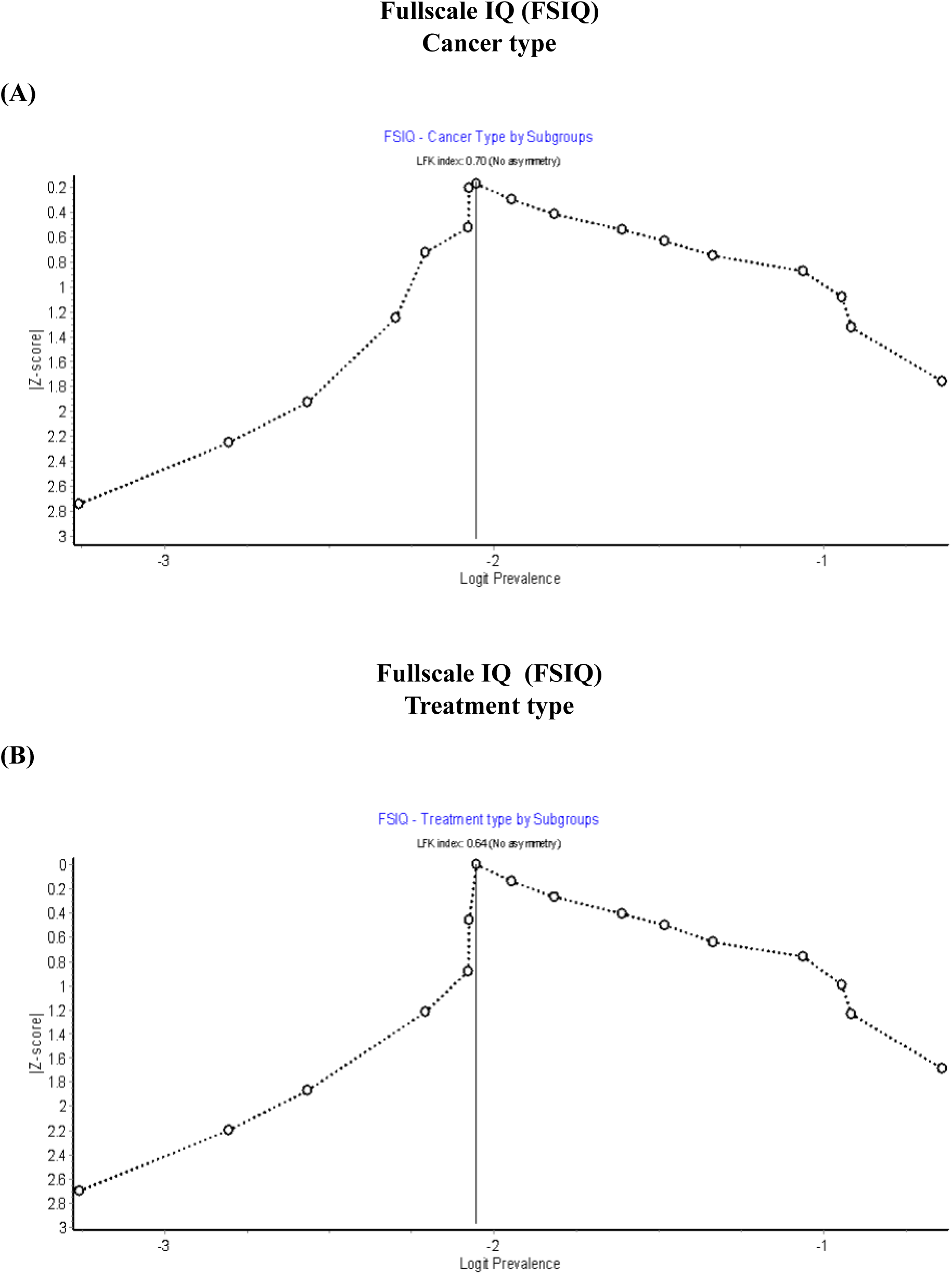
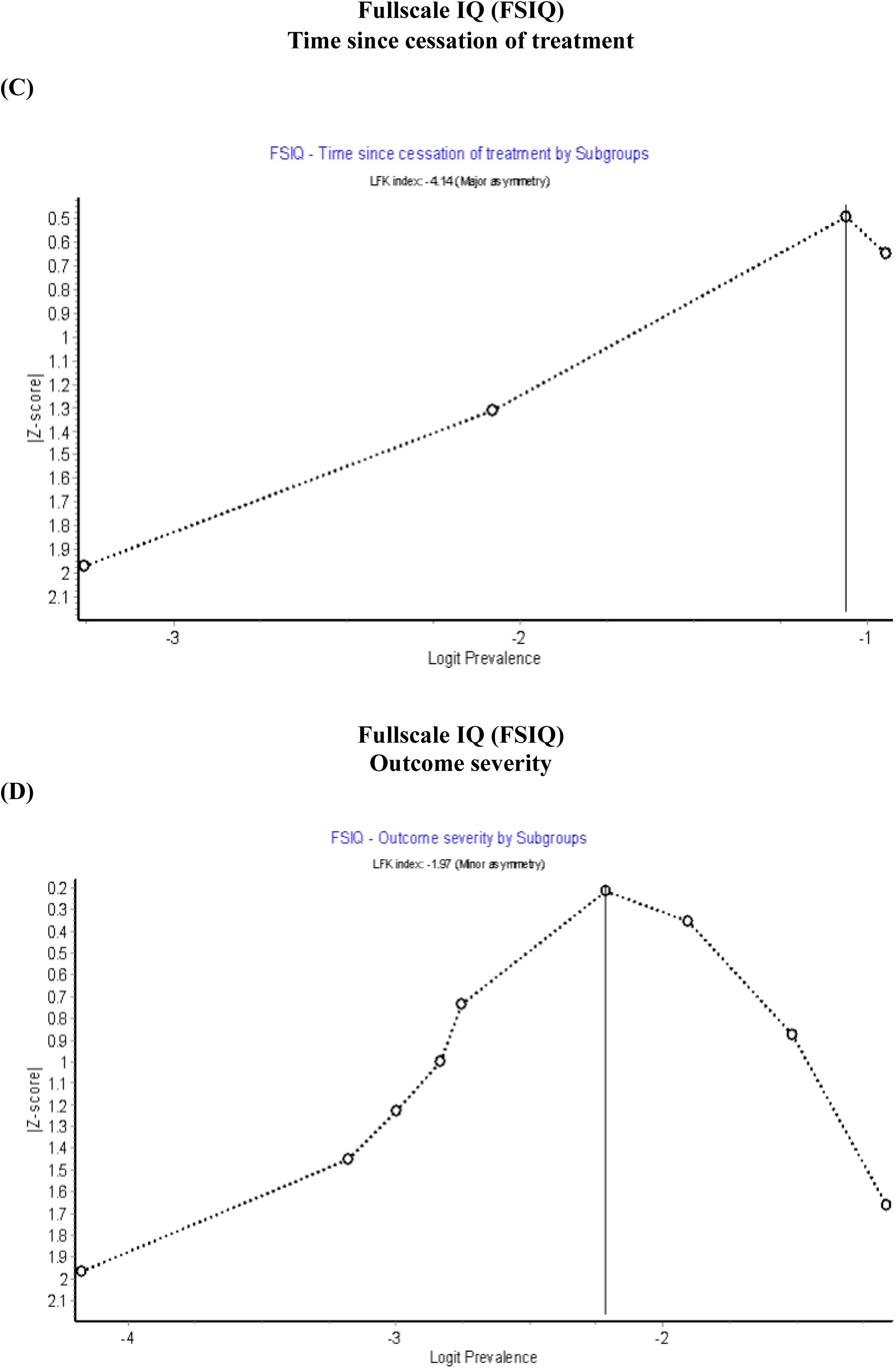
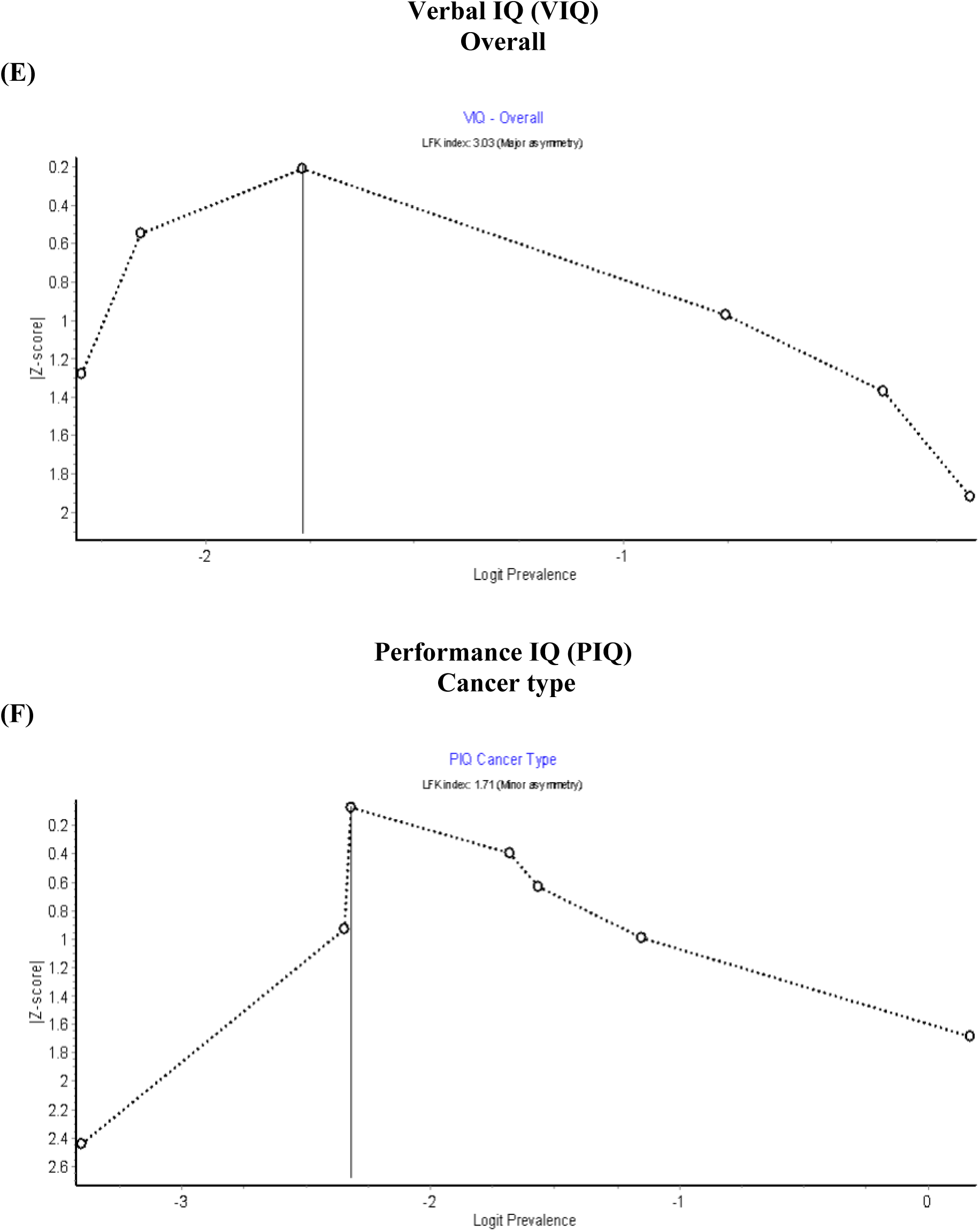
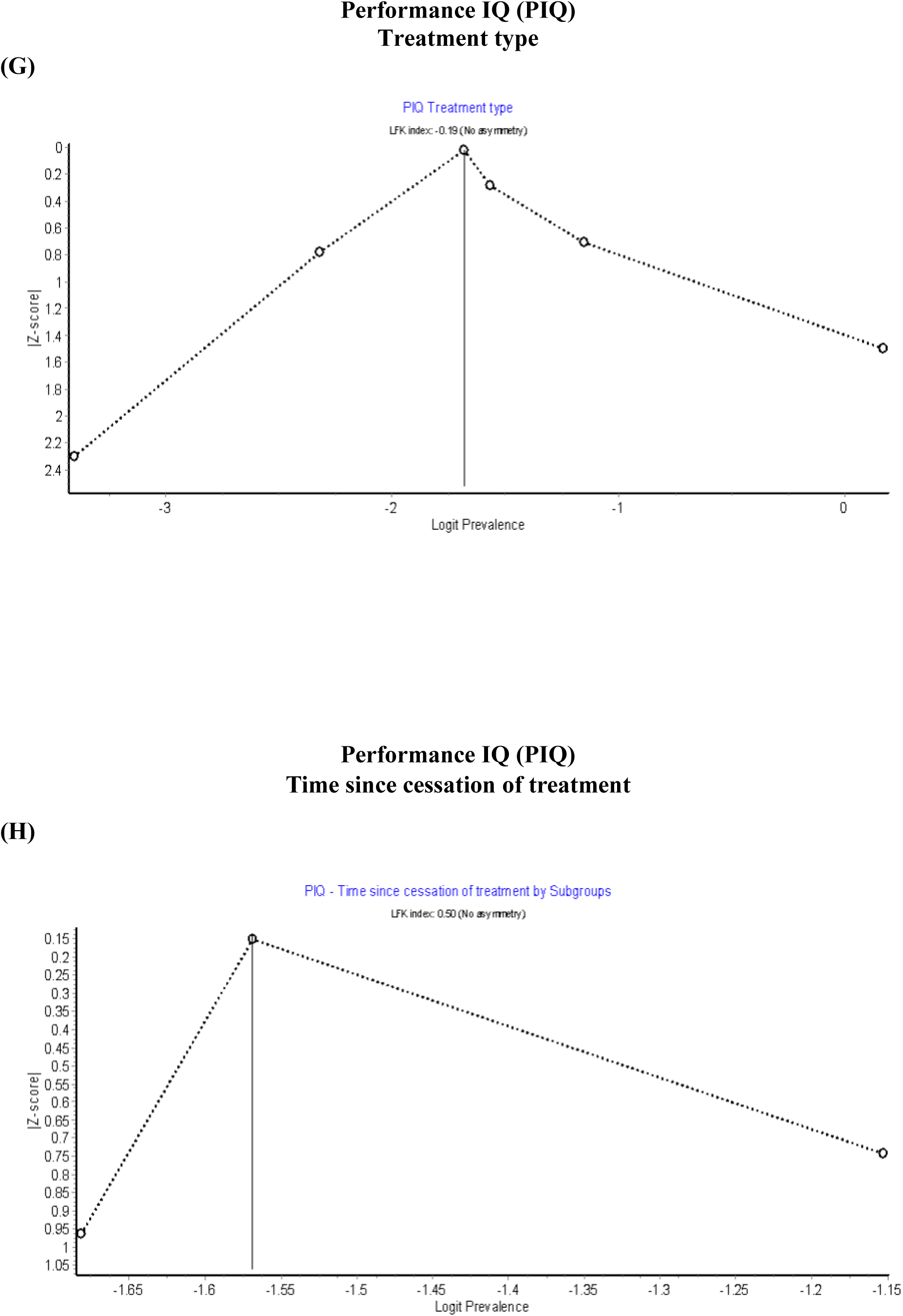
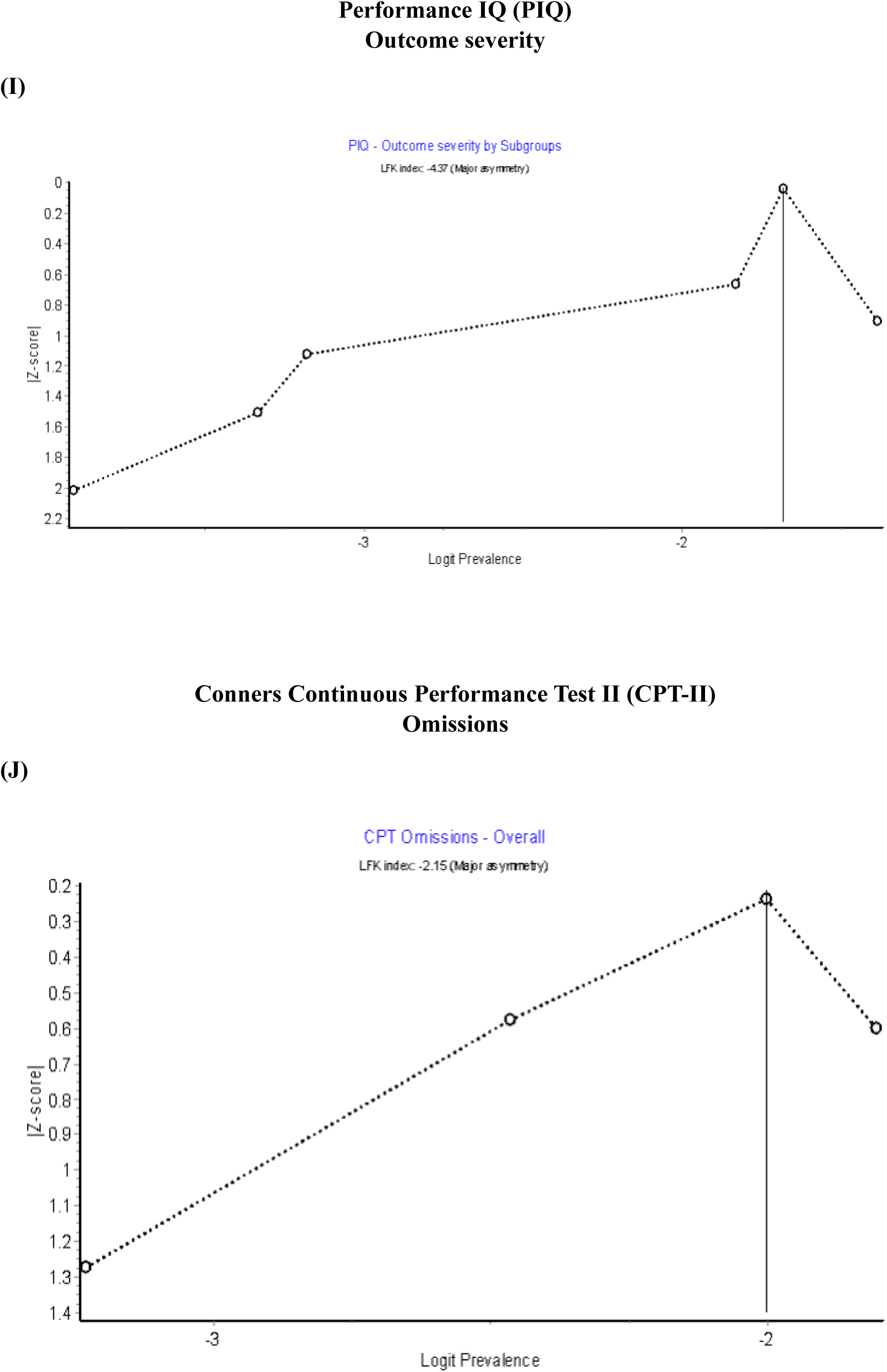
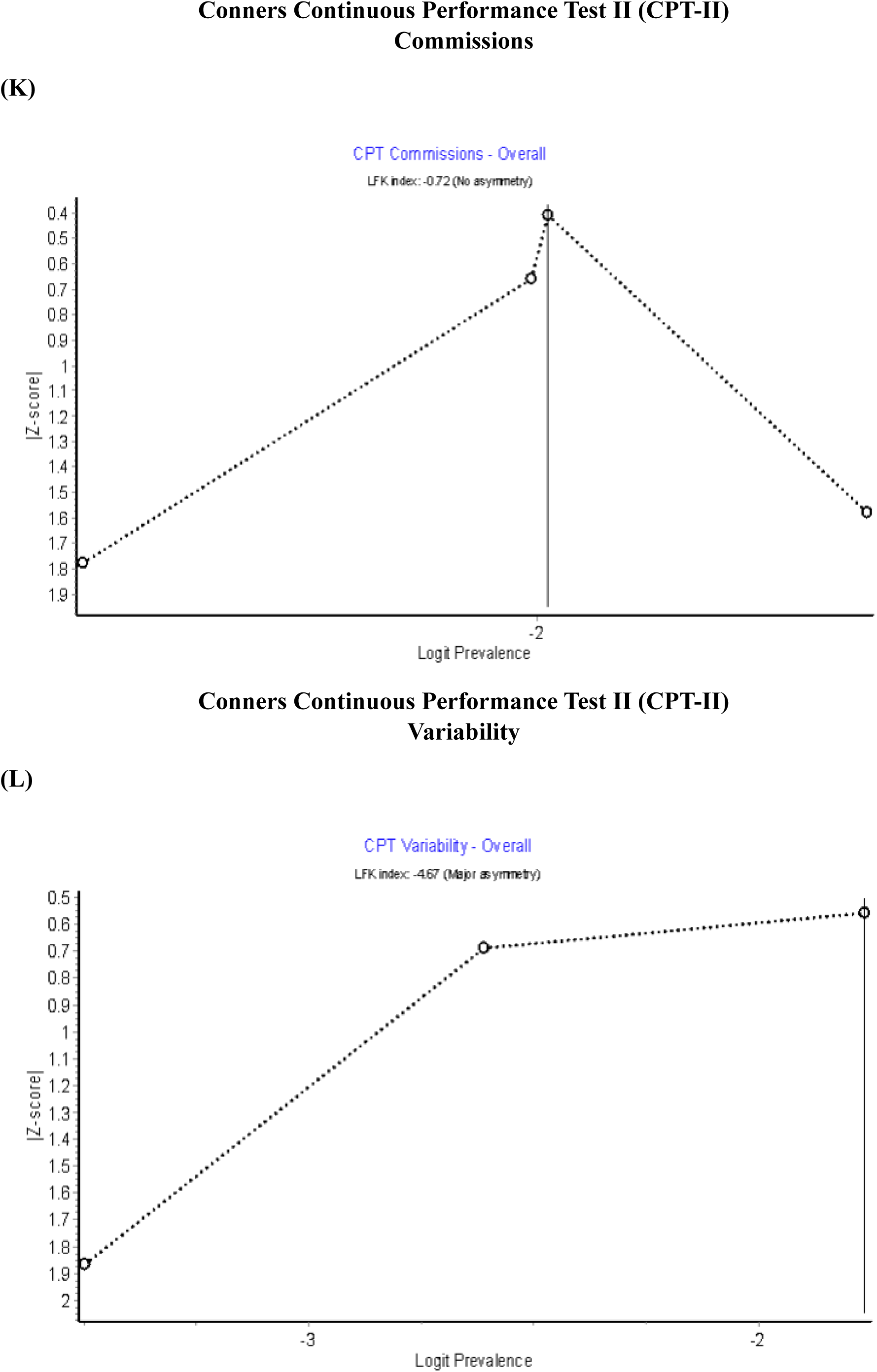

**Figure 1.** Doi plots and LFK index for Fullscale IQ (FSIQ) (A) cancer type, (B) treatment type, (C) time since cessation of treatment, (D) outcome severity, (E) Verbal IQ (VIQ), and for Performance IQ (PIQ) (F) cancer type, (G) treatment type, (H) time since cessation of treatment, (I) outcome severity, and for Conners Continuous Performance Test II (CPT-II) (J) omissions, (K) commissions, and (L) variability.

